# PRISM-AD quantifies clinical and genetic contributions to cognitive risk prediction

**DOI:** 10.64898/2026.11.03.26367206

**Authors:** Yulin Dai, Cassidy S. Wood, Xiaoqian Jiang, Yu-Chun Hsu

## Abstract

**Background:** Older adults with high inherited Alzheimer’s disease (AD) susceptibility can still be cognitively normal (CN). An overall risk estimate alone does not show the cognitive and genetic contributions within a forecast.

**Objective:** To quantify clinical and genetic contributions to cognitive prognosis.

**Methods:** PRISM-AD combined nonlinear clinical functions and a polygenic risk score (PRS) term in 4,435 European-ancestry participants. Participant-level internal validation evaluated first recorded mild cognitive impairment (MCI) after CN in 908 participants and AD dementia after MCI in 714, within three or five years. Separately fitted comparators used identical inputs. Post hoc clinical-course comparisons used regression with robust standard errors and Benjamini–Hochberg (BH) correction.

**Results:** PRISM-AD using current assessments achieved average precision of 0.2996 and 0.4094 for three- and five-year CN-to-MCI, with estimates close to logistic regression. No PRISM-AD PRS increment met BH q < 0.05. Three-year first-MCI forecasts characterized risk contributions in 893 initially CN participants. A retrospective comparison over available follow-up included 95 participants with later AD and 701 with CN at their last diagnostic assessment. Modeled genetic factors were higher in later AD in nine of ten shared cognitive-risk deciles. After adjustment for modeled cognitive risk and fold, the genetic-factor ratio was 1.135 (nominal 95% CI, 1.073 to 1.201, BH q < 0.001).

**Conclusions:** Higher genetic contributions were associated with later AD at comparable modeled cognitive risk. These exploratory profiles require independent validation before use in cognitive follow-up or prevention research.

## Introduction

Cognitively normal (CN) older adults vary in their risk of later impairment, motivating efforts to identify those who may warrant closer cognitive follow-up or evaluation for prevention research. Clinical stage also defines the setting for Alzheimer’s disease (AD) care: lecanemab slowed decline in early symptomatic AD,^1^ while amyloid and tau biomarkers support biological diagnosis and blood-based assays improve pathology detection in symptomatic settings.^2,3^ Within this clinical course, forecasting first recorded mild cognitive impairment (MCI) after CN addresses the emergence of recognized impairment, whereas forecasting AD dementia after MCI addresses progression once impairment is present. The latter course remains uncertain because MCI has heterogeneous causes and can revert to CN.^4,5^

Cognitive assessment provides clinical information for these forecasts. The Alzheimer’s Disease Sequencing Project (ADSP) Phenotype Harmonization Consortium (ADSP-PHC, hereafter PHC) harmonizes memory, executive function, language and visuospatial scores across neuropsychological batteries.^6^ PHC studies have characterized trajectories around incident AD,^7^ and National Alzheimer’s Coordinating Center (NACC) data have supported cognitive-status prediction.^8^ In the Anti-Amyloid Treatment in Asymptomatic Alzheimer’s (A4) and Longitudinal Evaluation of Amyloid Risk and Neurodegeneration (LEARN) studies, cognitive concerns and computerized testing combined with plasma p-tau217 helped predict decline.^9^ Inherited susceptibility provides a different type of information. Genome-wide association studies (GWAS) implicate amyloid and tau pathways, lipid processing and immunity in AD susceptibility.^10^ A polygenic risk score (PRS) summarizes associated variation without measuring pathway activity or pathological burden. In the Alzheimer’s Disease Neuroimaging Initiative (ADNI), a polygenic hazard score improved MCI-to-AD prediction when added to cognition and structural magnetic resonance imaging (MRI).^11^ Cognition and PRS can therefore be examined as distinct inputs to the same prognostic model.

Some older adults retain normal cognition despite a high AD PRS.^12^ Genetic-risk contrasts have also informed resilience research in other conditions, including schizophrenia.^13^ In AD, preserved cognition may reflect lower pathology or better function at a given pathological burden.^14^ Education is used as a proxy for cognitive reserve,^15^ whereas amyloid positron emission tomography (PET) can identify AD-related pathology despite normal cognition. Cognitive level, decline and pathology-adjusted performance capture different aspects of resilience.^16–20^ Comparing inherited susceptibility among people with similar modeled cognitive contributions can reveal genetic differences across subsequent clinical courses that cognitive status alone does not capture.

An overall risk estimate does not, by itself, show how measured cognition and inherited susceptibility contribute to a forecast. Neural additive models represent predictors with learned nonlinear functions that enter the prediction directly,^21^ and explainable AD models have combined PRS with clinical records and post hoc attribution.^22^ Building on this work, PRISM-AD combines separate clinical functions, prespecified clinical interactions and a distinct PRS term. The clinical functions allow nonlinear relationships between measured performance and subsequent impairment, while the PRS term retains inherited susceptibility as a separate contribution.

Summing these contributions on the log-odds scale and converting the total to a probability produces the forecast. Each contribution can be examined directly, with higher values indicating higher predicted risk when the other terms are held constant.

We developed and internally evaluated PRISM-AD in European-ancestry (EA) participants with linked ADSP genotypes and longitudinal cognitive assessments, forecasting first MCI after CN and, secondarily, AD dementia after MCI within three or five years (Fig. 1). Predictions were compared with separately fitted logistic regression, histogram-based gradient boosting (HGB) and TabPFN, a strong tabular prediction benchmark,^23^ using the same clinical and genetic inputs. Matched refits assessed whether adding PRS improved prediction, while the fitted terms described its contribution to each forecast. Post hoc analyses described cognitive and genetic profiles across inherited risk and later clinical courses, focusing on preserved cognition at high PRS and genetic differences at comparable modeled cognitive risk. Follow-up analyses examined sensitivity to unequal diagnostic observation, while separate education and CN-stage amyloid PET analyses provided context on cognitive reserve and AD-related pathology.

**Figure 1.**
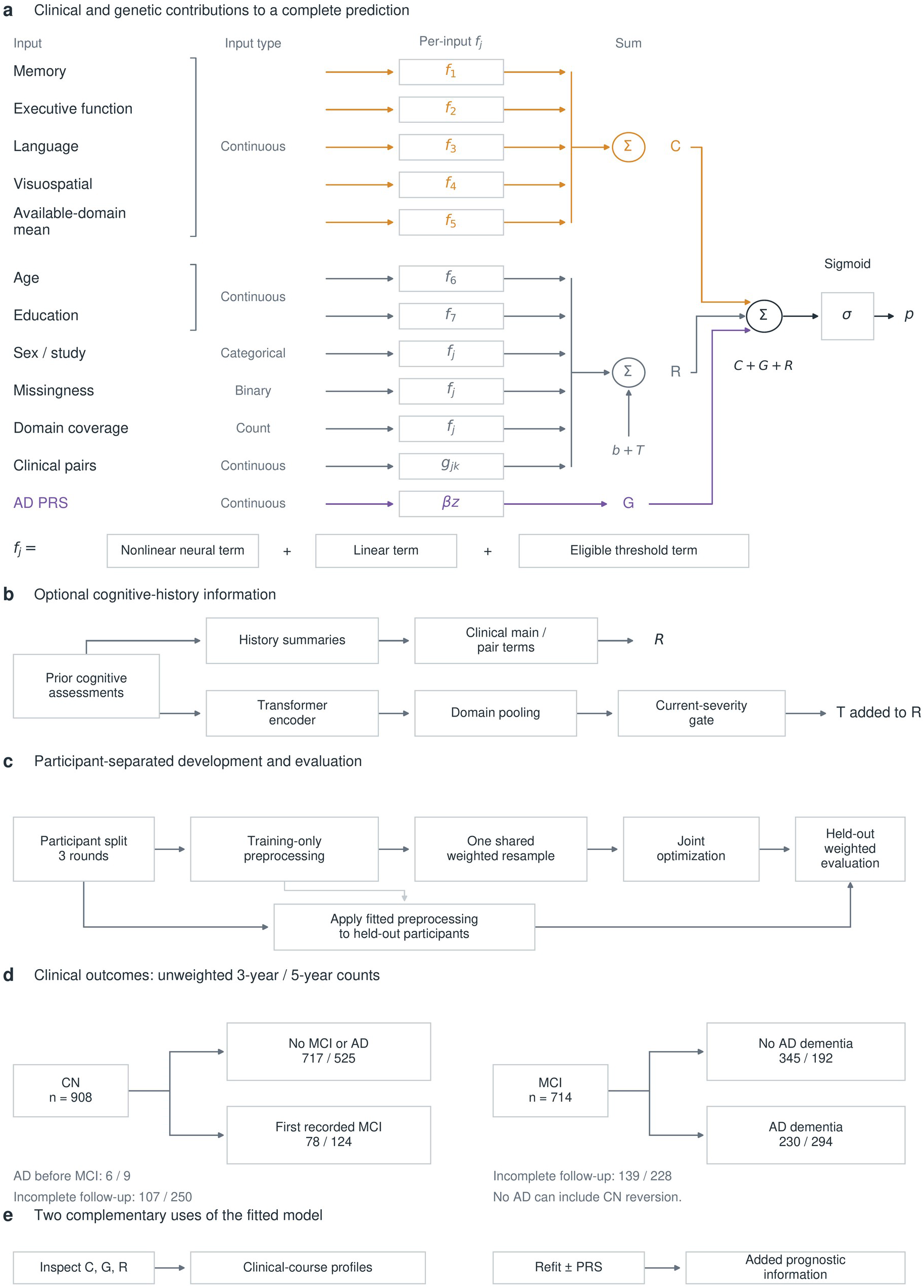
PRISM-AD architecture and stage-specific prognostic evaluation. **a,** Continuous, categorical, binary and count inputs enter separate clinical functions. Learned nonlinear functions, linear terms and eligible threshold terms describe each clinical input. The four cognitive domains and their available-domain mean sum to the cognitive contribution C. The standardized PRS forms a separate linear contribution G with a non-negative coefficient. It does not enter clinical functions or interactions. Remaining clinical functions, prespecified interactions, the intercept b and optional history term T form R. The sigmoid converts C + G + R from log odds to probability p. Higher terms increase predicted risk with other terms fixed. **b,** Cognitive assessments preceding prediction supply summary features and a Transformer-based term T, both included in R. Models using current assessments omit these history pathways. **c,** Participants used for evaluation were excluded from training in each round. Preprocessing was fitted to the training data, and models in the shared-resample comparison used the same sample drawn using censoring weights. Predictions in participants excluded from training were evaluated with censoring weights. Three internal validation rounds use a ten-fold assignment, with exploratory fold 0 excluded. Logistic regression, HGB and TabPFN are independently fitted comparators. **d,** Primary CN-to-first-MCI and secondary MCI-to-AD-dementia forecasts. Paired unweighted counts refer to three and five years among 908 CN and 714 MCI participants at first eligible held-out visits. Target events, observed non-events and unknown outcomes are distinguished. Direct AD before MCI is shown separately. Absence of AD after MCI may include CN reversion. Each participant contributes once per task and may enter both tasks (Table 1, Supplementary Fig. 2). **e,** Fitted contributions describe prediction profiles (Figs. 3–5), whereas matched refits assess added PRS information (Fig. 2c,d). These analyses distinguish the composition of a forecast from the predictive gain attributable to adding PRS. Implementation is given in Methods and Supplementary Methods.

**Table 1.** Participants and recorded CN-to-MCI and MCI-to-AD outcomes at first eligible validation visits.

| **Measure** | **CN to MCI: 3 years** | **CN to MCI: 5 years** | **MCI to AD: 3 years** | **MCI to AD: 5 years** |
|---|---|---|---|---|
| Participants at risk | 908 | 908 | 714 | 714 |
| Known horizon outcomes | 801 | 658 | 575 | 486 |
| Target events | 78 | 124 | 230 | 294 |
| Unknown horizon outcomes | 107 | 250 | 139 | 228 |
| Weighted event fraction | 8.62% | 15.34% | 35.19% | 47.88% |

Outcomes were first recorded MCI after CN and AD dementia after MCI, occurring within three or five years of the first eligible visit. CN-to-MCI was the primary endpoint and MCI-to-AD the secondary endpoint. One visit per participant was selected separately within each task, and the same participant could enter both tasks. Known outcomes include a target event or competing event by the horizon, or observed follow-up through the horizon without either event. Direct recorded CN-to-AD before MCI is a competing event and is not an MCI target. Participants with unknown outcomes remain in the reported denominators but do not contribute to the supervised prediction loss. Weighted event fractions use inverse-probability-of-censoring weights and are not unweighted population incidence estimates.

## Methods

### Data resource and polygenic risk score

We linked processed EA genotype data from ADSP accession NG00067.v17 to longitudinal phenotypes from ADSP-PHC. The ADSP-PHC cognition dataset was obtained from the National Institute on Aging Genetics of Alzheimer’s Disease Data Storage Site (NIAGADS) on 16 December 2024. The accompanying data dictionary and README describe the harmonized domains, visit definitions and diagnostic coding, with further details on the sequencing resource and phenotype harmonization methods reported elsewhere.^6,24^ The four harmonized cognitive domain scores were obtained from version 2 of the ADSP-PHC cognition dataset. These scores measured memory, executive function, language and visuospatial performance and were retained in PHC units.^6^ After genotype quality control (QC), 26,390 samples were retained. We included biallelic single-nucleotide polymorphisms with dbSNP identifiers, retained variants with gnomAD non-Finnish-European alternate-allele frequency (AF_nfe) of at least 0.01 or a missing frequency coded as a period, and required variant and per-chromosome sample call rates of at least 98% and 95%, respectively. Genome-wide KING filtering used a kinship cutoff of 0.0884, followed by selection within four standard deviations of the 1000 Genomes European-ancestry reference centroid on the first two principal components (PCs). PRS values and the genotype sample list matched one to one after quality control. The earlier quality-control framework is described separately.^12^

The APOE-inclusive PRS was calculated with LDpred2-auto^25^ using the GRCh38-harmonized Schwartzentruber et al. summary statistics (GWAS Catalog accession GCST90012877), which combined clinically diagnosed AD with a UK Biobank family-history proxy phenotype.^26^ We restricted the GWAS summary statistics to HapMap3+ variants and matched them to quality-controlled ADSP genotypes. Linkage disequilibrium (LD) was estimated directly from these genotypes within windows defined by genetic positions interpolated from the Eagle GRCh38 map. LDpred2-auto combined the GWAS effect estimates with this LD matrix to estimate variant weights, which were used to calculate each participant’s PRS as a weighted sum of genotypes. Scores were calculated for all post-QC participants, including those with MCI, before applying stage-specific eligibility criteria. PRS values linked to the phenotypes matched those in the full genotyped sample. Genotype quality control and LD estimation were completed before prediction-fold analyses, without reselecting the score using clinical progression outcomes. All prediction models used the same variant weights and raw participant-level PRS values. PRS was standardized using the training participants and eligible visits for each clinical transition. The available score documentation did not establish whether study participants overlapped the discovery GWAS. The effective information of the supplied meta-analysis effects was estimated from their standard errors using the 99.9th percentile of 8 / *S E*^2^ in the retained variant set, giving approximately 90,081. This estimate reflects the information on the supplied effect-size scale and cannot be interpreted as the combined participant count. LD-score regression used the same 618,470 variants represented in the LD-score sums. All downstream models and contribution analyses were fitted using the resulting PRS (Supplementary Methods).

### Eligible participants and clinical outcomes

Eligibility for first-MCI and AD-dementia prognosis was assessed at each CN and MCI visit, respectively, using documented diagnoses rather than PRS. Earlier MCI or AD was defined from diagnoses preceding prediction in the recorded visit order. A later diagnostic reversion did not invalidate an otherwise eligible prediction visit. For retrospective clinical-course comparisons, the last diagnostic assessment was the most recent visit with a documented cognitive diagnosis, which could precede the final study visit. The development analysis included ADNI, NACC, the Religious Orders Study (ROS) and the Rush Memory and Aging Project (MAP). Only visits before age 85 were used, so the five-year prediction window ended before ages recorded as 90 years or older. At least two cognitive domains had to be measured at the prediction visit, and a cognitive diagnosis had to be recorded at a later visit.

Within the linked development population, we identified 1,594 otherwise eligible CN visits and 927 MCI visits excluded solely because they had no subsequent assessment with a cognitive diagnosis at a later recorded age. These exclusions removed all otherwise eligible visits for 179 CN and 248 MCI participants, respectively. Other participants retained earlier eligible visits. Cohort-specific exclusions within this linked population are reported in Supplementary Methods. Cohorts were selected before model fitting according to diagnosis and cognitive-measure availability, independently of predictive performance. In particular, the published harmonization study notes that MCI is generally not diagnosed in the Adult Changes in Thought (ACT) study.^6^

For the CN-to-MCI analysis, participants were initially CN, with no earlier recorded MCI or AD diagnosis. Follow-up began at each eligible CN visit and continued until the first recorded MCI diagnosis, an AD diagnosis without a preceding MCI record, or censoring. MCI was the target event, and AD without an earlier MCI record was a competing event. For MCI-to-AD, every eligible MCI visit before the first AD record could initiate follow-up, including MCI visits after a return to CN. Recorded returns to an earlier diagnostic category were retained. The PHC diagnosis dictionary codes non-AD dementia together with unavailable diagnosis as missing, so these records could not define a separate competing event and did not extend follow-up with known cognitive status. Death dates were not incorporated into the outcome definition.

Follow-up therefore ended according to the cognitive-assessment rules below, without treating death as a separate event. Risk estimates therefore concern recorded diagnoses under these observation rules and do not account separately for competing mortality.

For each prediction horizon *h* (three or five years), a positive outcome indicated that the first recorded target diagnosis occurred within *h* years after the prediction visit. A target event recorded within this window established a positive outcome without requiring further follow-up to *h*. A negative outcome required observed follow-up through *h* without the target event or, in the CN-to-MCI analysis, a recorded AD diagnosis by *h* before any MCI record. The latter was a competing event, not evidence of preserved cognition. If follow-up ended before *h* without a recorded target or competing event, the outcome remained unknown rather than negative. This distinction between target events, competing events and censoring follows fixed-horizon risk-prediction conventions.^27^

If the first target or competing event was undated or had a top-coded age (reported only as a lower bound), follow-up was censored at the last preceding assessment with an exact age and a documented non-target diagnosis. Visits without a documented cognitive diagnosis did not extend follow-up. The three- and five-year estimates refer to recorded clinical diagnoses, rather than biological disease onset or diagnostic lead time. Similar horizons have been used for progression to MCI or dementia among cognitively normal individuals with subjective cognitive concerns.^28^

### Clinical predictors and preceding cognitive assessments

Clinical predictors included age at assessment, sex, years of education, the four cognitive domain scores, their mean across available domains and the number of measured domains. Four indicators identified the contributing study. Models fitted with PRS also included the genetic score, whereas models fitted without PRS used the same clinical measures alone. We compared models using measurements from the prediction visit with models that also incorporated earlier cognitive assessments. Diagnosis, participant identifiers, fold identities, future visits, outcome times and future measurements were excluded from the predictors.

We averaged available domain scores to summarize observed cognitive performance. This descriptive mean was not separately calibrated as a global cognition measure. Missing numerical predictors were imputed with the corresponding training-data medians, with indicators identifying missing measurements. Numerical predictors were scaled by their training-data interquartile ranges, using alternative scales for nearly constant variables and retaining cognitive scores across their observed range.

Earlier assessments contributed to cognitive history only when both the recorded visit order and the assessment age preceded the prediction visit. Rates of cognitive change were estimated from earlier scores, excluding the current assessment. Twenty-one summaries described the number, time span, domain coverage and recency of prior measurements, the average rate of change across domains, and the most recent value, recency, rate of change and observation count for each domain. The four most recent eligible prior visits were ordered chronologically, with four cognitive domains per visit. For each domain measured at a prior visit, the model received the score, standard error, precision, indicators of missing standard error or precision, log-transformed time since that visit and visit number. Unavailable domain measurements were masked. Current and past cognitive values used the same raw PHC units before training-only transformations. HGB and TabPFN models incorporating prior assessments received the same summaries and ordered measurements as PRISM-AD, flattened with the corresponding masks.

### Model development and participant-level validation

The existing ten-fold participant assignment (seed 42) kept each person’s visits together in folds 0–9. Folds 1–9 supplied development data. Folds 1, 2 and 3 each served once for validation while the other eight development folds supplied training. Fold 0 was excluded from this stage-transition analysis but had been used in earlier project development, so it was not an untouched test set. Preprocessing and censoring models used training participants only. This design comprised three rounds of participant-level internal validation within a ten-fold assignment. Evaluation used no independent external cohort. Sample sizes followed from the available eligible linked records, without a prospectively recruited target sample.

Two clinical transitions, two horizons, three validation folds and eight models yielded 96 fits. Models were trained using repeated eligible visits. Primary evaluation used the first eligible visit for each transition in participants excluded from that model’s training set. Analyses using all eligible visits or participant subgroups were secondary. No additional model was trained solely on each participant’s first eligible visit. Settings were fixed for these fits, with no hyperparameter search, validation-based early stopping or checkpoint selection. Predicted probabilities were not recalibrated. Results from these folds were available during earlier analysis development. Specifications were held fixed for the corrected-PRS reconstruction after that exploration. No prospective record established that all architecture, feature, PRS and analysis choices had been finalized before inspection of folds 1–3. The results therefore represent internal development and evaluation.

### Accounting for incomplete follow-up

To account for incomplete follow-up, we estimated the probability of continued observation using reverse Kaplan–Meier curves from all eligible training visits, stratified by study and age bands below 70, 70 to below 80, and 80 to below 85 years. Strata with fewer than 50 visits used their study curve, with a pooled fallback if a study was absent. Both target events and direct-AD competing events contributed as non-censoring observations when fitting the reverse Kaplan–Meier curves. Visits with a target or competing event recorded within the prediction window were weighted by the inverse probability of continued observation just before the event.

For visits observed through the horizon without an event, the probability just before the horizon was used in the same weighting rule. Visits with unknown horizon outcomes contributed to the training censoring model but received zero weight in the classification loss and evaluation. Weights were capped at 10, but no known training or validation weights required clipping. Validity depends on sufficient observation probability and conditional independence of censoring within the specified strata. Age was included in both the predictors and the censoring strata, but performance was evaluated across the eligible age distribution rather than standardized to a fixed age. Observation weights could not be supplied directly to TabPFN. To provide a common training sample, observations with known outcomes were sampled with replacement in proportion to their censoring weights, using a fixed seed. Seven models used this resample, whose size was the smaller of the number of observations with known outcomes and 20,000. All models using the resample received the same observations in the same order with the same labels. Observation weights were not applied again during fitting. A separate HGB model incorporating prior assessments was trained on all observations with known outcomes, with censoring weights applied directly. Validation used all eligible validation observations with their censoring weights and did not involve resampling. Training weights did not equalize each participant’s total contribution across repeated visits.

### Quantifying clinical and genetic contributions to predictions

PRISM-AD used cognitive measurements, clinical context and PRS to estimate the probability p of a specified transition within the prediction horizon. Separate functions for memory, executive function, language, visuospatial performance and their available-domain mean summed to the cognitive-test risk component C. The linear PRS term defined G, whereas the intercept and remaining clinical, interaction and optional history terms formed R (Fig. 1a):

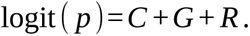

The logistic function converts the summed log odds to the event probability p. C is the contribution assigned to measured cognition on the model’s risk scale, not the cognitive test score itself. With G and R fixed, higher C increases predicted odds and probability, and lower C reduces them. G has the same directional interpretation when C and R are held fixed. These signed components are on the log-odds scale, and their exponentials multiply to give the complete predicted odds. Zero is a model reference, rather than a threshold for normal cognition or absence of inherited susceptibility.

Each clinical measurement or indicator was represented by a small neural network and a linear term, with an additional piecewise-linear function defined by training-data quantiles where eligible. Prespecified clinical interactions captured joint associations among selected measurements, while PRS entered only through its separate coefficient. The coefficient was constrained to be non-negative, although G could be negative because PRS was standardized relative to the training distribution. Architecture dimensions, the complete interaction list and the threshold functions are given in Supplementary Methods and Supplementary Table 1.

To incorporate earlier cognitive assessments, we added their summary measures to the clinical predictors and used one Transformer layer to represent the ordered prior measurements (Fig. 1b). Measurements were pooled within domains to produce a bounded history contribution T, which was zero when no earlier assessment was available. Models using current assessments omitted this pathway. The Supplementary Methods section describes the representation of earlier cognitive assessments, handling of missing domain measurements, domain weights and severity-dependent scaling.

Both PRISM-AD versions were fitted with AdamW for 60 fixed epochs on the shared training resample, jointly optimizing feature functions, interactions, the PRS coefficient and any history parameters. The final epoch was retained without validation-based checkpoint selection. The optimization settings and regularization equations are provided in Supplementary Methods, and training and validation loss trajectories are shown in Supplementary Fig. 1. Recorded contributions summed to each held-out predicted logit. The terms describe how the model forms its prediction and do not uniquely partition biological influences. C contains the specified cognitive main functions, while selected interactions and the optional history pathway carry additional cognitive information within R.

Logistic regression, HGB and TabPFN were fitted independently using matched inputs. TabPFN used the fixed TabPFN-3 classifier in package version 8.2.0. Comparator settings and the inference-only correction for reproducible stochastic preprocessing are documented in Supplementary Methods. That correction changed neither the training observations nor model parameters.

### Added prognostic information from polygenic risk

To assess whether PRS added prognostic information beyond the clinical and cognitive measurements, we compared independently fitted models using these measurements with and without PRS. Logistic regression, HGB, TabPFN and PRISM-AD using current assessments were fitted without PRS for both outcomes, both horizons and validation folds 1–3, giving 48 additional fits. Clinical features, participants, outcome definitions, training-only preprocessing, censoring weights, sampled training rows and their order, model settings and random seeds were held fixed. No genetic principal components or other genotype-derived predictors entered the models fitted without PRS. The PRS column was removed before fitting the tabular comparators. For PRISM-AD, the genetic term was disabled and its parameter frozen before a fresh fit. Models without PRS were trained independently, with the genetic term disabled during fitting rather than removed from an already fitted model. Earlier cognitive assessments were excluded from both input sets.

Before comparing performance, we verified that the models fitted with and without PRS used the same participants in the same order, with matching outcomes, censoring weights and training resamples. We also confirmed that PRISM-AD fitted without PRS had a zero genetic contribution. Performance was evaluated at the first eligible visit of participants excluded from training, pooling the three validation folds. Differences were calculated as performance with PRS minus performance without PRS. Nominal percentile 95% confidence intervals (CIs) used 500 paired participant-bootstrap resamples within fold, conditional on fitted prediction and censoring models. Censoring-weighted average precision (AP) was the primary discrimination measure. Area under the receiver operating characteristic curve (AUROC), the Hájek-normalized Brier score and mean risk were also reported (Supplementary Tables 2 and 3). Both input sets used the same genotyped population without comparison-based tuning. Eight exploratory PRISM-AD PRS-increment tests (two tasks, two horizons, AUROC and AP) used the bootstrap method below and a separate Benjamini–Hochberg (BH) family (Supplementary Table 4).

### Prediction performance and statistical inference

Discrimination was measured by censoring-weighted AUROC and AP. AP summarizes precision across recall levels and differs from trapezoidal precision– recall area, accuracy and precision at a single threshold. We assessed the accuracy of predicted probabilities using the Brier score normalized by the sum of inverse-probability-of-censoring weights (IPCW, Hájek normalization). Mean predicted risk and weighted observed event fractions were compared descriptively (Supplementary Table 3). Their agreement does not establish calibration across the risk range. Performance was reported separately for CN-to-MCI and MCI-to-AD. Additional calibration summaries used fixed-width bins at first eligible visits for the PRS comparison and at all eligible visits for the benchmark models. The weighted summaries describe participants with known outcomes and were not used to recalibrate the models. First-visit calibration bins, support counts and weighted event fractions for PRISM-AD are shown in Supplementary Table 3. Prespecified paired comparisons evaluated PRISM-AD incorporating prior assessments against PRISM-AD using current assessments, HGB and TabPFN incorporating prior assessments, and a second HGB model trained by applying censoring weights directly rather than resampling. Percentile 95% intervals used 500 participant-bootstrap resamples within validation fold. Each sampled participant retained all relevant visits and paired predictions. These intervals are conditional on the fitted prediction and censoring models and do not capture model-refitting, selection or earlier adaptive-development uncertainty. No multiplicity-adjusted discovery claim or non-inferiority margin was specified. We also counted predictions for which three-year risk exceeded five-year risk, leaving their order unchanged after observing results. Reporting of the development-validation design, analysis units, censoring and comparator construction was informed by the TRIPOD+AI guideline for clinical prediction models.^29^

Additional analyses compared PRISM-AD using current assessments with each comparator at the same first eligible visits using 500 within-fold participant-bootstrap resamples (Fig. 2b). Exploratory two-sided P values used 9,999 paired participant-bootstrap resamples within folds (seed 42), centered on the observed AP difference to approximate its null distribution. P was (1 + the count of resampled absolute deviations at least as large as the observed absolute difference)/10,000. Resampling retained all visits, paired predictions and fixed censoring weights. Supplementary Table 5 gives nominal P values and Benjamini–Hochberg q values for 28 first-visit AP contrasts (12 using current assessments and 16 incorporating prior assessments).

**Figure 2.**
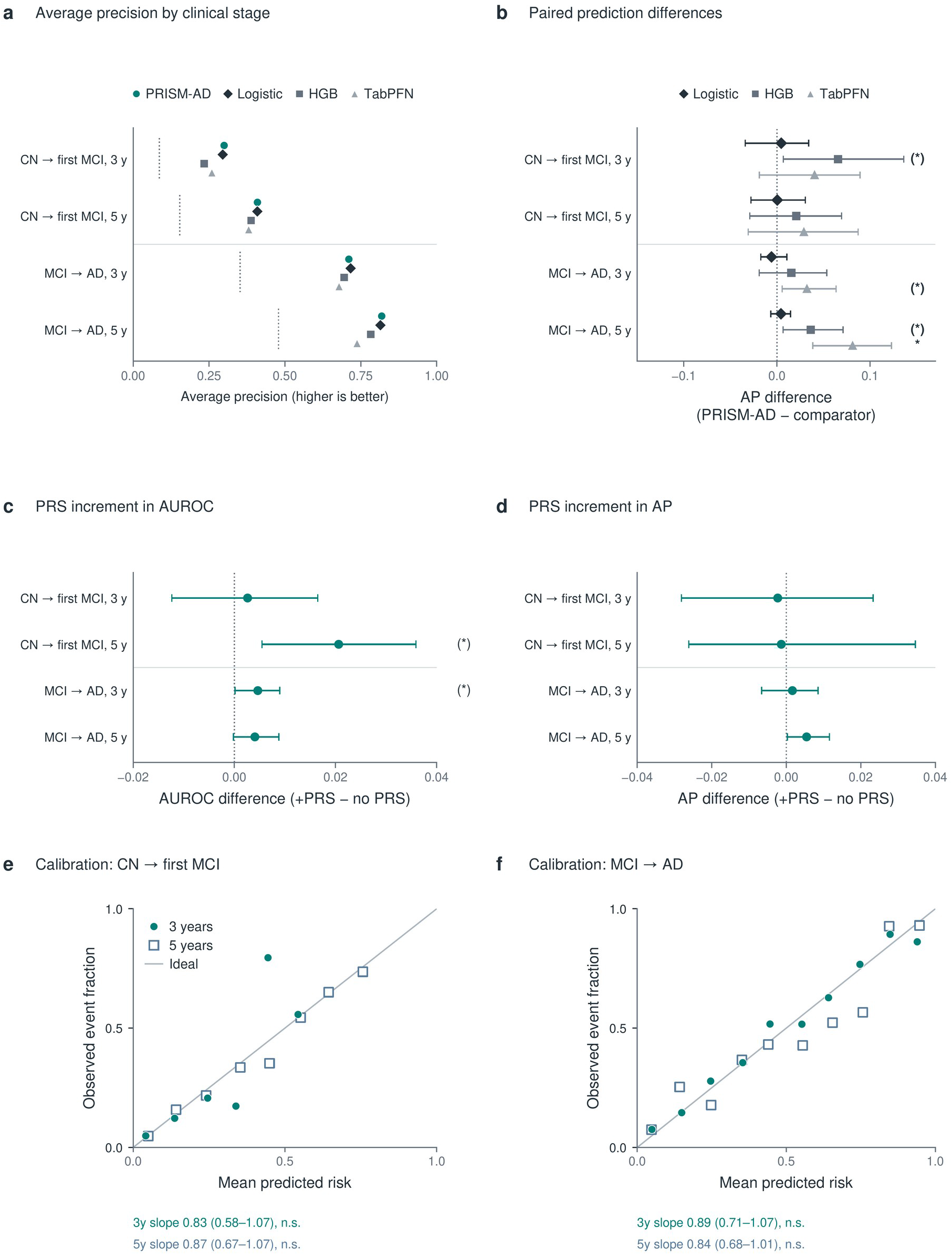
Prediction performance, PRS increments and probability calibration. **a,** Censoring-weighted average precision (AP) at first eligible held-out visits for PRISM-AD, logistic regression, HGB and TabPFN, all using current assessments and PRS. Dotted markers indicate the weighted event fraction. AP depends on event frequency and cannot be compared directly between clinical transitions. **b,** Paired AP differences, PRISM-AD minus each comparator. Lines are nominal 95% percentile intervals from 500 participant-bootstrap resamples within fold. Two-sided tests use 9,999 null-centered paired participant-bootstrap resamples. BH correction covers all 28 first-visit AP comparisons (Supplementary Table 5). **c,d,** PRS-augmented minus clinical-only PRISM-AD AUROC (c) and AP (d), using matched participants, clinical inputs, resamples and settings. Intervals use 500 resamples. BH correction covers eight task-by-horizon-by-metric tests (Supplementary Table 4). Ordinary *, ** and *** denote q < 0.05, 0.01 and 0.001, while (*) denotes nominal P < 0.05 with q *≥* 0.05. All displayed forecasts were evaluated in participants excluded from training. Inference is conditional on fitted models and censoring weights, without refitting or external validation. Positive differences favor PRISM-AD in b and adding PRS in c,d. **e,f,** Descriptive calibration of PRISM-AD using current assessments and PRS for CN-to-first-MCI (e) and MCI-to-AD (f). Filled teal circles denote three years and open blue squares five years. Within each fixed risk bin, mean predicted risk and recorded event fractions were weighted for censoring and pooled across folds using the sum of those weights. The thin gray diagonal marks equality. Known-outcome support was 801 and 658 for CN-to-MCI and 575 and 486 for MCI-to-AD at three and five years. Each point represents a risk bin with at least five known outcomes. Unsupported bins are omitted, and points are not connected or smoothed. Rows below the axes report calibration slopes with nominal normal 95% intervals from IPCW-weighted logistic diagnostics with a free intercept and HC1 participant-score sandwich covariance. Tests assess slope = 1, not a difference between horizons. These four tests share a new ten-test BH family with the six marginal comparisons in Fig. 3e,f. The label n.s. indicates nominal P *≥* 0.05. No bin-wise calibration intervals are shown, and forecasts were not recalibrated. Sparse upper-risk CN bins limit interpretation (Supplementary Table 3).

The 16 comparisons of models incorporating prior assessments at all eligible prediction visits formed a separate secondary family. Tests used saved predictions without refitting the prognostic models. The 500-resample percentile intervals are nominal and were not obtained by inverting these tests. Neither P nor q accounts for earlier adaptive development or establishes non-inferiority. The symbol (*) denotes nominal P < 0.05 with q ≥ 0.05. Ordinary stars denote BH q < 0.05, 0.01 or 0.001.

Post hoc calibration diagnostics estimated a free intercept and slope from individual first-eligible forecasts and known horizon outcomes using IPCW-weighted logistic regression. Intervals used participant-score sandwich covariance with HC1 finite-sample correction, and two-sided tests assessed whether the slope differed from 1. These four tests and the six marginal contribution contrasts below formed one additional ten-test BH family (Figs. 2e,f and 3e,f, Supplementary Tables 3 and 4F). Diagnostic fits were not used to recalibrate forecasts.

**Figure 3.**
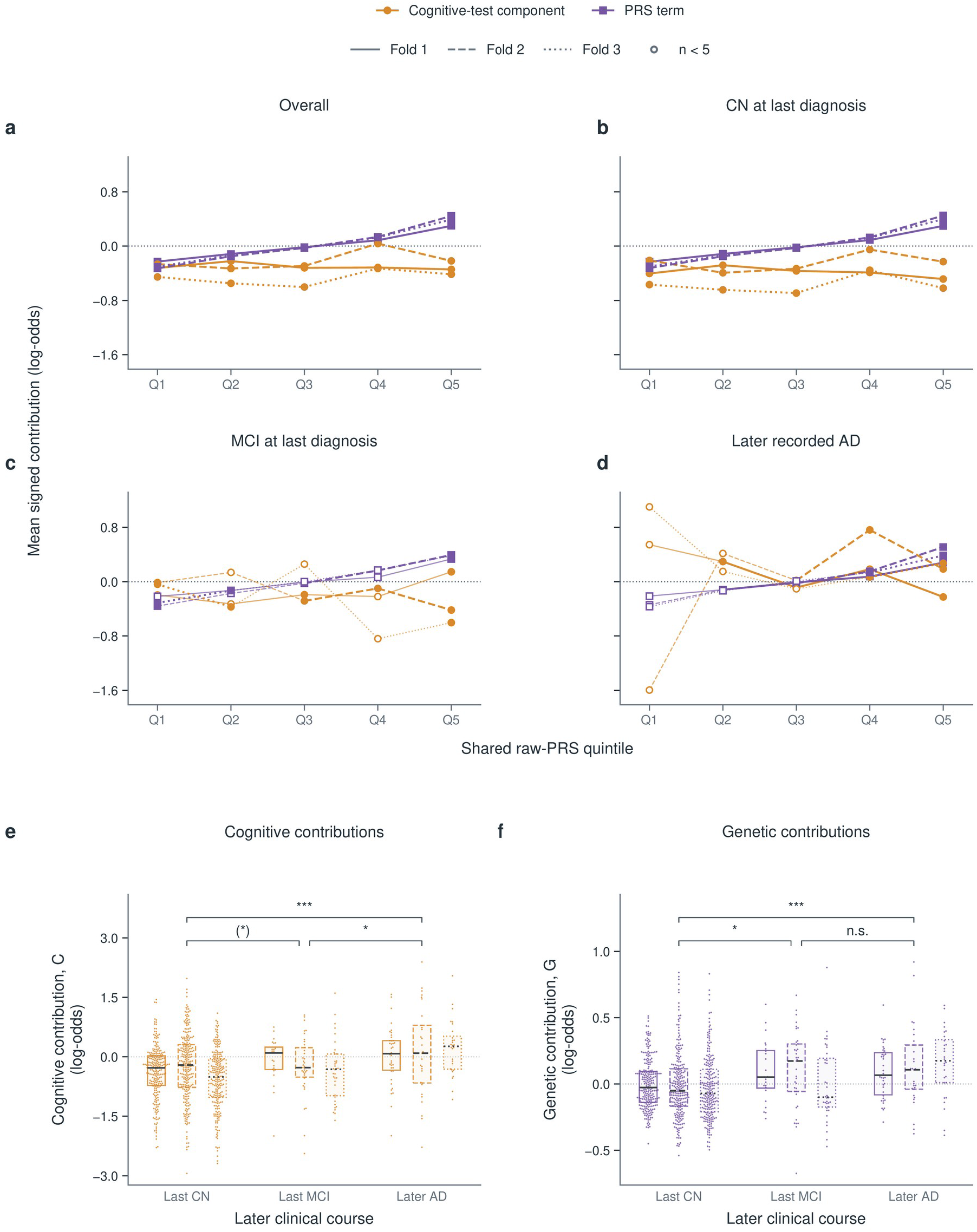
Cognitive and genetic contributions across inherited risk and clinical courses. **a–d,** Overall profiles and profiles of participants with normal cognition at their last diagnostic assessment, MCI at that assessment and later AD. All 893 participants were CN at prediction. Their subsequent clinical course was defined over available follow-up, rather than a fixed three-year AD window. The broader sample has no additional three-year non-AD follow-up requirement. Shared raw-PRS quintiles were defined in the complete sample. Orange lines show C, the sum of five current cognitive main functions, and purple lines show the distinct PRS term G. Line styles identify fitted folds. Higher terms increase predicted odds with the other terms fixed. Zero indicates a zero model contribution, rather than absence of susceptibility. Complete predictions additionally require the remaining terms. Lines connect unadjusted bin means, while open markers and thinner adjacent segments identify cells with fewer than five participants. Increasing G follows the PRS specification. Four participant-level, fold-adjusted HC3 omnibus F tests assessed C differences across PRS quintiles, with BH correction across the four groups (Supplementary Table 4). No omnibus test met BH q < 0.05. Ordinary stars denote BH q thresholds, and (*) denotes nominal P < 0.05 with q above threshold. These tests do not establish equality or biological independence. Figure 5d–g instead groups participants by their modeled cognitive contribution. **e,f,** Individual cognitive (e) and genetic (f) contributions in the same 701 last-CN, 97 last-MCI and 95 later-AD participants. Each dot represents one participant. Horizontal packing separates nearby observations without changing their signed vertical values, and all observations, including the tails, are retained. Boxes span participant interquartile ranges, with internal lines marking medians. These summaries describe participant spread and are not confidence intervals. Solid, dashed and dotted summaries identify folds 1, 2 and 3, arranged from left to right within each clinical course. C and G retain separate vertical-axis scales. Brackets compare clinical-course groups within each component using fold-stratified Wilcoxon tests, not C versus G or differences between folds. A separate ten-test BH family covers these six comparisons and the four calibration slopes in Fig. 2e,f. Ordinary *, ** and *** denote q < 0.05, 0.01 and 0.001. (*) denotes nominal P < 0.05 with q *≥* 0.05, and n.s. denotes nominal P *≥* 0.05. These post hoc marginal comparisons do not adjust for C, age, study or follow-up. They differ from the PRS-quintile tests in a–d and the comparable-C analysis in Fig. 5 (Supplementary Table 4F).

### Contributions across clinical courses

We examined fitted functions from all twelve CN-to-MCI PRISM-AD models across two input versions, two horizons and three validation folds. Main-text function plots report models using current assessments, and Supplementary Table 6 additionally reports models incorporating prior assessments. Functions were evaluated on the original measurement scale over the common training-fold 10th–90th percentile range and centered at shared reference values. High-minus-low contrasts summarized changes in the main functions, and pairwise Spearman correlations assessed their rank agreement across fitted models. These summaries excluded interactions and did not establish causal effects or agreement in magnitude.

To describe contributions in participants assessed while CN, we used their saved three-year first-MCI forecasts at the first eligible validation visit. The broader sample comprised 893 participants grouped retrospectively by clinical course: normal cognition or MCI at the last diagnostic assessment without recorded AD, or later AD. Groups were defined over all available subsequent follow-up, without imposing a common three-year AD outcome window. These later diagnoses were used to interpret forecasts and were excluded from the predictors. Follow-up distributions by clinical course and study are reported in Supplementary Table 7. Quintile boundaries defined across the full sample were used to describe cognitive and genetic contributions across inherited susceptibility. Four fold-adjusted omnibus tests used heteroskedasticity-consistent (HC3) standard errors to assess cognitive differences across quintiles, with BH correction (Fig. 3 and Supplementary Table 4). Swarmplots displayed every individual signed cognitive and genetic contribution, with fold-specific medians and interquartile ranges (Fig. 3e,f). Pairwise clinical-course comparisons used fold-stratified Wilcoxon tests with midranks, tie-corrected variance and inverse-(pair-specific stratum size + 1) weights. These marginal comparisons did not adjust for C, age, study or follow-up.

A targeted analysis of 688 participants compared high-PRS participants with normal cognition at their last diagnostic assessment with other CN participants and those later diagnosed with AD. High-PRS selection used the previously defined interval described below. Both non-AD groups required at least three years of follow-up with known cognitive status, allowing intermediate MCI followed by CN. The AD comparator was not PRS-restricted, so these comparisons did not test protection at matched inherited susceptibility. A sensitivity analysis required normal cognition at every subsequent diagnostic assessment. Contributions and total predicted log odds were compared using ordinary least squares with HC3 robust t-based intervals, adjusting for age, age squared, sex, study, PC1–5 and prediction fold. The adjusted sample included 622 participants with adequate group representation. BH correction covered all 128 planned contribution comparisons, with P = 1 assigned to non-estimable comparisons (Fig. 4a,b and Supplementary Table 8).

**Figure 4.**
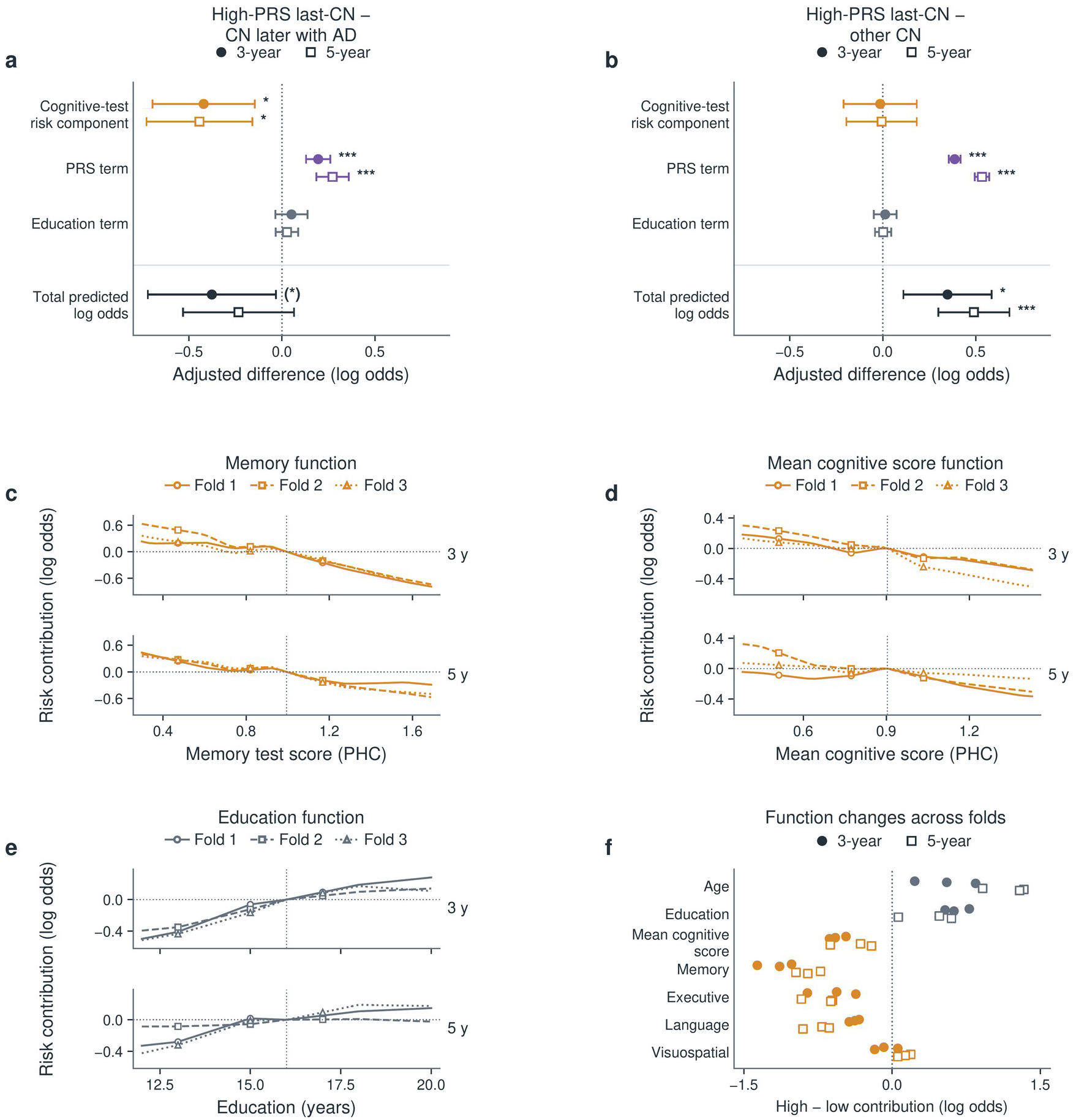
High-PRS clinical-course comparisons and the cognitive functions underlying prognosis. **a,b,** Adjusted contributions from models using current assessments and total predicted log odds in high-PRS CN at the last diagnostic assessment minus later AD (a) or other CN (b). All participants were CN at prediction. The adjusted common-study sample comprised 64 high-PRS last-CN, 63 later-AD and 495 other-CN participants. Negative differences indicate lower values in the high-PRS last-CN group. Both non-AD groups required CN at the last diagnostic assessment and at least three years of follow-up with known cognitive status. Intermediate MCI followed by CN was allowed. The AD comparator was not PRS-restricted, and the genetic contrast partly reflects group definition. C sums four cognitive-domain main functions and their available-domain mean. Cognitive and education terms exclude missingness and interactions. Lower C contributes less predicted first-MCI risk with other terms fixed. Total log odds includes all terms and is not an additional component. Circles and squares denote three and five years, with nominal HC3 robust t-based 95% intervals adjusted for age, age squared, sex, study, PC1–5 and prediction fold. BH correction covers 128 planned contribution contrasts (Supplementary Table 8). Ordinary *, ** and *** denote q < 0.05, 0.01 and 0.001. The symbol (*) denotes P < 0.05 with q ≥ 0.05. Unmarked estimates have P ≥ 0.05. **c–e,** Memory, mean cognition and education main functions from models using current assessments, with three-year plots above five-year plots. Measurements are on their original scales. Higher vertical values represent higher predicted risk with other terms fixed. Functions are centered at shared reference values: memory 0.994, mean cognition 0.904 and education 16 years. Curves span common training-fold 10th–90th percentile ranges, exclude interactions and do not represent individual trajectories. **f,** High-minus-low main-function changes for seven continuous inputs, one point per fitted model and horizon. These descriptive contrasts have no confidence intervals or tests and do not rank causal importance. Training sets overlap across folds.

To examine genetic contributions at comparable modeled cognitive risk, we returned to the broader 893-person sample without the targeted analysis’s additional three-year non-AD follow-up requirement (Fig. 5d–g). Ten shared C deciles were defined once in all 893 participants, rather than separately within clinical-course groups. C is the fitted contribution of the cognitive main functions, distinct from a raw test score or a prediction based on cognition alone. Post hoc comparisons of signed G between later AD and last-CN participants adjusted for C decile, continuous C and fold.

**Figure 5.**
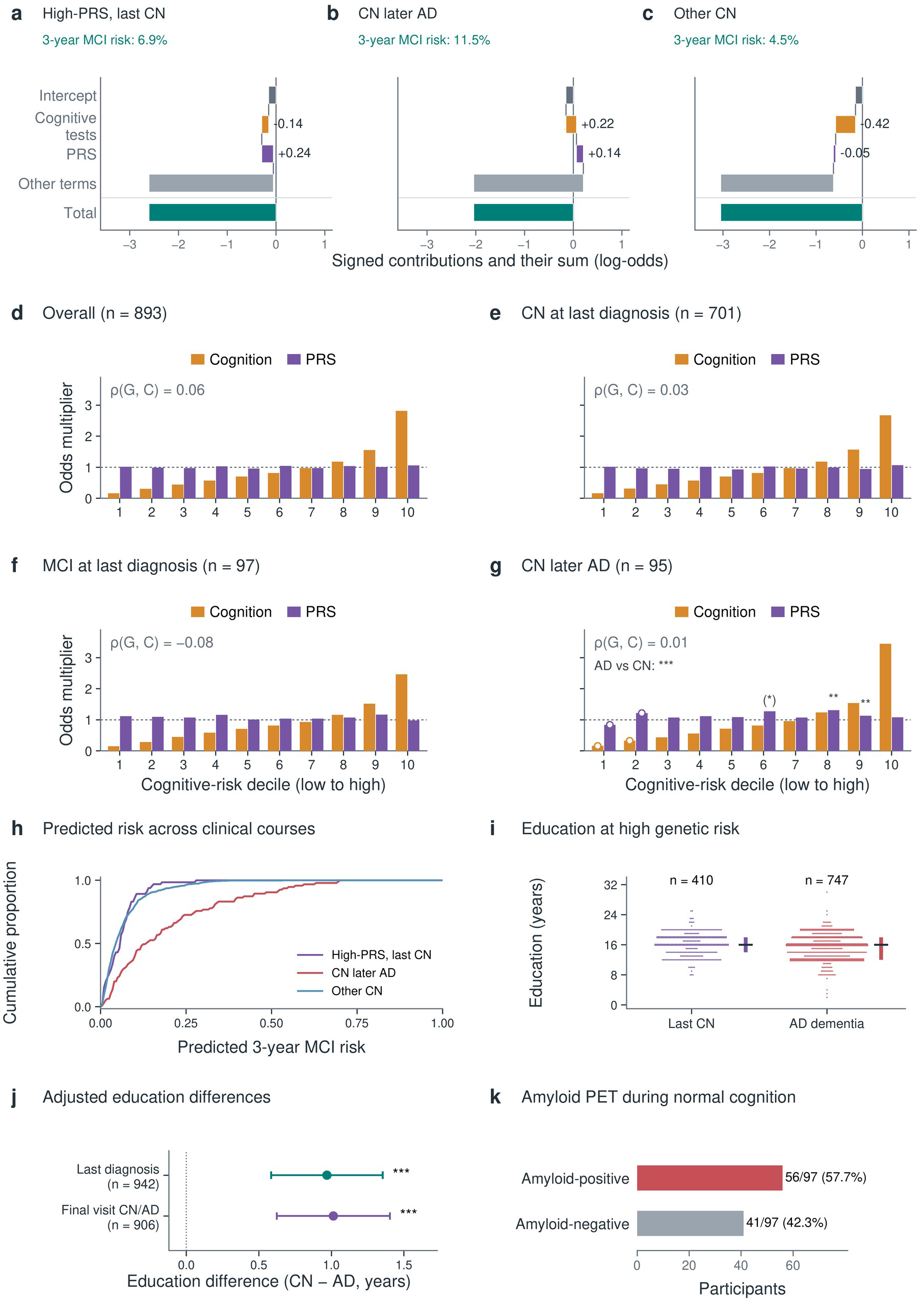
Genetic contributions at comparable modeled cognitive risk during normal cognition. Panels a–g use three-year CN-to-first-MCI forecasts based on current assessments. C sums the five current cognitive main functions, G is the PRS term, and R contains the remaining terms, including the intercept. Higher C or G increases predicted risk with the other terms fixed. **a–c,** Individual fold-1 forecasts for high-PRS last-CN, later AD after CN and other CN. Each was selected nearest the within-group median cognitive, genetic and other contributions after common fold-1 IQR scaling. Predicted probability was not used for selection. Signed bars show the intercept, C, G and Other terms, followed by their total. Applying the sigmoid gives the displayed probability. R is split between the intercept and Other terms, and history is zero. These illustrations show how the terms form complete individual predictions and do not represent group-average effects. **d–g,** Geometric mean cognitive and genetic odds factors in all 893 initially CN participants, last CN without AD (n = 701), last MCI without AD (n = 97) and later AD (n = 95). Ten shared C deciles are defined in all 893 participants. Similar C does not require identical domain scores or equal total predicted risk. No total-risk window or added non-AD follow-up requirement is imposed. Orange bars show exp(*C*), purple bars show exp(*G*), and the dotted reference is 1. These positive factors retain the direction of signed contributions and are not percentages. For an individual, exp(*C*), exp(*G*) and exp(*R*) multiply to give complete predicted odds. Open circles flag cells with fewer than five participants. Bars are unadjusted summaries. Spearman correlations use individual G and C before binning. The rise in C follows the bin definition. In g, stars compare later AD with CN in the same C decile, adjusting for continuous C and fitted fold. They do not compare the two bar colors. The pooled annotation also adjusts for C-decile fixed effects. Differences in G use ordinary least squares, HC3 robust standard errors and two-sided t tests. BH correction includes 12 planned slots: the pooled comparison, ten deciles and expanded covariate sensitivity. Sparse or unsupported tests retain P=1 slots. Ordinary *, ** and *** denote q < 0.05, 0.01 and 0.001, and (*) denotes nominal P < 0.05 with q *≥* 0.05. Genetic factors were higher in later AD in nine of ten bins. The pooled adjusted AD-to-CN factor ratio was 1.135 (nominal 95% CI, 1.073 to 1.201, BH q < 0.001). This post hoc ratio concerns fitted genetic contributions, not clinical AD odds. Fold indicators do not align the genetic coefficient scales, and within-fold and raw-PRS checks are given in Supplementary Table 4D. **h,** Cumulative distributions of predicted three-year first-MCI risk in the targeted sample: 65 high-PRS last-CN, 528 other CN and 95 later-AD participants. Non-AD groups required at least three years of diagnostic follow-up and CN at the last diagnostic assessment, while allowing intermediate MCI. The curves summarize the distribution of model predictions and do not estimate cumulative incidence. **i,j,** Education in a separate high-PRS population aged *≥*75 at last diagnostic assessment. Schooling was observed for 410 of 411 CN and 747 of 751 AD participants (i). The swarm retains every observed value with narrow display-only packing bands. Medians and IQRs use unshifted schooling years. Adjusted CN-minus-AD differences (j) use 293 CN and 649 AD participants in the primary analysis, and 270 CN and 636 AD participants with status confirmed at the final recorded visit. Nominal HC3 robust t-based 95% intervals adjust for age, age squared, sex, study, continuous PRS and PC1–5. Stars use the same q thresholds within four planned education comparisons (Supplementary Table 9). **k,** Amyloid-positive and amyloid-negative participants in the selected older high-PRS last-CN group: 56 and 41 of 97, respectively. PET was obtained during CN, with a cognitive assessment within 90 days and no preceding recorded MCI or AD. Age eligibility refers to the last diagnostic assessment. The median PET-to-last-CN interval was 0.22 years. Education and PET samples have different eligibility rules and may overlap the prediction sample. They provide biological context without establishing independent prognostic validation or sustained resilience.

Within-decile tests adjusted for C and fold, and an expanded sensitivity additionally adjusted for age, age squared, sex, study and PC1–5. HC3 robust t-based inference and BH correction covered twelve contrasts: pooled, ten deciles and expanded sensitivity. Exponentiated differences describe ratios of modeled genetic factors, not AD disease odds. Post hoc sensitivity analyses repeated the AD–CN genetic-factor comparison separately within each fitted fold, retaining the shared C deciles and continuous C adjustment. A pooled comparison instead analyzed raw PRS standardized by the full genotyped population’s standard deviation, adjusting for the same C terms and fold. HC3 t-based inference and a separate four-test BH family were used. These checks did not refit prediction models or alter the original twelve-test family. Full group definitions, contribution summaries, function evaluation and estimability criteria are provided in Supplementary Methods.

To assess sensitivity to unequal diagnostic follow-up, exploratory follow-up analyses included all 908 first-eligible CN prediction participants without selection by later clinical course. We analyzed time to first recorded AD using a Cox working model with fold-stratified baseline hazards, and compared raw PRS by five-year AD status using AD-specific censoring weights. MCI was an intermediate diagnosis and did not terminate observation for AD. Raw PRS was expressed in population SD, with adjustment for fold-specific C terms, age, age squared, sex and study. The Cox model used robust sandwich inference, and the five-year score comparison used HC3 t-based inference, with a separate two-test BH family. Proportional-hazards diagnostics and full outcome/censoring rules are reported in Supplementary Methods and Supplementary Table 4E.

### Education in older high-genetic-risk participants

The exploratory education analysis used the broader post-QC EA genotyped population with linked PHC phenotypes, independently of model-validation eligibility. CN and AD participants were in the same previously defined high-PRS interval (approximately 0.474–1.135) and were at least 75 years old at their last diagnostic assessment. The interval retained the earlier GRPa-PRS outlier-trimming definition, whose upper bound was anchored at the 97.5th percentile of the original CN reference distribution.^12^ This definition selects a bounded subgroup and excludes scores above its upper limit. Scores above the upper bound were outside this selected phenotype but remained eligible for prediction and the unrestricted profiles. The interval was not retuned against the present clinical courses. Its original unrounded bounds were applied inclusively, and training-set PRS standardization did not redefine group membership.

CN participants with any prior recorded AD were excluded. Education in years was taken from the last diagnostic assessment, with negative values treated as missing. Ages reported as 90+ met descriptive eligibility but were not treated as exact ages in adjusted models. A sensitivity analysis required the final recorded visit itself to document the same CN or AD diagnosis.

The CN-minus-AD education difference was estimated by ordinary least squares with HC3 robust t-based intervals, adjusting for age, age squared, sex, study, continuous PRS and PC1–5. Education was not imputed, and current cognition was not included as a covariate. Complete-case analyses required adequate group representation within each retained study. The full selection flow, high-PRS interval definition and criteria for estimating adjusted comparisons are given in Supplementary Methods and Supplementary Table 9.

### Amyloid PET during normal cognition

Amyloid positivity was taken from the PHC-supplied PHC_AMYLOID_STATUS field, with only codes 0 and 1 treated as known. According to its data dictionary, this classification uses thresholds on GAAIN summary cortical standardized uptake value ratio (SUVR) normalized to whole cerebellum. Centiloids are supplied conversions of the same summary measure. No common Centiloid positivity threshold was imposed or estimated in this analysis. Tracer-specific conversions and positivity thresholds are described in the PHC MRI-free PET methods documentation.

Two additional comparisons required CN-stage amyloid PET in both diagnostic groups: one included all amyloid statuses, and the other was restricted to amyloid positivity. Scans required passing image and timing quality control, finite amyloid-burden measurements on the Centiloid scale, normal cognition documented in both PET and paired cognitive records, no preceding MCI or AD, and a scan-to-cognition age difference of at most 90 days. The earliest eligible scan was selected before filtering on positivity. The age 75 threshold applied to the last documented cognitive diagnosis, not to the PET scan. AD cases required a dated first AD diagnosis after joint availability of the CN scan and cognition. PET-subset education came from the paired CN visit, and planned adjustment also included tracer and the interval from that visit to the last documented cognitive diagnosis. Neither PET subset met the same sample-size and study-representation requirements, so no adjusted effects were estimated. Benjamini–Hochberg correction covered all four planned education comparisons, assigning p = 1 to non-estimable tests. The analyses do not estimate an educational intervention effect or preservation at matched pathological burden.

### Genetic susceptibility and amyloid burden during normal cognition

Using the existing one-scan-per-person PET selection, we examined continuous Centiloid burden in participants whose last documented cognitive diagnosis was CN at age 75 years or older. A sensitivity analysis used the broader all-age, last-known-CN sample without redefining the older high-PRS group with normal cognition at the last diagnostic assessment. The high-PRS subgroup retained the previously defined score interval. PRS was scaled by its standard deviation in the genotyped population, so slopes represent the Centiloid difference per one standard-deviation increase in that population. The main ordinary least-squares models adjusted for PET age, sex, PC1–5, study and tracer, with HC3 robust t-based intervals and covariates restricted to information available at or before the scan.

Sensitivity analyses required normal cognition documented at the final recorded visit, restricted the all-age high-PRS group to those with at least three years of follow-up with known cognitive status, or additionally adjusted for the post-PET interval to the last documented cognitive diagnosis. The small follow-up-restricted subgroup omitted tracer to avoid a singleton high-leverage category. Post-PET follow-up duration was not included in the main model because it was determined after imaging. Each participant contributed one CN-stage observation per model, and AD-stage scans were excluded. The small group later diagnosed with AD after a CN-stage scan was summarized descriptively. Prompted by review of earlier exploratory material, these analyses were neither prespecified confirmatory tests nor independent replication, and no multiplicity-adjusted claim was made (Supplementary Table 10).

## Results

### Cognitive stage defined three- and five-year prognostic settings

The linked cohort comprised 4,435 participants from ADNI, NACC, ROS and MAP. We examined first recorded MCI after CN and AD dementia after MCI separately, each within three and five years of an eligible visit (Fig. 1). The CN-to-MCI analysis included 2,759 participants contributing 14,351 eligible visits, and the MCI-to-AD analysis included 2,082 participants contributing 5,692 visits. A total of 406 participants contributed to both analyses at eligible visits. Each participant’s visits were assigned to the same fold. Internal validation evaluated three development folds in turn, with each excluded from that round’s training data. A separate fold was excluded from the present analysis (Methods). Evaluation at the first eligible visit included 908 CN participants and 714 MCI participants (Fig. 1d and Table 1).

Reconstructing diagnostic histories and recorded outcomes for all 20,043 eligible visits identified no entry with an earlier disqualifying diagnosis. Four CN participants later had AD recorded and then returned to CN or MCI at their last diagnostic assessment. They remained eligible for first-MCI prediction and were excluded only from the retrospective clinical-course profiles (Supplementary Methods).

The two transitions differed markedly in frequency. At the first eligible visit, the censoring-weighted fraction with first MCI after CN was 8.62% by three years and 15.34% by five years, whereas the corresponding fractions with AD after MCI were 35.19% and 47.88% (Table 1). First MCI was recorded for 78 of 801 CN participants with a determinable three-year outcome and 124 of 658 with a determinable five-year outcome. Participant characteristics and predictor missingness at the first eligible visit are reported for the development population and evaluated subset in Supplementary Table 11. AP depends on event frequency, so its values cannot be compared directly between the CN and MCI starting groups (Fig. 2a). Age distributions and follow-up availability are shown in Supplementary Fig. 2 and Supplementary Table 12.

### PRISM-AD predicted three- and five-year clinical transitions

The primary first-MCI comparison evaluated participants who were CN at prediction, using measurements from that visit and the same clinical and genetic inputs for PRISM-AD and the separately fitted comparators. Models incorporating prior assessments were evaluated separately. Current-assessment PRISM-AD achieved censoring-weighted AP of 0.2996 and 0.4094 for first MCI within three and five years, respectively (Fig. 2a and Table 2). Logistic regression achieved 0.2951 and 0.4091. Corresponding PRISM-AD AUROCs were 0.7332 and 0.7708.

**Table 2.** Censoring-weighted average precision at first eligible visits.

| **Model** | **CN to MCI: 3 years** | **CN to MCI: 5 years** | **MCI to AD: 3 years** | **MCI to AD: 5 years** |
|---|---|---|---|---|
| Logistic, current | 0.2951 | 0.4091 | 0.7164 | 0.8146 |
| HGB, current | 0.2340 | 0.3887 | 0.6951 | 0.7827 |
| HGB, history (resampled) | 0.2446 | 0.4249 | 0.7021 | 0.7785 |
| HGB, history (native weights) | 0.2615 | 0.4123 | 0.7238 | 0.7688 |
| TabPFN, current | 0.2593 | 0.3805 | 0.6784 | 0.7377 |
| TabPFN, history | 0.2480 | 0.4226 | 0.7130 | 0.7788 |
| PRISM-AD, current | 0.2996 | 0.4094 | 0.7105 | 0.8189 |
| PRISM-AD, history | 0.2916 | 0.3989 | 0.7036 | 0.8082 |

All models in this table included PRS. Values are AP calculated from held-out predictions, not classification accuracy. Current denotes models using measurements from the prediction visit. History denotes separately fitted models that also used summaries of strictly earlier assessments and the last four available prior cognitive visits. Seven models shared the same censoring-weighted training resample. HGB with native weights instead used all training observations with known outcomes and applied their censoring weights directly. Algorithms and horizons were fitted independently. The ordering of point estimates does not establish statistical superiority.

For first-MCI prediction, PRISM-AD minus logistic regression AP differences were 0.0046 (95% CI, −0.0341 to 0.0340) at three years and 0.0003 (−0.0279 to 0.0304) at five years. Differences against TabPFN using current assessments were 0.0403 (−0.0190 to 0.0892) and 0.0289 (−0.0309 to 0.0872). These paired intervals quantify uncertainty in the first-MCI AP comparisons with logistic regression and TabPFN (Fig. 2b and Supplementary Table 5).

For AD dementia after MCI, PRISM-AD achieved AP of 0.7105 within three years and 0.8189 within five years. At five years, logistic regression reached 0.8146. PRISM-AD exceeded TabPFN using current assessments by 0.0812 AP (nominal 95% CI, 0.0385 to 0.1229, BH q = 0.0252). This was the only current-assessment AP comparison to meet BH q < 0.05 within the family of 28 first-visit comparisons, which also included models incorporating prior assessments. The comparison is specific to these clinical transitions, forecast horizons and fitted comparators.

Calibration-bin summaries showed discrepancies between predicted and observed risks in some ranges, with limited support in several bins (Fig. 2e,f and Supplementary Table 3). First-MCI calibration slopes were 0.826 (nominal 95% CI, 0.582–1.069) at three years and 0.868 (0.667–1.070) at five years. Corresponding MCI-to-AD slopes were 0.894 (0.714–1.073) and 0.844 (0.680–1.009). All intervals included 1, and no slope departure met the BH threshold. These global diagnostics do not establish calibration throughout the risk range.

Repeated training under the same computational conditions reproduced parameters and predictions exactly. First-visit three-year CN AP ranged from 0.2160 to 0.3975 across validation folds, illustrating variation between evaluated participant groups. Optimization trajectories are shown in Supplementary Fig. 1.

### PRS increments varied by horizon and metric for first-MCI risk ranking

Matched refits tested whether PRS added predictive information beyond the same clinical and cognitive measurements. Five-year first-MCI AUROC changed from 0.7502 without PRS to 0.7708, a paired difference of 0.0206 (0.0055 to 0.0359) (nominal 95% CI). The three-year difference was 0.0026 (−0.0124 to 0.0165). For MCI-to-AD, AUROC differences were 0.0046 (0.0001 to 0.0090) at three years and 0.0041 (−0.0002 to 0.0088) at five years (Fig. 2c, Supplementary Fig. 3 and Supplementary Table 2).

Of the eight PRISM-AD tests across tasks, horizons and metrics, two reached nominal P < 0.05 and none met BH q < 0.05. The five-year CN-to-MCI AUROC increment had nominal P = 0.0152 and BH q = 0.1216, and the three-year MCI-to-AD AUROC increment had nominal P = 0.0362 and BH q = 0.1448. These exploratory tests concern added PRS information, rather than whether fitted genetic terms differ between clinical-course groups (Supplementary Table 4).

First-MCI AP differences after adding PRS were −0.0023 (−0.0281 to 0.0233) at three years and −0.0013 (−0.0262 to 0.0346) at five years (Fig. 2d). All four model families and both forecast windows are shown in Supplementary Fig. 3. Improvements in ranking do not by themselves establish better probability calibration or clinical benefit.

### Current assessments supported prediction with sparse cognitive histories

Earlier measurements were available before the first eligible visit for only 64 of 2,759 CN participants (2.3%), compared with 446 of 2,082 MCI participants (21.4%). For CN-to-MCI, models incorporating earlier assessments achieved AP of 0.2916 and 0.3989, compared with 0.2996 and 0.4094 using current assessments. At three years, the history-minus-current AP difference was −0.0080 (nominal P = 0.5968), and at five years it was −0.0105 (nominal P = 0.4562). Table 2 and Supplementary Table 5 report all paired comparisons of models incorporating prior assessments, including the HGB sensitivity using observation weights directly.

The two horizons were fitted independently. Their predictions therefore do not define a joint cumulative-risk trajectory. For PRISM-AD using current assessments, three-year risk exceeded five-year risk at 53 eligible CN validation visits and 29 MCI visits. Visit spacing also limits the timing resolution of recorded impairment.

### Contributions described distinct later clinical courses

Across all 24 PRISM-AD fits with PRS, the recorded terms summed to the held-out predicted log odds. With the remaining terms fixed, a higher cognitive or genetic contribution increases the predicted odds and probability of the specified clinical transition. The broader profile analysis included 893 adults assessed while CN: 701 with normal cognition at their last diagnostic assessment, 97 with MCI at that assessment and 95 later diagnosed with AD. The cognitive term describes the contribution of test performance to predicted first-MCI risk, and subsequent diagnoses define the retrospective clinical-course groups.

Figure 3 compares these cognitive and genetic contributions across shared raw-PRS quintiles within each fitted fold. The PRS term increases with PRS by specification. The cognitive contribution need not decrease as the genetic contribution increases, because total risk and the other clinical terms vary across participants. Fold-adjusted omnibus tests gave BH q values from 0.4539 to 0.6816 across the four displayed groups (Supplementary Table 4). The marginal distributions overlapped across clinical courses, but participants later diagnosed with AD tended to have higher cognitive and genetic contributions than those with normal cognition at their last diagnostic assessment (rank-biserial r = 0.359 and 0.317, respectively, both BH q < 0.001, Fig. 3e,f and Supplementary Table 4F). The rank tests compare contributions between clinical courses, while the omnibus tests compare PRS quintiles within each course. The marginal comparisons do not hold C constant or adjust for age, study or follow-up, so they address a different question from the analysis at comparable C below.

### Preserved cognition at high PRS illustrated contrasting risk contributions

The targeted comparison included 65 high-PRS last-CN participants, 528 other CN participants without recorded AD and 95 participants later diagnosed with AD (Figs. 4a,b and 5h). Both non-AD groups required at least three years of diagnostic follow-up and normal cognition at their last diagnostic assessment. Intermediate MCI followed by CN was allowed. The AD comparator was not PRS-restricted. Groups were defined by PRS and clinical course, rather than predicted risk.

The adjusted analysis included 64 high-PRS last-CN participants, 495 other CN participants and 63 participants later diagnosed with AD in studies with adequate group representation. The cognitive-test contribution in high-PRS last-CN participants was lower than in later AD, with adjusted high-minus-AD differences of −0.421 (95% CI, −0.696 to −0.146) at three years and −0.444 (95% CI, −0.728 to −0.159) at five years (BH q = 0.0133 and 0.0122, respectively, Fig. 4a). The model therefore assigned a lower first-MCI risk contribution to their cognitive measurements than to those of the later-AD group. The PRS term was higher by 0.195 and 0.272 log-odds units. This genetic contrast partly reflects high-PRS selection and the model’s non-negative PRS coefficient.

Against other CN participants, cognitive-contribution differences were −0.014 (95% CI, −0.210 to 0.183) at three years and −0.006 (95% CI, −0.196 to 0.183) at five years. The high-PRS last-CN group nevertheless had higher total predicted log odds: 0.348 (95% CI, 0.111 to 0.585) and 0.490 (95% CI, 0.299 to 0.681), respectively (BH q = 0.0171 and < 0.001, Fig. 4b and Supplementary Table 8). Thus, a lower cognitive contribution than in later AD coexisted with higher total predicted risk than in other CN participants. This pattern describes conditional forecasts and does not establish that cognition biologically offsets genetic susceptibility.

### Cognitive functions related test performance to predicted risk

Across the fitted models, higher memory scores and higher available-domain mean scores contributed less predicted first-MCI risk over the common training-supported range. For memory, high-minus-low contrasts ranged from −1.364 to −1.017 log-odds units at three years and from −0.973 to −0.724 at five years. Corresponding contrasts for mean cognition ranged from −0.633 to −0.467 and from −0.626 to −0.207, respectively. These main functions exclude interactions and describe fitted associations rather than within-person trajectories (Fig. 4c,d,f and Supplementary Table 6).

Age, education and the individual cognitive domains contributed distinct fitted functions (Fig. 4c–f). Education terms describe prognosis conditional on the measured cognitive state and the other inputs. They address a different question from the separate comparison of educational attainment between older CN and AD participants. Neither the fitted function nor the between-group education association identifies a causal schooling effect.

### Genetic contributions differed at comparable modeled cognitive risk

The individual examples show how C, G and the remaining clinical terms combine to determine the predicted first-MCI probability (Fig. 5a–c). We then grouped all 893 initially CN participants by shared deciles of C, complementing the PRS-based ordering in Fig. 3. Cognitive factors increase across these bins by construction. Genetic odds factors, exp(*G*), were higher in later AD than last-CN participants in nine of ten deciles (Fig. 5d–g). The comparison uses subsequent diagnoses over all available follow-up, rather than AD diagnosed within a fixed three-year window.

Among 95 later-AD and 701 last-CN participants, the C- and fold-adjusted AD-to-CN genetic-factor ratio was 1.135 (nominal 95% CI, 1.073 to 1.201, BH q < 0.001). Expanded demographic, study and genetic-ancestry adjustment gave 1.158 (nominal 95% CI, 1.095 to 1.224, BH q < 0.001). Deciles 8 and 9 met BH q < 0.01. Decile 6 reached nominal P < 0.05 but not BH q < 0.05. Median diagnostic follow-up was 7.69 years (IQR, 5.50–11.50) in later AD and 6.38 years (4.00–10.20) in last CN. Median time to first AD was 6.68 years (4.07–9.18, Supplementary Table 7). Because follow-up was not matched in this clinical-course comparison, we examined observation time in the additional analyses below. The ratios quantify fitted genetic factors and cannot be interpreted as clinical AD odds. Individual G–C correlations address how the two contributions vary together, rather than the difference between later clinical courses at comparable C (Supplementary Methods).

Within-fold genetic-factor ratios were 1.120 (nominal 95% CI, 1.045–1.199), 1.119 (0.985–1.271) and 1.186 (1.069–1.315) in folds 1, 2 and 3, respectively. All three ratios exceeded 1, but the interval for fold 2 included 1. In the pooled raw-PRS comparison, the later-AD group had values 0.514 population SD higher than the last-CN group after adjustment (nominal 95% CI, 0.295–0.734, Supplementary Table 4D). These analyses conditioned on the same fitted C values and reused the original evaluation participants. The prediction models also had overlapping training samples, so the results do not provide independent replication.

Separate follow-up analyses included all 908 eligible CN prediction participants and identified 99 dated first AD diagnoses. At five years, 31 had recorded AD, 598 were observed without AD through that horizon, and AD status remained unknown for 279. The no-AD group included participants with MCI and those diagnosed with AD after five years. After cognitive and clinical adjustment and AD-specific censoring weighting, raw PRS was 0.652 population SD higher among participants with AD by five years than among those observed without AD through five years (nominal 95% CI, 0.297–1.006, BH q < 0.001). The planned time-to-AD working model also yielded a positive association, but global diagnostics indicated nonproportional hazards (P < 0.001), limiting interpretation of a single hazard ratio across the full follow-up period (Supplementary Table 4E). These checks address observation time but remain exploratory associations rather than independently validated AD forecasts.

### Greater education accompanied preserved cognition at high genetic risk

In a separate genotyped population at high PRS and age 75 or older, 411 participants had normal cognition at their last diagnostic assessment and 751 had AD. Education was available for 410 and 747, with means of 16.31 and 15.37 years, respectively (Fig. 5i). Group membership did not depend on model-validation eligibility or predicted risk.

After adjustment for age, age squared, sex, study, continuous PRS and PC1–5, the CN group had 0.97 (95% CI, 0.58 to 1.35) more years of education, based on 293 CN and 649 AD participants. Requiring an explicit CN or AD diagnosis at the final recorded visit gave 1.01 (95% CI, 0.62 to 1.40) years among 270 CN and 636 AD participants. Both associations met BH q < 0.001 within four planned comparisons (Fig. 5j and Supplementary Table 9). The similar estimates indicate that the education difference persisted when clinical status was documented at the final visit.

### Amyloid pathology coexisted with normal cognition at high PRS

CN-stage amyloid PET was available for 97 older high-PRS participants with normal cognition at their last diagnostic assessment and 6 later diagnosed with AD. Of these, 56 and 5, respectively, were amyloid-positive (Fig. 5k). The median PET-to-last-CN interval was 0.22 years. These scans document amyloid pathology during normal cognition, but the short observation interval limits inference about sustained cognitive maintenance.

In the broader older CN population, the adjusted association with amyloid burden was 14.1 (10.4 to 17.8, n = 802) Centiloids per PRS standard deviation. Within the high-PRS interval, it was 21.6 (−15.3 to 58.5, n = 97). Requiring an explicit CN diagnosis at the final recorded visit gave 5.6 (2.4 to 8.8, n = 577) across all PRS values and −1.2 (−34.1 to 31.7, n = 54) within the high-PRS interval. The all-age high-PRS sensitivity gave 20.0 (6.9 to 33.1, n = 462), whereas the subgroup with at least three years of diagnostic follow-up gave 23.6 (−13.1 to 60.2, n = 48). These exploratory estimates support a PRS–amyloid relationship in the broader CN population, with less precision in restricted subgroups (Supplementary Table 10).

## Discussion

PRISM-AD made cognitive and genetic contributions explicit within first-MCI forecasts and used the same fitted terms to examine subsequent clinical course in adults assessed while CN. At comparable modeled cognitive risk, participants later diagnosed with AD had higher genetic contributions than those with normal cognition at their last diagnostic assessment. Within each fold, G is the training-standardized PRS multiplied by a non-negative fitted coefficient. Its contrast therefore expresses the original score in units specific to that fitted model and provides no independent genetic signal. Fold indicators account for mean shifts but do not align the fitted genetic coefficient scales. The within-fold and raw-PRS checks examined this scale dependence while retaining the exploratory interpretation.

The genetic difference remained positive after demographic, study and ancestry adjustment, but this association addresses a different question from the predictive value of adding PRS. Models fitted with and without PRS assessed changes in prediction performance, whereas the C-stratified comparison examined fitted genetic contributions across later clinical courses. None of the matched PRISM-AD PRS increments met the BH threshold, so the contribution contrasts do not establish improved prediction from adding PRS. Comparable C does not require identical domain scores or equal overall predicted risk. Likewise, the near-zero correlation between G and C describes how the terms vary together across participants and can coexist with genetic differences between clinical courses at comparable C. The separate five-year AD-status analysis also supported a genetic difference over a common outcome window. Its exploratory comparison used participants with and without recorded AD through five years, so its reference group differed from the original last-CN group.

Building on established additive modeling and explainable AD prediction,^21,22^ PRISM-AD uses the fitted clinical and genetic terms both to estimate risk and to describe individual forecasts. Its primary first-MCI AP estimates were close to logistic regression, while five-year MCI-to-AD AP exceeded TabPFN using current assessments after BH correction. Comparisons with independently fitted models using matched inputs provided the predictive context for the contribution analyses. First MCI after CN and AD dementia after MCI address different points in the clinical course, complementing work on cognitive-status categories, diagnosis, trajectories and rapid decline. Biomarker-based diagnosis and prognosis could provide additional information on the recorded clinical transitions examined here.^8,30,31,3,9,32^

Participants with high PRS and normal cognition at their last diagnostic assessment had lower cognitive contributions than those later diagnosed with AD, yet higher overall predicted risk than other CN participants. The targeted CN subgroup was selected for high PRS, whereas the AD comparator was unrestricted, so the genetic contrast partly reflects group selection. A lower cognitive term alongside a higher genetic term describes a prediction profile without establishing biological compensation. Higher memory scores were generally assigned lower first-MCI risk contributions, which may reflect preserved function, premorbid ability or less impairment before clinical diagnosis. The present analyses cannot distinguish these explanations, and correlations among cognitive domains prevent linking individual terms to isolated neural deficits. Allowing intermediate MCI followed by CN also makes the interpretation of preserved cognition dependent on follow-up and outcome definition.^19,20^

In the separate older high-PRS population, CN participants had approximately one more year of education than participants with AD after covariate adjustment, including when diagnosis was confirmed at the final visit. This association is consistent with the use of education as a proxy for cognitive reserve.^15^ Pathology-adjusted studies have associated education more consistently with cognitive level than slower decline, depending on tau burden and outcome definition.^16,33^ In the prognosis model, the positive education term estimated risk conditional on measured cognition and the other inputs, addressing a different question from the comparison of educational attainment. Conditioning on cognition can alter this association,^34^ and neither analysis identifies a causal schooling effect.

Amyloid PET extended our earlier GRPa-PRS work^12^ by showing that 56 of 97 selected high-PRS CN participants were amyloid-positive. The education association and PET findings provide complementary evidence on reserve-related characteristics and measurable AD-related pathology in participants with preserved cognition. The median PET-to-last-CN interval of 0.22 years, small clinical-course groups and unequal study representation limit inference about sustained cognition despite pathology. Longitudinal cognition, amyloid, tau and neurodegeneration measurements could distinguish lower pathology from better function at a given burden.^17,35^

The clinical value of these profiles depends on whether they improve decisions about cognitive assessment and follow-up. Prospective evaluation could compare selection using cognitive and genetic contributions with selection using the complete three- and five-year first-MCI estimate or cognition alone, including participant stratification for prevention research. With assessment capacity held fixed, relevant outcomes include incident-MCI yield, time to recognition, assessment burden and false-positive referrals. These horizons specify forecast windows and have not been shown to provide diagnostic lead time. Predicted first-MCI risk alone does not establish AD pathology or treatment eligibility.

### Limitations

PRISM-AD was evaluated by participant-level internal validation across four development cohorts. Performance in an independent external cohort or a new care setting remains untested. Prognostic analyses were restricted to EA participants with linked genotypes and follow-up with known cognitive status, limiting applicability to other ancestry groups and routine clinical care. Prior model exploration may make performance estimates optimistic, and overlapping training samples limit their independence. Continued decreases in training loss alongside modest late-epoch increases in validation loss for models incorporating prior assessments indicate possible overfitting (Supplementary Fig. 1).

Visuospatial scores were unavailable for 71.8% of the evaluated CN sample and 52.8% of the evaluated MCI sample, limiting interpretation of domain-specific contributions. Sparse prior cognitive assessments and visit spacing limited assessment of change and impairment timing.^36^ Separately fitted three- and five-year estimates can reverse risk ordering and do not define a cumulative-risk curve.

Recorded clinical transitions differ from disease time addressed by biomarker clocks^37^ and disease-progression models.^38^ Joint modeling of transitions and calibration at each horizon for competing outcomes^27^ were outside scope. Outcome-dependent attendance,^39^ participation and survival may affect selection despite censoring adjustment. Death and non-AD dementia were not modeled separately.

Possible overlap with the discovery GWAS could not be excluded. The discovery meta-analysis does not provide variant-level effective sample sizes. The SE-derived effective-information estimate remains an approximation for combined clinical and proxy effects, despite close agreement across the evaluated estimation checks.

Bootstrap intervals omit refitting and adaptive-development uncertainty. The C-stratified comparison was post hoc, and multiplicity correction does not cover earlier project exploration. Conditioning on C and unequal follow-up may affect the original clinical-course contrasts and preclude causal interpretation. The five-year and time-to-event checks do not remove informative ascertainment or uncertainty in censoring weights and fitted C. Global nonproportional hazards also limit the Cox working-model summary. Including cognitive domain scores alongside their mean makes their separate contributions depend on how the model allocates information among correlated inputs. The education and PET analyses provide biological context rather than prognostic validation. Contributions describe conditional predictions, not mechanisms that preserve cognition. Nonsignificant slope tests do not establish adequate calibration, and the marginal contribution contrasts do not remove age, study or follow-up differences. Clinical use requires independent external validation, calibration and prospective evaluation of decision benefit.

## Conclusions

PRISM-AD quantifies cognitive and genetic contributions within the same three- and five-year first-MCI forecasts. In exploratory analyses of adults assessed while CN, those later diagnosed with AD had a higher modeled genetic factor than those with normal cognition at their last diagnostic assessment after accounting for modeled cognitive risk (adjusted ratio, 1.135). These internally derived profiles relate the cognitive and genetic information within a forecast to later clinical course. The findings support prospective evaluation of the profiles for cognitive follow-up and participant stratification in prevention research. Independent external validation is needed before clinical use.

## Acknowledgments

The authors thank the participants, families, investigators and research staff who contributed to ADSP, ADSP-PHC and the source studies. Data for this study were prepared, archived, and distributed by the National Institute on Aging Alzheimer’s Disease Data Storage Site (NIAGADS) at the University of Pennsylvania (U24-AG041689), funded by the National Institute on Aging. ADSP sequencing data generation and harmonization were supported by the Genome Center for Alzheimer’s Disease (U54AG052427). ADSP-PHC was supported by the National Institute on Aging (U24 AG074855, U01 AG068057 and R01 AG059716).

The analyses drew on data from the Anti-Amyloid Treatment in Asymptomatic Alzheimer’s Disease study (A4), Adult Changes in Thought (ACT), Alzheimer’s Disease Neuroimaging Initiative (ADNI), Knight Alzheimer’s Disease Research Center, National Alzheimer’s Coordinating Center (NACC), National Institute on Aging Alzheimer’s Disease Family Based Study (NIA-AD FBS), Religious Orders Study (ROS), Rush Memory and Aging Project (MAP), Washington Heights/Inwood Columbia Aging Project (WHICAP) and Wisconsin Registry for Alzheimer’s Prevention (WRAP). ADNI data collection and sharing were supported by the National Institutes of Health (U01 AG024904) and Department of Defense (W81XWH-12-2-0012). Full data-resource and study-specific funding acknowledgments, including ADNI sponsors and contributing centers, are provided in the Supplementary Acknowledgments. These acknowledgments describe support for the source data resources.

The authors acknowledge the High-Performance Computing for Research facility at the Center for Secure Artificial Intelligence for Healthcare, McWilliams School of Biomedical Informatics, The University of Texas Health Science Center at Houston, for providing computational resources.

## Author contributions

**Yulin Dai:** Conceptualization, investigation, visualization, writing – original draft, writing – review and editing, and funding acquisition. **Cassidy S. Wood:** Investigation, visualization, writing – original draft, and writing – review and editing. **Xiaoqian Jiang:** Supervision, resources, and writing – review and editing. **Yu-Chun Hsu:** Conceptualization, investigation, visualization, writing – original draft, writing – review and editing, supervision, and funding acquisition.

## Statements and declarations

## Ethical considerations

The Committee for the Protection of Human Subjects (CPHS) at The University of Texas Health Science Center at Houston determined that this secondary analysis did not meet the regulatory definition of human subjects research (HSC-SBMI-24-0898, September 2, 2024). No further CPHS review was required.

## Patient and public involvement

Patients and members of the public were not directly involved in the design, conduct or reporting of this secondary analysis.

## Consent to participate

Not applicable.

## Consent for publication

The applicable consent requirements for publication of the deidentified individual forecasts in Figure 5a-c were satisfied. No identifiable participant-level information is reported.

## Data availability

Processed ADSP genotype data were obtained from the NIAGADS ADSP Umbrella dataset, accession NG00067.v17 (https://dss.niagads.org/datasets/ng00067-v17/). Corresponding harmonized phenotypes were provided by ADSP-PHC through NIAGADS. The ADSP-PHC cognition dataset (version 2) was obtained from NIAGADS on 16 December 2024. Access to participant-level data requires an approved NIAGADS Data Access Request and compliance with the applicable data-use certification, consent restrictions and source-cohort conditions, including ADNI access requirements where applicable. Controlled participant-level datasets, including individual predictions and contribution ledgers, are not redistributed. Figure 5a–c shows three selected individual forecasts without participant identifiers, rather than a downloadable participant-level dataset. Aggregate tables supporting the analyses accompany the supplementary material.

## Code availability

Analysis code, synthetic tests, environment specifications, input schemas and figure-generation scripts are publicly available at https://github.com/davidroad/prismAD. The repository also includes a data-free model schematic. The public implementation is release 1.5.0. Public checks use synthetic data. Reconstructing study results requires authorized private inputs, and fitted weights are not redistributed. Controlled ADSP/PHC data, participant-level derivatives, empirical figure source data, cohort summary tables, fitted model weights and access credentials are excluded.

## Study registration and protocol availability

This study was not registered, and no study protocol is publicly available. The post hoc analysis plans described in Supplementary Methods were specified before their corresponding estimates were calculated.

