## Supplementary material for "PRISM-AD quantifies clinical and genetic contributions to cognitive risk prediction"

##### Contents

Supplementary Methods describe the analysis populations, implementation and exploratory inference. Supplementary Tables 1–12 provide component definitions, performance comparisons, cohort accounting and supporting analyses. Supplementary Figures 1–3 show PRISM-AD training and validation loss trajectories, age distributions, and PRS increments across all four model families. Source-cohort acknowledgments follow the scientific supplement.

#### Supplementary Methods

##### Analysis populations

Prediction was evaluated at the first eligible visit in 908 CN and 714 MCI participants (Fig. 2). Contribution profiles included 893 participants assessed while CN and linked to subsequent clinical-course groups (Figs. 3 and 5d–g). The targeted high-PRS illustration used 688 participants, of whom 622 met the adjusted-comparison criteria (Fig. 4a,b). Education and amyloid PET provided supporting context in separate older high-PRS populations (Fig. 5i–k). The samples overlap and address different questions, so they do not provide independent replication. An audit against full diagnostic histories reconstructed all 20,043 eligible prediction visits without a prior disqualifying diagnosis or a saved-outcome mismatch. The 908 CN and 714 MCI first-visit evaluation records matched the saved validation data at both horizons. The retrospective profile exclusions therefore did not require changing the prediction population or refitting models.

##### Data sources and genotyped participants

The phenotype-linked data contained the same 26,390 sample identifiers as the post-KING European-ancestry genotype sample, with a one-to-one mapping between genotype and phenotype identifiers. Their PRS values agreed exactly with the scores calculated in the full genotyped sample. The QC log records a KING kinship cutoff of 0.0884, confirming close-relative filtering without excluding all distant relatedness or duplicates recorded under different identifiers. LD uses the post-QC target genotypes, before stage-specific phenotype eligibility, without supervised progression labels. Whether participants overlapped the discovery GWAS could not be established.

The cognition data dictionary and README describe the dataset date, harmonization method and diagnostic coding. The PET dictionary defines positivity using thresholds on GAAIN summary cortical SUVR normalized to whole cerebellum and provides Centiloid conversions. We used the supplied amyloid-status categories directly, with tracer-specific thresholds and conversions specified in the referenced MRI-free PET methods document. The available A4 image-collection descriptions did not establish which scans came from screening participants and which came from trial enrollees.

The ADSP-PHC cognition dataset (version 2) was obtained from NIAGADS on 16 December 2024 and interpreted using its accompanying data dictionary. The GWAS summary statistics were obtained from the GWAS Catalog under accession GCST90012877. PRS values were calculated for the genotyped study population using the scoring procedure described in the main Methods.

##### Eligibility before assigning horizon outcomes

Prediction-visit selection was assessed in the linked development roster in folds 1–9, rather than in all genotyped participants. Eligible visits required at least two measured cognitive domains. Visits required follow-up with a documented cognitive diagnosis at a later exact age to establish an observation interval. Visits failing this requirement included some with undated first events. An excluded visit could belong to a participant with retained earlier visits. Entirely excluded participants had no otherwise eligible retained visit for that task. CN and MCI participant counts may overlap. Selection before the linked roster was assembled was not reconstructed. IPCW within the retained analysis populations does not address initial attendance selection.

| Task | Study | Before follow-up | Excluded visits | Entirely excluded persons | Retained visits |
|---|---|---|---|---|---|
| CN_MCI | ADNI | 2294 | 405 | 52 | 1889 |
| CN_MCI | MAP - Rush | 438 | 31 | 2 | 407 |
| CN_MCI | NACC | 12235 | 1125 | 120 | 11110 |
| CN_MCI | ROS | 978 | 33 | 5 | 945 |
| MCI_AD | ADNI | 3216 | 448 | 79 | 2768 |
| MCI_AD | MAP - Rush | 176 | 16 | 9 | 160 |
| MCI_AD | NACC | 2862 | 440 | 153 | 2422 |
| MCI_AD | ROS | 365 | 23 | 7 | 342 |

Death dates were not used to assign outcomes, and PHC codes unavailable diagnosis and non-AD dementia together as missing. Such visits did not extend follow-up with known cognitive status. The resulting estimates therefore do not account separately for competing mortality.

**Education selection and high-PRS definition**

The previously defined GRPa-PRS interval was not recalculated as a top quintile in the present sample. Its upper bound was the 97.5th percentile of CN scores in the reference population. The lower bound was obtained by moving 2,392 ranks down the pooled sorted score array from the observation nearest that upper bound, using 11,962 CN and AD participants with finite scores. The interval is approximately 0.474–1.135 on the unstandardized scale. The displayed values are rounded, and eligibility used the original unrounded cutoffs inclusively, independently of training-set PRS standardization. The upper bound follows the earlier outlier-trimmed control reference and deliberately excludes scores above it from this operational subgroup. Those scores were not removed from prediction or from the unrestricted contribution profiles. We retained these bounds without optimizing them against the present clinical courses.

Eligibility required age at the last documented cognitive diagnosis of at least 75 years. CN participants with prior AD were excluded, and MCI was not treated as CN. Education came from the last diagnostic assessment, with negative values treated as missing and no imputation. Age recorded as 90+ met descriptive eligibility but was not substituted for an exact age in adjusted models. The final-record-confirmed sensitivity required the final visit itself to document the same CN or AD diagnosis.

Ordinary least squares estimated the CN-minus-AD education difference with HC3 robust t-based intervals, adjusting for age, age squared, sex, study, continuous PRS and PC1–5. Current cognition was not an adjustment variable in this phenotype comparison. Required complete covariates also included the applicable measurement lag for the PET-restricted comparisons. Retained studies required at least five participants per group, with at least twenty per group overall, residual degrees of freedom of at least twenty and maximum leverage below 0.99. Six studies met the primary adjusted-analysis criteria: ACT, ADNI, Knight Alzheimer’s Disease Research Center (Knight ADRC), NACC, the National Institute on Aging Alzheimer’s Disease Family Based Study (NIA-AD FBS) and ROS. All participants belonged to the verified post-KING genotype roster. No additional kinship model was fitted.

Top-coded age accounted for 28 CN and 74 AD participants without an exact age for adjustment. These participants are already included among those with missing required covariates in the selection counts below. The primary and sensitivity samples overlap within the same genotyped population.

| Comparison | Group | Eligible | Education observed | Complete covariates | Adjusted |
|---|---|---|---|---|---|
| Primary | CN | 411 | 410 | 382 | 293 |
| Primary | AD | 751 | 747 | 673 | 649 |
| Final-record confirmed | CN | 340 | 339 | 311 | 270 |
| Final-record confirmed | AD | 734 | 730 | 660 | 636 |

#### PRISM-AD functions and training

The network architecture and training workflow are shown in main-text Fig. 1a–c. The predicted logit is the sum of an intercept, individual clinical functions, selected pair functions, a PRS term with a non-negative coefficient and the optional history contribution. The residual neural branch was disabled. Single-feature neural functions use one hidden layer with 16 sigmoid linear units and a linear skip term and are zero-anchored at the training-transformed reference $x_j=0$. Pair networks use tanh, width 8, with pure joint contribution $f(x,y)-f(x,0)-f(0,y)+f(0,0)$. The PRS coefficient is $softplus(\theta)$.

The eight threshold bases are

$$B_{jk}(x)=clip((x-l_{jk})/max(u_{jk}-l_{jk},10^{-5}),0,1),\sum_k c_{jk}[B_{jk}(x)-B_{jk}(0)].$$

Knots are training-data quantiles. Categorical, missingness and insufficient-distinct-value bases are inactive. Threshold additions saturate outside the outer knots. The neural and linear feature terms are not clipped.

Training minimizes mean binary cross-entropy on the shared resample plus $10^{-4}P$, without applying IPCW again. $P$ sums pair-output group norms and, for active threshold coefficients, their mean square and mean squared adjacent-coefficient differences. AdamW weight decay $10^{-4}$ is separate. AdamW used learning rate 0.001, batch size 256, gradient clipping at 5 and seed 42. Training ran for 60 fixed epochs, with the final epoch retained without validation-based selection.

Each prior-assessment token contains the seven measurement fields, the current-minus-past difference for the same domain and an indicator of missing current cognition. A Gaussian error linear unit numeric projection and domain embedding feed the one-layer, width-32, four-head Transformer with feed-forward width 64 and dropout 0.1. Attention is normalized within each domain. Each domain has fixed weight $1/4$, without renormalizing absent domains. For the event-level output $v_e$ and within-domain attention $a_e$,

$$T=g(s)\frac{1}{4}\sum_{d=1}^{4}\sum_{e\in d} a_e 0.75\tanh(v_e),g(s)=1+0.5\tanh(\alpha s+b_s).$$

Here $s$ is the current mean cognitive score after training-data transformation, and $b_s$ is the severity-gate intercept. Missing observations are masked, and the history contribution is exactly zero when no earlier assessment is available. Its absolute bound is 0.75 before gating and 1.125 after gating. These bounds constrain the model and are not measured clinical effects. Recorded fitting software includes Python 3.11.15, Torch 2.6.0+cu124, scikit-learn 1.9.0 and TabPFN 8.2.0. The rendering environment is separate.

#### Training and validation loss

We reconstructed optimization trajectories for the 24 PRISM-AD fits with PRS used in the primary comparisons using the same participant partitions, preprocessing, training resamples, random seeds and fixed 60-epoch schedule. These validation folds came from the original development population and did not constitute external validation. At the end of each epoch, training loss was evaluated on the original IPCW resample, and validation loss was evaluated across all eligible held-out visits with known outcomes. Validation loss was normalized by the sum of IPCW weights, using the corresponding training-fold censoring estimates and cap of 10. Both losses were binary cross-entropy evaluated with dropout disabled and without the regularization penalty. These visit-level diagnostics differ from the primary evaluation at each participant's first eligible visit. A replay was accepted only when every recorded training loss, final model parameter and held-out prediction matched the corresponding corrected-PRS fit exactly. The supplementary monitoring did not alter the training schedule or select a checkpoint.

##### Model development and reconstruction

Earlier project analyses had already made results from folds 1–3 available before the corrected-PRS reconstruction. No prospective record established that architecture and feature exploration, the inherited score definition and later contribution-analysis choices were completed before these folds were inspected. The corrected-PRS rerun retained the same prediction-model settings, 60-epoch schedule, clinical features, folds and training resamples after that earlier development. Corrections to score construction used the GWAS and LD inputs without selecting a new PRS against progression performance. The reconstruction therefore remains part of internal model development and evaluation, rather than an untouched test.

##### Censoring, performance summaries and multiplicity

Reverse Kaplan–Meier estimation used all eligible training visits. Event code 0 denoted observation censoring, while target events and direct-AD competing events were treated as non-censoring observations. Known events by the horizon use $G(t-)^{-1}$. Horizon-observed non-events use $G(h-)^{-1}$. Unknown horizon outcomes receive zero classification/evaluation weight. Study/age fallback rules and the weight cap are specified in the main Methods.

Calibration analyses distinguished the benchmark models across all eligible visits from matched genetic-input comparisons at first eligible visits. Both used fixed 0.1-wide risk bins to summarize censoring-weighted known outcomes. Binning did not fit a recalibration model. The analysis records retained normalized and non-normalized IPCW Brier scores, weight sums, log loss and unweighted metrics. Supplementary Table 3 reports mean-risk agreement and first-visit calibration bins. Mean agreement alone does not establish calibration across the risk range, and sparse bins limit the assessment.

The complete set of contribution comparisons contains 128 tests: 112 estimable and 16 constant contributions without inference. Unavailable tests retain $p=1$ for BH correction. Representation and leverage criteria establish estimability, not covariate overlap. Ordinary stars mean BH $q<0.05$ (*), $q<0.01$ (**) and $q<0.001$ (***): BH $q$ from the 28 first-visit AP tests in Fig. 2b; the eight-test PRS-increment family in Fig. 2c,d; the four-test PRS-quintile family in Fig. 3; the 128-comparison family in Figs. 4a,b; the twelve-test shared-C family in Fig. 5d–g; and the four-comparison education family in Fig. 5j. The symbol (*) indicates nominal two-sided $P<0.05$ with $q\geq 0.05$. In inferential figure panels, unmarked tested estimates have $P\geq 0.05$. Descriptive panels without tests receive no significance markers. Distributions shown without a corresponding test and fitted main-function panels carry no inferential stars. The additional calibration-slope and marginal contribution test family is specified below.

The exploratory PRS-increment extension used only saved predictions from PRISM-AD using current assessments, with and without PRS, at each participant's first eligible visit. It tested all eight contrasts from two tasks, two horizons and two metrics (AUROC and AP), rather than selecting contrasts from the earlier intervals. Participants were sampled with replacement within validation fold, retaining both predictions and the original IPCW. For each metric difference $\Delta$, the two-sided null-centered bootstrap value was

$P = [1 + \sum_{b=1}^{9999} I(|\Delta_b^* - \Delta| \geq |\Delta|)]/10000$ (seed 42). BH correction used all eight P values. The first 500 draws reproduced the retained percentile intervals exactly. Those intervals remain nominal and are not inversions of the tests. The five-year CN-to-MCI and three-year MCI-to-AD AUROC increments reached nominal significance ($P = 0.0152$ and $0.0362$, with $q = 0.1216$ and $0.1448$), marked (*) in Fig. 2c. No PRS-increment test met $q < 0.05$, and no AP increment reached nominal $P < 0.05$. This family does not test other models, Brier scores or between-stage differences. Supplementary Table 4 reports every added test, including nonsignificant results. The PRS-increment tests used fixed predictions and censoring weights without refitting either model. Their inference is conditional on those fitted quantities and development choices, and does not capture uncertainty from overlapping training sets or adaptive development.

##### Evaluation of fitted cognitive functions

We evaluated the fitted functions from all twelve CN-to-MCI PRISM-AD models and verified that they reproduced the validation predictions. These included models using current assessments and models incorporating prior assessments, each fitted across three folds and two horizons. Main-text function plots and cross-fold summaries describe fits using current assessments. Supplementary Table 6 additionally reports models incorporating prior assessments. For each current variable, functions were evaluated on the original measurement scale over the intersection of the three training folds' 10th–90th percentile ranges. Continuous variables were evaluated at 41 evenly spaced points, whereas sex and observed-domain count used supported integer values. Functions were centered at a common reference value on the original measurement scale. High-minus-low contrasts summarized main-function changes over the evaluated range, and pairwise Spearman correlations summarized rank agreement between functions at the same grid points. Both summaries excluded interactions. Rank agreement did not measure curve shape or magnitude. Overlapping training sets precluded independent replication.

##### Exploratory overall contribution profiles

The overall display in Fig. 3 used one first-eligible CN prediction per participant from the three completed validation folds and saved three-year CN-to-MCI models using current assessments. All 908 first-eligible validation participants were considered. Of these, 906 were matched to their initial CN assessment by participant, study, visit and assessment age. For the other two participants, only one cognitive domain had been measured at the first documented CN assessment. Their prediction visit was a later eligible CN assessment with three measured domains. They remained valid prediction participants but did not satisfy the profile analysis's exact initial-visit linkage. Among the 906 linked records, nine had an undated first AD record or nonpositive elapsed time to AD. Another four had AD documented after prediction and subsequently returned to CN (two participants) or MCI (two) at the last diagnostic assessment. In this exclusion, prior AD meant AD preceding the last reverted diagnosis. It did not mean AD preceding prediction. None of the four had MCI or AD before prediction. They remained in the 908-person prediction evaluation, with direct AD treated as a competing event where applicable, and were excluded only from retrospective groups requiring no recorded AD or a last AD diagnosis. These exclusions left 893 participants: 701 with normal cognition at their last diagnostic assessment, 97 with MCI at that assessment and 95 later diagnosed with AD. The CN and MCI groups had no recorded AD. Unlike the targeted 688-person comparison, this descriptive sample imposed no additional three-year non-AD follow-up requirement. Groups used the complete recorded clinical course and were never predictors.

Applying the saved models reproduced the recorded probabilities, logits and contributions exactly. For all 688 participants shared with the targeted application, the cognitive-test component, PRS term, logit and predicted probability matched the corrected-release targeted-application records exactly. No parameters were fitted or selected. Shared raw-PRS quintile boundaries were defined across all 893 participants and applied within every clinical-course group and validation fold. Curves connect unweighted bin means. Bin counts were retained in the private analysis records. Hollow markers identify cells with fewer than five participants. Separately fitted folds retain their own contribution scales. The curves do not represent independent replication. Marginal displays in Fig. 3e,f report participant medians and interquartile ranges separately within each fitted fold and later clinical course. All individual signed C and G values are shown, with horizontal swarm packing and no pooling of fold-specific contribution scales. Interquartile ranges describe participant spread. The exploratory pairwise clinical-course tests are specified in the post hoc calibration and contribution section below.

To assess cognitive-component differences across the displayed PRS groups, we fitted $CognitiveComponent \sim PRSQuintile + ValidationFold$ separately for the full sample and each clinical-course subgroup. Quintile and fold were categorical fixed effects. Each participant contributed one observation. The five bins and three validation models were not treated as replicate observations. An HC3 heteroskedasticity-robust joint F test assessed the four quintile coefficients, with residual degrees of freedom $n-7$. BH correction covered all four omnibus tests (Supplementary Table 4). Adjusted q values ranged from 0.4539 to 0.6816. No pairwise PRS-quintile comparisons or alternative trend tests were selected afterward. No genetic-term test was performed because its increase with PRS follows from model specification. These exploratory tests condition on the fitted models and adjust for fold only. Retrospective clinical-course grouping, sparse cells, model-specific contribution scales and overlapping training sets limit interpretation. A nonsignificant omnibus result does not establish equality or exclude clinically relevant associations. The plotted bin means remain unadjusted.

**Targeted high-PRS contribution comparisons**

To characterize preserved cognition at high genetic risk, we selected CN participants in the previously defined high-PRS interval given in the main Methods. The same predefined raw interval was applied to corrected scores without selecting a new top-quintile cutoff. Participants in this group and the two comparison groups were linked to validation predictions by participant, study, visit and cognitive-assessment age. All groups began as CN without prior recorded MCI or AD. The two non-AD groups required no recorded AD, a last diagnostic assessment showing normal cognition and at least three years of follow-up with known cognitive status. The CN-later-AD clinical-course group required a subsequent dated AD event and a final AD record. A fixed sensitivity analysis required every subsequent diagnostic assessment in the non-AD groups to remain CN. Clinical-course groups were defined retrospectively to interpret the forecasts and were never used as predictors. The analysis did not provide prospective validation of resilience. Because the CN-later-AD comparator was not restricted to the high-PRS interval, these group comparisons cannot establish protection at matched inherited susceptibility.

Fitted terms were assigned to non-overlapping groups whose sum was checked against each predicted logit. C was the contribution assigned to current cognitive performance on the fitted risk scale, distinct from a raw cognitive score or a separately fitted prediction based on cognition alone. The education contribution contained the education main function alone. Missingness indicators and all pairwise interactions, including education with the mean cognitive score, remained separate.

For the targeted group comparisons, eight summaries comprised the education and cognition main functions, main functions of prior-assessment summaries, missingness indicators, pairwise interactions, PRS, the Transformer contribution from prior visits and total predicted logit (Supplementary Table 1). The total logit combined all contributions to form the forecast and was not an additional component. Reconstruction also included the intercept, age, sex, study and domain coverage. Contributions were log-odds terms, not risk percentages.

These contribution summaries were compared across two group contrasts, two model versions, two horizons and two group scopes, yielding 128 planned tests. The full family comprised 128 comparisons, including 112 estimable comparisons and 16 constant-contribution comparisons without inference. Supplementary Table 8 reports the selected main-text contrasts from this family. Analyses used ordinary least squares with heteroskedasticity-consistent (HC3) standard errors and t-based intervals, adjusting for age, age squared, sex, study, PC1–5 and prediction fold. Adjusted comparisons required at least five participants in each group within each study and at least twenty in each group overall. Models also required at least twenty residual degrees of freedom and maximum leverage below 0.99. Constant contribution summaries and comparisons failing these criteria were reported as non-estimable. Benjamini–Hochberg correction covered all planned tests, assigning $p = 1$ to unavailable tests, while the plotted confidence intervals remained nominal. The primary adjusted comparisons included 64 high-PRS last-CN participants, 495 other CN and 63 CN-later-AD participants. Unadjusted distributions included all matched participants, whereas adjusted contrasts used the restricted sample with adequate representation of both groups within studies. For PET-defined subgroups, we assessed data availability only, without testing differences in model contributions.

##### Genetic and cognitive contributions across later clinical courses

Main-text Fig. 5d–g uses the same 893-person descriptive sample as Fig. 3: all participants were CN at their first eligible held-out prediction visit, with normal cognition at the last diagnostic assessment (n = 701), MCI without recorded AD (n = 97), or a dated subsequent AD diagnosis (n = 95). Unlike the targeted 688-person comparison, no additional three-year non-AD follow-up requirement is imposed. Participants were included without conditioning on total predicted risk. Groups were defined by subsequent diagnoses and were not used as predictors.

Ten shared deciles of the fitted cognitive component C are defined in all 893 participants and applied to every clinical-course group. Boundaries are not recalculated within groups. The deciles group fitted cognitive contributions and do not provide raw cognitive-test strata or exact matching on C. Within group and bin, positive factors are geometric means: $\exp(\overline{G})$ and $\exp(\overline{C})$. C contains the five cognitive main functions, excluding the intercept, missingness, interactions and other noncognitive terms. Higher factors increase predicted odds with remaining terms fixed. At the individual level, $\exp(C)$, $\exp(G)$ and $\exp(R)$ multiply to give the complete predicted odds. The displayed geometric means do not represent shares of probability.

Every participant contributes once. Private analysis records contain bin counts, boundaries and signed means. Overall bins contain 89–90 participants. CN bins contain 57–81, MCI bins 5–17 and AD bins 4–22. Open circles identify cells with fewer than five participants. No monotone fit, absolute values or percentage normalization is used. The increase in C follows the definition of its deciles. The genetic factor is higher in later AD than CN in nine of ten common bins. Bars remain unadjusted descriptive summaries.

**Exploratory AD–CN contrasts within cognitive-risk strata**

After examining the profiles, we specified the exploratory comparisons before calculating their P values. The comparison included the 95 later-AD and 701 participants with normal cognition at their last diagnostic assessment (n = 796). MCI participants contributed to the displayed profiles and common boundaries, but were excluded from this AD–CN contrast. We analyzed signed G rather than exponentiated values to avoid testing skewed factors or treating ten bin means as independent observations.

The pooled ordinary least-squares model regressed G on an AD indicator, nine C-decile indicators, centered continuous C and validation-fold indicators. The AD coefficient estimated the adjusted AD-minus-CN difference in G. Within each decile, a separate model included the AD indicator, continuous C and fold. An expanded pooled sensitivity added age, age squared, sex, study and PC1–5, using complete covariates and retaining studies with at least five participants in each group. All 796 participants met this expanded support requirement. Exactly redundant nuisance columns were removed by a deterministic rule while requiring the AD contrast to remain identifiable.

Each comparison required at least five AD and five CN participants, residual degrees of freedom of at least twenty, maximum leverage below 0.99 and nonzero residual variation. HC3 heteroskedasticity-consistent standard errors, two-sided residual-df t tests and nominal 95% t intervals were used. AD deciles 1 and 2 were non-estimable under the group-size rule. Their numerical effects and inferential P values are not reported. One BH correction family included all twelve planned slots: the pooled comparison, ten within-decile comparisons and the expanded sensitivity. Non-estimable slots entered correction as P = 1. Stars above AD genetic bars compare AD with CN within the corresponding C decile, not PRS with cognition. Ordinary *, ** and *** denote BH q below 0.05, 0.01 and 0.001. The symbol (*) denotes nominal P below 0.05 with q at least 0.05.

Exponentiating a coefficient gives a ratio of modeled genetic odds factors, not an odds ratio for developing AD. The corrected-release estimates and all twelve planned test slots are reported in Supplementary Table 4C. Deciles 8 and 9 met BH q below 0.01, while decile 6 met nominal P below 0.05 only.

The models are post hoc group comparisons, conditional on saved fitted contributions. Their intervals exclude prediction-model refitting and adaptive-development uncertainty. Fold adjustment accounts for mean shifts but does not establish a common biological scale across separately fitted models with overlapping training sets. Adjusting linearly for C within broad bins does not make participants exactly matched on C. Conditioning on a model component, residual confounding, unequal follow-up and diagnostic ascertainment limit interpretation. The tests do not establish genetic causation, compensation, clinical AD risk ratios, or prevention benefit. Fig. 3 retains its separate PRS-quintile tests.

**Within-fold and raw-PRS sensitivity checks**

We specified four additional post hoc comparisons before calculating their estimates, retaining the original twelve-test family and all prediction models. Shared C-decile boundaries were retained from the complete 893-person profile sample. Each fold-specific model compared signed G between later AD and last CN, adjusting for continuous C and those common decile indicators within that fold.

Within a fitted fold, G is a non-negative coefficient times its training-standardized PRS. The empirical G–raw-PRS relationship was affine to numerical precision. Fold indicators in a pooled regression can absorb mean shifts but do not align the fold-specific coefficient scales. We therefore also compared raw PRS on a common scale, standardized using the mean and population standard deviation (ddof = 0) in the complete corrected post-QC genotype roster. This pooled model adjusted for continuous C, C-decile indicators and fold.

All four comparisons used ordinary least squares with HC3 standard errors, residual-df t intervals and two-sided tests. The same group-size, residual-df, leverage and identifiability criteria were applied. A separate BH family included the three within-fold G comparisons and the pooled raw-PRS comparison, with unavailable slots assigned P = 1. All four were estimable. Exponentiated G coefficients are model-specific factor ratios. The raw-PRS coefficient is a difference in population SD and is not exponentiated or interpreted as clinical AD odds. The results appear in Supplementary Table 4D.

These checks assess scale dependence without providing independent replication: training sets overlap, C remains a fitted quantity, and the same retrospective clinical-course and follow-up selection applies. The additional follow-up sensitivity analyses below examine a different outcome definition and use all eligible CN prediction participants.

**Sensitivity to observation time and a common AD outcome window**

The two exploratory comparisons were specified before estimating effects, including a clarification of censoring at the inclusive horizon. Both analyses used all 908 first-eligible CN validation participants without selecting their later clinical course. Frozen three-year CN-to-first-MCI checkpoints reproduced C and raw PRS for this complete sample. The original 893 profile records and their contribution values were unchanged. The original profile sample supplied 95 first AD events. Four post-prediction AD/reversion cases supplied four more, while two original-index nonmatches and nine records with an undated first AD contributed censored observations.

The endpoint was the first subsequently recorded AD diagnosis, allowing intermediate MCI and retaining a first AD event despite later reversion. Records were ordered by visit. The first AD record defined event time when its date was known. If that first record was undated or top-coded, observation ended at the preceding dated non-AD assessment, without substituting a later diagnosis. If no AD was recorded, follow-up ended at the last exact assessment with an explicit non-AD diagnosis. All 908 had eligible endpoints and complete adjustment variables, with 99 AD events and 809 right-censored observations. This endpoint differs from first MCI and does not reuse its event labels or censoring weights.

Raw PRS used the complete corrected genotype roster mean and population SD. The planned primary sensitivity was a Cox working model fitted in R 4.5.3 with survival 3.8.9, using fold-specific baseline hazards, Efron ties and the package's default near-tie time correction. Predictors were raw PRS, three separately centered fold-specific C terms, centered age divided by ten, its square, sex and study indicators. Robust sandwich standard errors supplied nominal normal intervals and two-sided Wald tests. Rank-time Schoenfeld diagnostics evaluated individual terms and the global proportional-hazards assumption. The global diagnostic indicated nonproportional hazards (P < 0.001), chiefly involving C in folds 1 and 2. The raw-PRS diagnostic gave P = 0.0634, which does not establish proportional hazards. The specified estimate is therefore reported as a working-model association, with no alternative model selected after the diagnostic.

The planned secondary sensitivity compared raw PRS between participants with and without recorded AD within five years. AD at or before five years counted as an event. An explicitly observed absence of AD through five years counted as no AD, including intermediate MCI and AD recorded later than five years. Earlier censoring left the five-year outcome unknown. This yielded 31 AD and 598 no-AD observations, with 279 unknown outcomes. New AD-specific reverse-Kaplan–Meier censoring models used study and the original age bands, with study fallback for strata smaller than 50. Weights used left-limit censoring survival at the event time for AD and at five years for known no-AD observations. No event or censoring time equaled exactly five years. The cap of ten clipped no weights. Effective weighted sample size was 563.1.

Weighted least squares treated raw PRS in population SD as the outcome and five-year AD status as the contrast. Adjustment included the same C, age, sex and study terms plus fold indicators and an intercept. HC3 residual-df t inference used 616 degrees of freedom. Maximum weighted leverage was 0.255. The coefficient is an adjusted score difference, not an odds ratio or absolute AD risk. A separate two-test BH family covered the raw-PRS Cox and five-year contrasts, without changing the original twelve-test or four-test families (Supplementary Table 4E). All specified results and diagnostics are reported. No PRISM parameters were refitted.

These analyses address recorded diagnostic follow-up under conditional censoring assumptions. Death and non-AD dementia were not separately modeled, so competing-risk cumulative AD incidence was not estimated. Diagnostic attendance, interval uncertainty, fitted C, estimated weights and earlier adaptive development remain sources of uncertainty. Neither analysis provides independent validation, a biological-onset estimate or a diagnostic lead time.

##### Descriptive component correlations

Spearman correlations between the individual fitted PRS term G and current cognitive main-function component C were calculated in all 893 participants and separately within the CN, MCI and AD follow-up groups, before decile aggregation. The pooled coefficients were 0.056, 0.031, -0.078 and 0.007, respectively. Identical coefficients were obtained after exponentiating G and C because ranks were unchanged. The figure rounds them to two decimals. No P values or significance markers accompany these descriptive correlations. Correlations within individual validation folds were also retained in the private analysis records. CN coefficients ranged from -0.035 to 0.047. MCI and AD directions differed across folds, limiting a claim of a stable inverse relation. Pooled fitted terms and retrospective clinical-course groups do not establish biological independence or genetic compensation.

##### Comparator settings and inference reproducibility

Logistic regression used inverse regularization strength 1 and at most 2,000 iterations. HGB used learning rate 0.05, 250 boosting iterations, at most 15 leaves, at least 20 observations per leaf, L2 regularization 1 and no early stopping. TabPFN package version 8.2.0 used the fixed TabPFN-3 classifier with eight estimators, preprocessing-cache inference and automatic precision. Automatic estimator scaling was disabled. TabPFN seeds were 42 plus validation-fold number. All models ran locally. An inference-only correction made randomized GPU preprocessing reproducible by resetting and restoring the Torch random state. The correction left labels, training rows and model parameters unchanged, and repeated predictions were identical, with no selection by predictive performance.

##### PRS effective information, LD normalization and reconstruction checks

The GWAS summary statistics under accession GCST90012877 combine clinical and family-history meta-analysis effects without variant-level effective sample sizes. Following the LDpred2 guide,[S1] effective information was estimated on the supplied log-odds scale as

$$\widehat{N}_{eff} = Q_{0.999}\left(8/SE^2\right),$$

using R type-7 quantiles across the retained variants. The resulting value was 90,080.7768, used without rounding in LDpred2-auto. Corresponding estimates from the complete supplied GWAS and common-variant allele-frequency/SE checks were approximately 89,960 and 88,773. These checks support the scale choice but do not establish a literal combined participant count or remove uncertainty from proxy phenotypes. The discovery publication already transformed proxy effects before meta-analysis, so no additional proxy multiplier was applied to the supplied combined effects.

A fresh allele match reproduced 618,471 matched variants. Removal of one monomorphic variant left 618,470 variants represented in both the LD-score sums and their normalization. LD scores were independently recomputed from the cached target-genotype matrix and matched exactly. LDSC normalization therefore used $M = 618,470$, matching the variants in the LD-score sums instead of the complete HapMap3+ map.

The reconstruction retained the original LDpred2-auto seed, 30 starting chains, 500 burn-in iterations and 500 sampling iterations. The original joint h2/p chain filter retained 29 chains for the corrected LD normalization and effective-information estimate. Independent correlation-range diagnostics flagged no chains. Weights were averaged over the retained chains before scoring all 26,390 post-QC participants.

An isolated historical replay reproduced the earlier PRS to numerical precision. Relative to that historical score, the corrected score had Pearson correlation 0.999949 and Spearman correlation 0.999940, with standardized RMSE 0.01008 SD. Applying the same prespecified raw high-PRS interval changed membership for 100 participants in the full genotyped roster. The high correlation alone was insufficient to establish that downstream results were unchanged. Clinical and genotype IDs were aligned one to one, all non-PRS prediction inputs and outcomes were verified unchanged, and all prognostic models, comparison families, contribution analyses, education and PET summaries were recalculated from the corrected scores. The unchanged raw interval was retained without optimizing its boundaries to recover historical groups. These checks assess implementation consistency and reconstruction, while external clinical validity remains untested.

##### Participant characteristics and diagnostic follow-up

Supplementary Table 11 describes each clinical task at the first eligible visit, separating folds 1–9 from the evaluated folds 1–3. Missingness is reported before imputation, and overlapping training rounds are not treated as independent populations. Supplementary Table 7 uses the frozen records of the 893-person contribution-profile sample. Dated follow-up excludes ages recorded as 90 years or older and does not impose a common AD outcome window.

Supplementary Table 3 reports calibration bins from the corrected-PRS first-visit predictions and their IPCW summaries. Bin means and event fractions are pooled across the three validation folds by the sum of their weights. Binning did not recalibrate forecasts or refit prognostic models, and the diagnostic tests below used individual predictions. Means and event fractions are not displayed for bins with fewer than five known-outcome participants.

#### Post hoc calibration and contribution comparisons

Calibration slopes were estimated from individual first-eligible held-out forecasts among participants with known horizon outcomes and positive IPCW. The plotted bin means were not used to fit the slopes. For each task and horizon, IPCW-weighted logistic regression estimated a free intercept and a slope on the logit of the predicted probability. The reference slope was 1.[S2] Probabilities were bounded at $10^{-8}$ and $1-10^{-8}$ for the diagnostic logit, but no observed probability required clipping. Empirical participant-score sandwich covariance used HC1 correction $n/(n-2)$. Nominal normal 95% intervals and two-sided Wald tests were conditional on saved forecasts and censoring weights, without propagating prognostic-model fitting, censoring estimation or earlier adaptive-development uncertainty. Diagnostic coefficients were not applied to alter forecasts. A slope compatible with 1 does not establish calibration across the risk range.

Each of the 893 participants contributed one signed value per component. Swarm packing changed only horizontal display positions. The six pairwise clinical-course comparisons used van Elteren fold-stratified Wilcoxon statistics.[S3] Within each fold, midranks and tie-corrected null variance were combined with weights $1/(n_A+n_B+1)$ for the compared pair. Two-sided normal inference avoided pooling native contribution scales across fitted models. The rank-biserial effect was $r=2P_S-1$, where $P_S$ was the sum of weighted within-fold concordant-pair counts (ties counted as one half), divided by the corresponding weighted total pair count. Positive effects indicate larger values in the second group. The tests compare rank distributions and do not isolate median shifts or estimate clinical risk differences or causal effects. They do not adjust for C, age, study or unequal diagnostic follow-up.

The four slope tests and six marginal contrasts formed one additional ten-slot BH family for these post hoc analyses. Unsupported tests retained a P = 1 slot, although all ten tests were estimable. Ordinary stars use q thresholds of 0.05, 0.01 and 0.001. (*) denotes nominal P below 0.05 with q at or above 0.05, and n.s. denotes nominal P at or above 0.05. Earlier families were unchanged. This correction does not account for earlier project exploration, and none of these analyses provides external validation.

### Supplementary Table 1 | PRISM-AD contribution definitions and feature pairs

#### Contribution summaries used in the group comparisons

All contributions are expressed on the fitted log-odds scale. The eight summaries were evaluated across two group contrasts, two model versions, two horizons and two group scopes, yielding a family of 128 tests.

| Contribution summary | Included terms | Excluded terms and interpretation |
|---|---|---|
| Education main | The individual-feature function for years of education. | Does not include the education missingness indicator or education-by-mean-cognitive-score interaction. |
| Cognition main | Sum of individual-feature functions for memory, executive function, language, visuospatial performance and the mean of available domain scores. | Does not include missingness indicators, number of observed domains, interactions or prior cognitive summaries. |
| Past-summary main | Sum of individual-feature functions for the 21 strictly prior cognitive summaries, in models incorporating prior assessments. | Excludes their missingness indicators, interactions and the Transformer contribution from prior visits. Absent in models using current assessments. |
| Missingness main | Sum of individual-feature functions for all retained static-input missingness indicators, including cognitive, education and history-summary indicators where present. | Kept separate from the corresponding measured-variable functions. Does not include sequence masks. |
| Pairwise interactions | Sum of all retained clinical pair functions listed on the next page. | No interaction is assigned to either constituent main term. No phenotype-by-PRS pair is included. |
| PRS | Training-standardized PRS multiplied by the fitted non-negative genetic coefficient. | A separate genetic term, not part of cognition or clinical interactions. |
| Prior-visit sequence | The complete signed sequence contribution after within-domain attention, fixed domain weights and current-severity gating. | Separate from past-summary main functions. Exactly zero when no prior sequence is available and in models using current assessments. |
| Total logit | The complete predicted log odds, including the intercept and every retained additive term. | The full prediction already includes the component terms; adding it to them would count their contributions twice. |

“Cognition” and “Education” in Figs. 4a,b denote the corresponding main-function contributions. Cognitive and education measurements can also influence predictions through other model pathways. In particular, changing a cognitive value can also change an interaction or the history gate without changing the definition of the cognition main contribution. The cognitive-test risk component is expressed in log-odds units. Higher values correspond to higher predicted MCI risk when the other terms are held fixed. It is distinct from a raw cognitive test score or a standalone probability.

The mean cognitive score is the arithmetic mean of available PHC domain scores, with at least two domains required for eligibility. It is not a separately harmonized or validated global instrument, and its domain composition may differ between visits. Domain counts and missingness indicators enter separately.

#### Supplementary Table 1 | Complete clinical pair list

The first five pairs are retained in both models using current assessments and models incorporating prior assessments. The remaining nine require history-summary inputs and occur only in models incorporating prior assessments. Pairs are included whenever both named inputs are present, without selection by validation performance. The reported models use this complete pair list.

| First input | Second input | Model versions |
|---|---|---|
| Age | Mean cognitive score | Current and history |
| Age | Memory | Current and history |
| Years of education | Mean cognitive score | Current and history |
| Mean cognitive score | Memory | Current and history |
| Mean cognitive score | Executive function | Current and history |
| Mean cognitive score | Mean prior rate of cognitive change | History only |
| Mean cognitive score | Prior memory rate of change | History only |
| Mean cognitive score | Prior executive-function rate of change | History only |
| Mean cognitive score | Number of prior observed domain measurements | History only |
| Memory | Most recent prior memory score | History only |
| Memory | Prior memory rate of change | History only |
| Executive function | Most recent prior executive-function score | History only |
| Executive function | Prior executive-function rate of change | History only |
| Span of prior cognitive observations | Mean prior rate of cognitive change | History only |

Prior rates are expressed per five years and calculated using strictly earlier measurements, excluding the prediction visit. The mean prior rate averages the domain-specific rates that can be estimated. The prior-observation count refers to observed domain measurements, which may include several measurements from the same visit. Mean cognitive score refers to the available-domain mean defined on the preceding page.

Each pair contribution is centered to isolate the jointly varying term: for a fitted pair function $f$, the contribution is $f(x,y)-f(x,0)-f(0,y)+f(0,0)$ on the transformed input scale. This accounting convention does not identify a biological interaction.

#### Supplementary Table 1 | Complete additive accounting

The comparison summaries include selected contributions and the total logit. Adding them would omit some model terms and count the total logit again. The prediction is reconstructed from the disjoint terms as follows:

$$\begin{aligned}\text{total logit} &= \text{intercept} + \text{age main} + \text{sex main} + \text{education main} \\ &+ \text{cognition main} + \text{domain-coverage main} + \text{study main} \\ &+ \text{past-summary main} + \text{missingness main} + \text{pairwise interactions} \\ &+ \text{clinical residual} + \text{PRS} + \text{prior-visit sequence}\,.\end{aligned}$$

| Additional accounting term | Definition |
|---|---|
| Intercept | The fitted clinical intercept. |
| Age main | Individual-feature function for age at the prediction visit. |
| Sex main | Individual-feature function for PHC sex. |
| Domain-coverage main | Individual-feature function for the number of observed cognitive domains. |
| Study main | Sum of functions for the four study indicators: ADNI, NACC, ROS and MAP. |
| Clinical residual | A reserved ledger term that is identically zero in the reported architecture because its residual network is disabled. |

Missingness assignment takes precedence over all other groups: a missingness indicator for a history summary belongs to missingness main, not past-summary main. The sequence contribution includes its own masks and severity gate. These are not allocated again to the static missingness or cognition terms. All main terms refer to the complete feature function, including its nonlinear neural term, linear term and any active piecewise-linear addition.

The disjoint terms reconstructed every held-out logit, verifying the model's numerical accounting. This check does not establish causal effects or a unique allocation among correlated predictors. The seven selected component summaries and total logit were compared statistically. The additional terms above remained in the prediction without being added to that comparison family.

#### Supplementary Table 2 | Genetic information and prediction

Models fitted with and without PRS used identical clinical inputs, participant folds and censoring-weighted training resamples. Evaluation used the first eligible held-out visit per participant. Differences are PRS-augmented minus clinical-only performance. Intervals are nominal 95% percentile intervals from 500 paired participant-bootstrap resamples within fold, conditional on fitted models and censoring weights. The intervals omit model-refitting and development uncertainty.

##### CN to MCI, 3 y

###### AUROC

| Model | Without PRS | With PRS | Difference (95% CI) |
|---|---|---|---|
| PRISM-AD | 0.7306 | 0.7332 | 0.0026 (-0.0124 to 0.0165) |
| Logistic | 0.7424 | 0.7446 | 0.0022 (-0.0158 to 0.0188) |
| HGB | 0.6928 | 0.6873 | -0.0055 (-0.0397 to 0.0269) |
| TabPFN | 0.7195 | 0.7314 | 0.0118 (-0.0124 to 0.0364) |

###### Average precision

| Model | Without PRS | With PRS | Difference (95% CI) |
|---|---|---|---|
| PRISM-AD | 0.3019 | 0.2996 | -0.0023 (-0.0281 to 0.0233) |
| Logistic | 0.2941 | 0.2951 | 0.0010 (-0.0274 to 0.0320) |
| HGB | 0.2312 | 0.2340 | 0.0028 (-0.0412 to 0.0407) |
| TabPFN | 0.2504 | 0.2593 | 0.0089 (-0.0324 to 0.0607) |

###### Hajek Brier score

| Model | Without PRS | With PRS | Difference (95% CI) |
|---|---|---|---|
| PRISM-AD | 0.0710 | 0.0708 | -0.0002 (-0.0014 to 0.0009) |
| Logistic | 0.0705 | 0.0705 | -0.0000 (-0.0013 to 0.0012) |
| HGB | 0.0758 | 0.0759 | 0.0001 (-0.0027 to 0.0027) |
| TabPFN | 0.0740 | 0.0731 | -0.0009 (-0.0035 to 0.0018) |

#### CN to MCI, 5 y

##### AUROC

| Model | Without PRS | With PRS | Difference (95% CI) |
|---|---|---|---|
| PRISM-AD | 0.7502 | 0.7708 | 0.0206 (0.0055 to 0.0359) |
| Logistic | 0.7490 | 0.7696 | 0.0206 (0.0018 to 0.0380) |
| HGB | 0.7457 | 0.7531 | 0.0073 (-0.0172 to 0.0299) |
| TabPFN | 0.7659 | 0.7645 | -0.0014 (-0.0284 to 0.0270) |

##### Average precision

| Model | Without PRS | With PRS | Difference (95% CI) |
|---|---|---|---|
| PRISM-AD | 0.4108 | 0.4094 | -0.0013 (-0.0262 to 0.0346) |
| Logistic | 0.3900 | 0.4091 | 0.0191 (-0.0099 to 0.0514) |
| HGB | 0.3820 | 0.3887 | 0.0067 (-0.0454 to 0.0548) |
| TabPFN | 0.3528 | 0.3805 | 0.0277 (-0.0184 to 0.0845) |

##### Hajek Brier score

| Model | Without PRS | With PRS | Difference (95% CI) |
|---|---|---|---|
| PRISM-AD | 0.1127 | 0.1106 | -0.0022 (-0.0050 to 0.0007) |
| Logistic | 0.1130 | 0.1106 | -0.0024 (-0.0056 to 0.0008) |
| HGB | 0.1155 | 0.1137 | -0.0019 (-0.0083 to 0.0042) |
| TabPFN | 0.1215 | 0.1190 | -0.0025 (-0.0109 to 0.0053) |

#### MCI to AD, 3 y

##### AUROC

| Model | Without PRS | With PRS | Difference (95% CI) |
|---|---|---|---|
| PRISM-AD | 0.8170 | 0.8216 | 0.0046 (0.0001 to 0.0090) |
| Logistic | 0.8171 | 0.8225 | 0.0055 (-0.0019 to 0.0122) |
| HGB | 0.7937 | 0.8034 | 0.0098 (-0.0057 to 0.0246) |
| TabPFN | 0.7939 | 0.7901 | -0.0038 (-0.0211 to 0.0137) |

##### Average precision

| Model | Without PRS | With PRS | Difference (95% CI) |
|---|---|---|---|
| PRISM-AD | 0.7089 | 0.7105 | 0.0016 (-0.0066 to 0.0085) |
| Logistic | 0.7180 | 0.7164 | -0.0016 (-0.0168 to 0.0106) |
| HGB | 0.7001 | 0.6951 | -0.0050 (-0.0319 to 0.0188) |
| TabPFN | 0.6876 | 0.6784 | -0.0092 (-0.0400 to 0.0198) |

##### Hajek Brier score

| Model | Without PRS | With PRS | Difference (95% CI) |
|---|---|---|---|
| PRISM-AD | 0.1619 | 0.1605 | -0.0014 (-0.0032 to 0.0008) |
| Logistic | 0.1617 | 0.1601 | -0.0016 (-0.0044 to 0.0018) |
| HGB | 0.1736 | 0.1732 | -0.0004 (-0.0085 to 0.0092) |
| TabPFN | 0.1739 | 0.1779 | 0.0040 (-0.0046 to 0.0130) |

#### MCI to AD, 5 y

##### AUROC

| Model | Without PRS | With PRS | Difference (95% CI) |
|---|---|---|---|
| PRISM-AD | 0.8129 | 0.8170 | 0.0041 (-0.0002 to 0.0088) |
| Logistic | 0.8050 | 0.8097 | 0.0047 (-0.0041 to 0.0148) |
| HGB | 0.7859 | 0.8009 | 0.0150 (-0.0030 to 0.0354) |
| TabPFN | 0.7802 | 0.7712 | -0.0090 (-0.0329 to 0.0144) |

##### Average precision

| Model | Without PRS | With PRS | Difference (95% CI) |
|---|---|---|---|
| PRISM-AD | 0.8134 | 0.8189 | 0.0055 (0.0003 to 0.0116) |
| Logistic | 0.8081 | 0.8146 | 0.0065 (-0.0042 to 0.0193) |
| HGB | 0.7666 | 0.7827 | 0.0161 (-0.0063 to 0.0370) |
| TabPFN | 0.7676 | 0.7377 | -0.0299 (-0.0590 to -0.0016) |

##### Hajek Brier score

| Model | Without PRS | With PRS | Difference (95% CI) |
|---|---|---|---|
| PRISM-AD | 0.1789 | 0.1770 | -0.0018 (-0.0040 to 0.0003) |
| Logistic | 0.1827 | 0.1813 | -0.0014 (-0.0060 to 0.0033) |
| HGB | 0.2020 | 0.1946 | -0.0074 (-0.0211 to 0.0046) |
| TabPFN | 0.2081 | 0.2183 | 0.0102 (-0.0056 to 0.0266) |

#### Supplementary Table 3 | Mean predicted and observed risk

Means were calculated at the first eligible visits in the same validation populations, using censoring weights and known outcomes. Agreement of means is a limited descriptive check and does not establish calibration across the risk range. Risk-bin summaries are reported separately.

| Task, horizon | Model | Mean risk, without PRS | Mean risk, with PRS | Observed fraction |
|---|---|---|---|---|
| CN to MCI, 3 y | PRISM-AD | 0.0881 | 0.0876 | 0.0862 |
| CN to MCI, 3 y | Logistic | 0.0883 | 0.0878 | 0.0862 |
| CN to MCI, 3 y | HGB | 0.0804 | 0.0779 | 0.0862 |
| CN to MCI, 3 y | TabPFN | 0.0755 | 0.0781 | 0.0862 |
| CN to MCI, 5 y | PRISM-AD | 0.1583 | 0.1599 | 0.1534 |
| CN to MCI, 5 y | Logistic | 0.1568 | 0.1583 | 0.1534 |
| CN to MCI, 5 y | HGB | 0.1489 | 0.1510 | 0.1534 |
| CN to MCI, 5 y | TabPFN | 0.1686 | 0.1782 | 0.1534 |
| MCI to AD, 3 y | PRISM-AD | 0.3378 | 0.3381 | 0.3519 |
| MCI to AD, 3 y | Logistic | 0.3370 | 0.3373 | 0.3519 |
| MCI to AD, 3 y | HGB | 0.3383 | 0.3466 | 0.3519 |
| MCI to AD, 3 y | TabPFN | 0.3644 | 0.3785 | 0.3519 |
| MCI to AD, 5 y | PRISM-AD | 0.5074 | 0.5077 | 0.4788 |
| MCI to AD, 5 y | Logistic | 0.5038 | 0.5042 | 0.4788 |
| MCI to AD, 5 y | HGB | 0.5149 | 0.5237 | 0.4788 |
| MCI to AD, 5 y | TabPFN | 0.5576 | 0.5771 | 0.4788 |

#### Supplementary Table 3 | First-visit calibration-bin summaries

These are descriptive calibration summaries for PRISM-AD using current assessments and PRS, evaluated at the same first eligible held-out visits as the primary discrimination analysis. Forecasts were not recalibrated. Only known horizon outcomes contribute. Within each fixed risk bin, fold summaries are pooled by the sum of their original IPCW weights. Event fractions therefore reflect censoring weighting and are not crude proportions.

The table reports the number of known-outcome participants, IPCW-weighted mean prediction and IPCW-weighted recorded event fraction. Empty bins are retained. Means and event fractions are not displayed for bins with fewer than five known-outcome participants. Sparse upper-risk bins therefore cannot establish calibration.

##### CN-to-first-MCI, 3 years

| Predicted-risk bin | Known outcomes, n | Mean predicted risk | Observed event fraction |
|---|---|---|---|
| 0.0–0.1 | 579 | 0.041 | 0.049 |
| 0.1–0.2 | 133 | 0.137 | 0.122 |
| 0.2–0.3 | 53 | 0.245 | 0.207 |
| 0.3–0.4 | 20 | 0.339 | 0.173 |
| 0.4–0.5 | 6 | 0.444 | 0.795 |
| 0.5–0.6 | 7 | 0.543 | 0.557 |
| 0.6–0.7 | 3 | — | — |
| 0.7–0.8 | 0 | — | — |
| 0.8–0.9 | 0 | — | — |
| 0.9–1.0 | 0 | — | — |

##### CN-to-first-MCI, 5 years

| Predicted-risk bin | Known outcomes, n | Mean predicted risk | Observed event fraction |
|---|---|---|---|
| 0.0–0.1 | 328 | 0.050 | 0.048 |
| 0.1–0.2 | 163 | 0.142 | 0.158 |
| 0.2–0.3 | 65 | 0.240 | 0.218 |
| 0.3–0.4 | 36 | 0.353 | 0.335 |
| 0.4–0.5 | 24 | 0.449 | 0.353 |
| 0.5–0.6 | 20 | 0.552 | 0.544 |
| 0.6–0.7 | 10 | 0.644 | 0.650 |
| 0.7–0.8 | 9 | 0.757 | 0.736 |
| 0.8–0.9 | 3 | — | — |
| 0.9–1.0 | 0 | — | — |

#### MCI-to-AD, 3 years

| Predicted-risk bin | Known outcomes, n | Mean predicted risk | Observed event fraction |
|---|---|---|---|
| 0.0–0.1 | 130 | 0.049 | 0.075 |
| 0.1–0.2 | 94 | 0.149 | 0.146 |
| 0.2–0.3 | 67 | 0.246 | 0.278 |
| 0.3–0.4 | 57 | 0.354 | 0.355 |
| 0.4–0.5 | 53 | 0.445 | 0.517 |
| 0.5–0.6 | 41 | 0.552 | 0.516 |
| 0.6–0.7 | 45 | 0.642 | 0.627 |
| 0.7–0.8 | 36 | 0.747 | 0.767 |
| 0.8–0.9 | 34 | 0.848 | 0.893 |
| 0.9–1.0 | 18 | 0.939 | 0.862 |

#### MCI-to-AD, 5 years

| Predicted-risk bin | Known outcomes, n | Mean predicted risk | Observed event fraction |
|---|---|---|---|
| 0.0–0.1 | 40 | 0.048 | 0.075 |
| 0.1–0.2 | 44 | 0.142 | 0.253 |
| 0.2–0.3 | 43 | 0.248 | 0.178 |
| 0.3–0.4 | 30 | 0.351 | 0.367 |
| 0.4–0.5 | 46 | 0.439 | 0.431 |
| 0.5–0.6 | 43 | 0.555 | 0.427 |
| 0.6–0.7 | 46 | 0.655 | 0.523 |
| 0.7–0.8 | 50 | 0.756 | 0.566 |
| 0.8–0.9 | 62 | 0.845 | 0.927 |
| 0.9–1.0 | 82 | 0.947 | 0.930 |

#### Calibration slope diagnostics

Calibration slopes were estimated by weighted logistic regression with a free intercept, using individual first-eligible held-out forecasts among participants with known horizon outcomes and positive IPCW. The reference slope is 1. Empirical participant-score sandwich covariance used HC1 finite-sample correction, with nominal normal 95% intervals and P values testing slope = 1.

| Clinical transition, horizon | Known n | Slope (nominal 95% CI) | P | BH q | Annotation |
|---|---|---|---|---|---|
| CN to first MCI, 3 y | 801 | 0.826 (0.582 to 1.069) | 0.1599 | 0.2233 | n.s. |
| CN to first MCI, 5 y | 658 | 0.868 (0.667 to 1.070) | 0.2010 | 0.2233 | n.s. |
| MCI to AD, 3 y | 575 | 0.894 (0.714 to 1.073) | 0.2466 | 0.2466 | n.s. |
| MCI to AD, 5 y | 486 | 0.844 (0.680 to 1.009) | 0.0635 | 0.1059 | n.s. |

These four slope tests share one new ten-test BH family with the six marginal contrasts in Supplementary Table 4F. A significant slope departure indicates miscalibration of risk spread, not superior prediction. Nonsignificance does not establish calibration across the risk range. Diagnostic fits were not applied to alter forecasts, and uncertainty is conditional on the saved predictions and censoring weights.

#### Supplementary Table 4 | Exploratory tests for genetic increments and contribution profiles

##### A. Matched PRISM-AD refits with and without PRS

Two-sided null-centered participant-bootstrap tests use 9,999 resamples within fold. BH correction includes all eight task-by-horizon-by-metric tests. Corresponding 500-resample nominal percentile intervals appear in Supplementary Table 2.

| Task, horizon | Metric | Participants | Difference | P | BH q |
|---|---|---|---|---|---|
| CN to MCI, 3 y | AUROC | 908 | 0.0026 | 0.7405 | 0.9320 |
| CN to MCI, 3 y | AP | 908 | -0.0023 | 0.8613 | 0.9320 |
| CN to MCI, 5 y | AUROC | 908 | 0.0206 | 0.0152 | 0.1216 |
| CN to MCI, 5 y | AP | 908 | -0.0013 | 0.9320 | 0.9320 |
| MCI to AD, 3 y | AUROC | 714 | 0.0046 | 0.0362 | 0.1448 |
| MCI to AD, 3 y | AP | 714 | 0.0016 | 0.6711 | 0.9320 |
| MCI to AD, 5 y | AUROC | 714 | 0.0041 | 0.0808 | 0.1616 |
| MCI to AD, 5 y | AP | 714 | 0.0055 | 0.0574 | 0.1531 |

##### B. Cognitive contributions across raw-PRS quintiles

Four participant-level HC3 omnibus F tests adjust for fitted fold and compare the modeled cognitive contribution across shared raw-PRS quintiles. BH correction covers four tests.

| Clinical-course group | n | F | P | BH q |
|---|---|---|---|---|
| All | 893 | 1.563 | 0.1821 | 0.4539 |
| CN | 701 | 1.416 | 0.2270 | 0.4539 |
| MCI | 97 | 0.794 | 0.5324 | 0.6816 |
| AD | 95 | 0.575 | 0.6816 | 0.6816 |

#### C. Genetic factors at comparable modeled cognitive risk

Shared C deciles are defined across the complete profile sample, whose participants were initially CN. AD–CN comparisons regress the signed genetic log-odds term G on clinical-course group, with continuous C and fold adjustment. The pooled model also includes decile indicators. Expanded sensitivity adds age, age squared, sex, study and PC1–5 with common-study support. HC3 robust standard errors and t inference are used. Exponentiated contrasts compare modeled genetic factors, not clinical AD odds. BH correction covers 12 planned slots. Deciles with fewer than five participants in either group are not tested and retain P=1 for correction.

| Test | AD n | CN n | Factor ratio (nominal 95% CI) | P | BH q | Status |
|---|---|---|---|---|---|---|
| Pooled | 95 | 701 | 1.135 (1.073 to 1.201) | $<0.001$ | $<0.001$ | Estimable |
| C decile 1 | 4 | 81 | — (— to —) | — | 1.0000 | Fewer than 5 per group |
| C decile 2 | 4 | 76 | — (— to —) | — | 1.0000 | Fewer than 5 per group |
| C decile 3 | 8 | 74 | 1.113 (0.947 to 1.309) | 0.1907 | 0.2860 | Estimable |
| C decile 4 | 5 | 75 | 1.105 (0.900 to 1.356) | 0.3338 | 0.4451 | Estimable |
| C decile 5 | 9 | 64 | 1.155 (0.990 to 1.348) | 0.0665 | 0.1331 | Estimable |
| C decile 6 | 8 | 73 | 1.247 (1.029 to 1.512) | 0.0249 | 0.0599 | Estimable |
| C decile 7 | 6 | 75 | 1.101 (0.956 to 1.267) | 0.1781 | 0.2860 | Estimable |
| C decile 8 | 11 | 65 | 1.339 (1.124 to 1.594) | 0.0014 | 0.0055 | Estimable |
| C decile 9 | 18 | 61 | 1.226 (1.080 to 1.392) | 0.0021 | 0.0062 | Estimable |
| C decile 10 | 22 | 57 | 1.008 (0.839 to 1.211) | 0.9326 | 1.0000 | Estimable |
| Expanded adjustment | 95 | 701 | 1.158 (1.095 to 1.224) | $<0.001$ | $<0.001$ | Estimable |

#### D. Within-fold and shared-scale sensitivity checks

Review-requested post hoc comparisons retain the common C deciles from the complete profile sample. Each within-fold genetic contrast adjusts for continuous C and decile indicators. The pooled raw-PRS contrast additionally adjusts for fold and uses the full corrected genotype roster population SD (ddof=0). HC3 residual-df t intervals are nominal. A separate four-comparison BH family covers these rows and does not change the twelve-slot family in part C. Raw PRS differences are in SD units, not odds ratios. No prediction models were refitted, and the folds have overlapping training sets.

| Comparison | AD n | CN n | Units | Estimate (nominal 95% CI) | P | BH q |
|---|---|---|---|---|---|---|
| Fold 1 | 38 | 238 | Genetic-factor ratio | 1.120 (1.045 to 1.199) | 0.0013 | 0.0018 |
| Fold 2 | 29 | 241 | Genetic-factor ratio | 1.119 (0.985 to 1.271) | 0.0838 | 0.0838 |
| Fold 3 | 28 | 222 | Genetic-factor ratio | 1.186 (1.069 to 1.315) | 0.0013 | 0.0018 |
| Pooled raw PRS | 95 | 701 | Population SD | 0.514 (0.295 to 0.734) | $<0.001$ | $<0.001$ |

#### E. Follow-up sensitivity analyses

These additional analyses used all 908 first-eligible CN prediction participants without future clinical-course exclusions. AD outcomes were reconstructed separately from the first-MCI prediction endpoints. MCI was an intermediate state, and a later diagnostic reversion did not remove the first recorded AD event. Raw PRS used the complete corrected genotype roster population SD. C slopes were allowed to differ by fitted fold, with age, age squared, sex and study adjustment. The five-year model also included fold indicators. A separate two-comparison BH family leaves the twelve-test and four-test families in parts C and D unchanged.

##### Time to first recorded AD

The Cox working model used fold-stratified baseline hazards, Efron ties and robust sandwich standard errors with nominal normal 95% intervals.

| Analysis | Participants | AD events | HR per PRS SD (nominal 95% CI) | P | BH q |
|---|---|---|---|---|---|
| First recorded AD | 908 | 99 | 1.664 (1.377 to 2.012) | $<0.001$ | $<0.001$ |

Rank-time proportional-hazards diagnostics gave P = 0.0634 for PRS and P $<0.001$ globally. The global result indicates that the proportional-hazards assumption is not supported. The reported HR therefore summarizes the prespecified working model and cannot establish a constant effect over follow-up or a fixed-time AD risk ratio.

##### Common five-year observation window

The five-year comparison included 629 known outcomes, with 31 AD events and 598 participants observed without AD through five years. The latter group includes MCI and AD recorded after five years. The 279 participants with unknown five-year AD status were not treated as controls. AD-specific reverse-Kaplan–Meier censoring weights used study/age-band strata, falling back to the pooled contributing study for age-band strata below 50 participants, with a cap of 10. Event weights used censoring survival immediately before diagnosis, while no-AD observations used censoring survival immediately before five years. No weights were clipped. The weighted effective sample size was 563.1.

| Analysis | Known n | AD n | No AD n | PRS difference, SD (nominal 95% CI) | P | BH q |
|---|---|---|---|---|---|---|
| Five-year AD status | 629 | 31 | 598 | 0.652 (0.297 to 1.006) | $<0.001$ | $<0.001$ |

The weighted least-squares coefficient is the adjusted AD-minus-no-AD difference in raw PRS population SD, not an odds ratio. HC3 residual-df t inference treats the censoring weights as fixed. No PRISM-AD prediction model was refitted, but the new Cox and weighted association models were fitted. Recorded diagnosis timing, noninformative-censoring assumptions, unavailable competing-mortality data, fitted-C uncertainty and the internally developed sample remain limitations. These analyses do not establish absolute AD risk or external validation.

#### F. Post hoc marginal contribution comparisons

All participants were CN at prediction and contributed one value to each component. Clinical-course groups were defined from all available later diagnoses, without a fixed AD horizon. Comparisons use within-fold midranks and inverse-(pair-specific stratum size + 1) weights, with tie-corrected van Elteren normal inference. No pooling of native contribution scales or adjustment for age, study or follow-up was performed.

| Component | First versus second group | n, first / second | Rank-biserial $r$ | P | BH q | Annotation |
|---|---|---|---|---|---|---|
| C | CN versus MCI | 701 / 97 | 0.136 | 0.0300 | 0.0600 | (*) |
| C | CN versus AD | 701 / 95 | 0.359 | $< 0.001$ | $< 0.001$ | *** |
| C | MCI versus AD | 97 / 95 | 0.244 | 0.0041 | 0.0104 | * |
| G | CN versus MCI | 701 / 97 | 0.182 | 0.0038 | 0.0104 | * |
| G | CN versus AD | 701 / 95 | 0.317 | $< 0.001$ | $< 0.001$ | *** |
| G | MCI versus AD | 97 / 95 | 0.110 | 0.1965 | 0.2233 | n.s. |

Positive rank-biserial r indicates larger values in the second group, based on weighted within-fold pairwise concordance. It is not a clinical risk difference or a causal effect. The six tests share one new ten-test BH family with four calibration-slope tests in Supplementary Table 3. Ordinary stars denote BH q below 0.05, 0.01 and 0.001. (*) denotes nominal P below 0.05 with q at or above 0.05. All tests are exploratory and conditional on fitted components, with unequal clinical follow-up and overlapping model-training samples.

#### Supplementary Table 5 | Paired first-visit average-precision comparisons

Differences are PRISM-AD minus reference in censoring-weighted average precision (AP). Nominal 95% percentile intervals use 500 paired participant-bootstrap resamples. Two-sided nominal P values use 9,999 null-centered paired participant-bootstrap resamples within fold (seed 42, plus-one correction). BH correction covers all 28 comparisons. Inference is conditional on fitted models and censoring weights, without model refitting. The percentile intervals are not inversions of the tests.

| Task, horizon | Model | Reference | AP difference (95% CI) | P | BH q |
|---|---|---|---|---|---|
| CN to MCI, 3 y | PRISM-AD current | Logistic current | 0.0046 (-0.0341 to 0.0340) | 0.7986 | 0.8600 |
| CN to MCI, 3 y | PRISM-AD current | HGB current | 0.0656 (0.0068 to 0.1361) | 0.0383 | 0.2503 |
| CN to MCI, 3 y | PRISM-AD current | TabPFN current | 0.0403 (-0.0190 to 0.0892) | 0.1522 | 0.4262 |
| CN to MCI, 3 y | PRISM-AD with history | PRISM-AD current | -0.0080 (-0.0311 to 0.0239) | 0.5968 | 0.6858 |
| CN to MCI, 3 y | PRISM-AD with history | HGB with history | 0.0470 (-0.0136 to 0.1122) | 0.1462 | 0.4262 |
| CN to MCI, 3 y | PRISM-AD with history | TabPFN with history | 0.0436 (-0.0133 to 0.1104) | 0.1374 | 0.4262 |
| CN to MCI, 3 y | PRISM-AD with history | HGB native weights | 0.0301 (-0.0244 to 0.0870) | 0.2734 | 0.5742 |
| CN to MCI, 5 y | PRISM-AD current | Logistic current | 0.0003 (-0.0279 to 0.0304) | 0.9817 | 0.9817 |
| CN to MCI, 5 y | PRISM-AD current | HGB current | 0.0207 (-0.0292 to 0.0693) | 0.4042 | 0.5806 |
| CN to MCI, 5 y | PRISM-AD current | TabPFN current | 0.0289 (-0.0309 to 0.0872) | 0.3420 | 0.5806 |
| CN to MCI, 5 y | PRISM-AD with history | PRISM-AD current | -0.0105 (-0.0407 to 0.0121) | 0.4562 | 0.5806 |
| CN to MCI, 5 y | PRISM-AD with history | HGB with history | -0.0260 (-0.0825 to 0.0214) | 0.2871 | 0.5742 |
| CN to MCI, 5 y | PRISM-AD with history | TabPFN with history | -0.0236 (-0.0768 to 0.0296) | 0.3887 | 0.5806 |
| CN to MCI, 5 y | PRISM-AD with history | HGB native weights | -0.0134 (-0.0652 to 0.0315) | 0.6123 | 0.6858 |
| MCI to AD, 3 y | PRISM-AD current | Logistic current | -0.0059 (-0.0173 to 0.0107) | 0.4395 | 0.5806 |
| MCI to AD, 3 y | PRISM-AD current | HGB current | 0.0154 (-0.0191 to 0.0535) | 0.4148 | 0.5806 |
| MCI to AD, 3 y | PRISM-AD current | TabPFN current | 0.0322 (0.0055 to 0.0634) | 0.0447 | 0.2503 |
| MCI to AD, 3 y | PRISM-AD with history | PRISM-AD current | -0.0069 (-0.0217 to 0.0076) | 0.3695 | 0.5806 |
| MCI to AD, 3 y | PRISM-AD with history | HGB with history | 0.0015 (-0.0252 to 0.0346) | 0.9255 | 0.9598 |
| MCI to AD, 3 y | PRISM-AD with history | TabPFN with history | -0.0093 (-0.0370 to 0.0225) | 0.5477 | 0.6668 |
| MCI to AD, 3 y | PRISM-AD with history | HGB native weights | -0.0201 (-0.0491 to 0.0124) | 0.2296 | 0.5357 |
| MCI to AD, 5 y | PRISM-AD current | Logistic current | 0.0043 (-0.0066 to 0.0146) | 0.4356 | 0.5806 |

| Task, horizon | Model | Reference | AP difference (95% CI) | P | BH q |
|---|---|---|---|---|---|
| MCI to AD, 5 y | PRISM-AD current | HGB current | 0.0362 (0.0066 to 0.0709) | 0.0325 | 0.2503 |
| MCI to AD, 5 y | PRISM-AD current | TabPFN current | 0.0812 (0.0385 to 0.1229) | $<0.001$ | 0.0252 |
| MCI to AD, 5 y | PRISM-AD with history | PRISM-AD current | -0.0106 (-0.0248 to 0.0052) | 0.1974 | 0.5025 |
| MCI to AD, 5 y | PRISM-AD with history | HGB with history | 0.0297 (0.0022 to 0.0625) | 0.0704 | 0.2816 |
| MCI to AD, 5 y | PRISM-AD with history | TabPFN with history | 0.0294 (0.0006 to 0.0590) | 0.0544 | 0.2539 |
| MCI to AD, 5 y | PRISM-AD with history | HGB native weights | 0.0394 (0.0029 to 0.0736) | 0.0203 | 0.2503 |

#### Supplementary Table 6 | Clinical functions across validation fits

CN-to-MCI main functions were estimated in models using current assessments or incorporating prior assessments. Changes are high-minus-low contributions in log-odds units across the common training-fold 10th–90th percentile range. Main functions exclude interactions. The minimum Spearman correlation is the smallest pairwise rank correlation across three fitted curves. It measures ordering rather than equality of magnitude. Overlapping training samples do not provide independent replication, and these contrasts are not intervention effects.

| Model | Years | Feature | Raw range | Fold 1 change | Fold 2 change | Fold 3 change | Min. rho |
|---|---|---|---|---|---|---|---|
| PRISM-AD current | 3 | Executive function | 0.168 to 1.573 | -0.561 | -0.857 | -0.367 | 0.880 |
| PRISM-AD current | 3 | Education | 12.000 to 20.000 | 0.781 | 0.536 | 0.626 | 0.938 |
| PRISM-AD current | 3 | Language | 0.227 to 1.681 | -0.334 | -0.377 | -0.434 | 0.912 |
| PRISM-AD current | 3 | Memory | 0.298 to 1.690 | -1.017 | -1.364 | -1.138 | 0.975 |
| PRISM-AD current | 3 | Visuospatial performance | -0.198 to 1.026 | -0.084 | 0.058 | -0.176 | 0.717 |
| PRISM-AD current | 3 | Mean cognition | 0.381 to 1.427 | -0.467 | -0.575 | -0.633 | 0.976 |
| PRISM-AD current | 3 | Age | 68.914 to 82.935 | 0.229 | 0.553 | 0.845 | 0.853 |
| PRISM-AD current | 5 | Executive function | 0.168 to 1.573 | -0.918 | -0.603 | -0.619 | 0.945 |
| PRISM-AD current | 5 | Education | 12.000 to 20.000 | 0.479 | 0.063 | 0.601 | 0.750 |
| PRISM-AD current | 5 | Language | 0.227 to 1.681 | -0.709 | -0.636 | -0.903 | 0.979 |
| PRISM-AD current | 5 | Memory | 0.298 to 1.690 | -0.724 | -0.973 | -0.853 | 0.960 |
| PRISM-AD current | 5 | Visuospatial performance | -0.198 to 1.026 | 0.189 | 0.134 | 0.058 | 0.598 |
| PRISM-AD current | 5 | Mean cognition | 0.381 to 1.427 | -0.320 | -0.626 | -0.207 | 0.701 |
| PRISM-AD current | 5 | Age | 68.914 to 82.935 | 0.917 | 1.334 | 1.291 | 0.932 |
| PRISM-AD with history | 3 | Executive function | 0.168 to 1.573 | -0.815 | -0.876 | -0.372 | 0.890 |
| PRISM-AD with history | 3 | Education | 12.000 to 20.000 | 0.779 | 0.485 | 0.640 | 0.954 |
| PRISM-AD with history | 3 | Language | 0.227 to 1.681 | -0.392 | -0.274 | -0.165 | 0.503 |
| PRISM-AD with history | 3 | Memory | 0.298 to 1.690 | -0.903 | -0.948 | -1.149 | 0.988 |
| PRISM-AD with history | 3 | Visuospatial performance | -0.198 to 1.026 | -0.175 | 0.027 | -0.385 | 0.147 |
| PRISM-AD with history | 3 | Mean cognition | 0.381 to 1.427 | -0.656 | -0.798 | -0.629 | 0.990 |
| PRISM-AD with history | 3 | Age | 68.914 to 82.935 | 0.061 | 0.174 | 0.641 | 0.414 |
| PRISM-AD with history | 5 | Executive function | 0.168 to 1.573 | -1.036 | -0.598 | -0.573 | 0.976 |
| PRISM-AD with history | 5 | Education | 12.000 to 20.000 | 0.525 | 0.108 | 0.634 | 0.886 |
| PRISM-AD with history | 5 | Language | 0.227 to 1.681 | -0.575 | -0.537 | -0.673 | 0.945 |
| PRISM-AD with history | 5 | Memory | 0.298 to 1.690 | -0.735 | -0.911 | -0.913 | 0.957 |
| PRISM-AD with history | 5 | Visuospatial performance | -0.198 to 1.026 | 0.062 | 0.101 | -0.136 | -0.401 |
| PRISM-AD with history | 5 | Mean cognition | 0.381 to 1.427 | -0.584 | -0.835 | -0.255 | 0.943 |
| PRISM-AD with history | 5 | Age | 68.914 to 82.935 | 0.436 | 1.104 | 0.704 | 0.769 |

#### Supplementary Table 7 | Follow-up in the contribution-profile sample

All 893 participants were cognitively normal at the first eligible three-year first-MCI prediction visit. Later clinical-course groups use the complete available diagnostic history and are not fixed three-year AD outcomes. The AD-versus-CN contrast uses 796 participants (95 later AD and 701 last CN), while MCI participants contribute to the shared decile boundaries only.

##### A. Overall clinical-course groups

| Later clinical course | n | With dated follow-up | Dated follow-up, years | Time to first AD, years |
|---|---|---|---|---|
| Normal cognition at last diagnosis | 701 | 701 | 6.38 [4.00, 10.20] | — |
| MCI at last diagnosis | 97 | 97 | 9.04 [5.54, 11.89] | — |
| Later AD | 95 | 95 | 7.69 [5.50, 11.50] | 6.68 [4.07, 9.18] |

Values are median [first quartile, third quartile]. Dated follow-up is the time from the index CN assessment to the last subsequent assessment with a documented cognitive diagnosis and an exact assessment age. Ages recorded as 90 years or older were not treated as exact. Time to AD uses the first subsequently recorded, dated AD diagnosis. These descriptive durations do not impose equal follow-up or establish a diagnostic lead time.

##### B. Clinical-course groups by contributing study

| Study / later course | n | With dated follow-up | Dated follow-up, years | Time to first AD, years |
|---|---|---|---|---|
| ADNI / Last CN | 129 | 129 | 4.02 [2.32, 6.00] | — |
| ADNI / Last MCI | 18 | 18 | 5.55 [2.28, 9.41] | — |
| ADNI / Later AD | 11 | 11 | 9.13 [6.16, 12.47] | 9.13 [6.16, 9.75] |
| NACC / Last CN | 531 | 531 | 7.35 [4.43, 10.99] | — |
| NACC / Last MCI | 62 | 62 | 9.43 [6.21, 12.15] | — |
| NACC / Later AD | 52 | 52 | 7.41 [5.26, 10.34] | 6.50 [3.45, 9.47] |
| ROS / Last CN | 20 | 20 | 10.47 [5.04, 14.97] | — |
| ROS / Last MCI | 13 | 13 | 10.04 [8.01, 14.63] | — |
| ROS / Later AD | 21 | 21 | 8.97 [5.99, 12.26] | 7.07 [4.98, 9.07] |
| MAP / Last CN | 21 | 21 | 4.05 [3.01, 6.92] | — |
| MAP / Last MCI | 4 | 4 | — | — |
| MAP / Later AD | 11 | 11 | 7.05 [5.29, 8.52] | 6.09 [4.25, 7.37] |

Quantiles are omitted for cells with fewer than five observed values, and counts report aggregate participant support. The summary uses the frozen clinical-course records without refitting a prognostic or AD-onset model. Unequal observation windows remain a limitation.

#### Supplementary Table 8 | High-PRS clinical-course comparisons

All groups were CN at prediction. The adjusted common-study sample contained 64 high-PRS last-CN participants, 495 other CN participants and 63 participants later diagnosed with AD. Contrasts are high-PRS last-CN minus the named reference. Models adjust for age, age squared, sex, study, PC1–5 and prediction fold. Intervals are nominal HC3/t intervals. BH q values account for all 128 planned component comparisons, including other model versions, horizons and group scopes. Last-CN groups allow intermediate MCI followed by CN and require at least three years of diagnostic follow-up. The later-AD group is not restricted by PRS. These conditional model contributions do not establish biological compensation.

| Years | Reference | Contribution | Difference (95% CI) | P | BH q |
|---|---|---|---|---|---|
| 3 | Other CN | Education | 0.013 (-0.048 to 0.074) | 0.6824 | 1.0000 |
| 3 | Later AD | Education | 0.051 (-0.035 to 0.138) | 0.2465 | 0.5348 |
| 3 | Other CN | Cognition | -0.014 (-0.210 to 0.183) | 0.8897 | 1.0000 |
| 3 | Later AD | Cognition | -0.421 (-0.696 to -0.146) | 0.0028 | 0.0133 |
| 3 | Other CN | PRS | 0.386 (0.355 to 0.418) | $<0.001$ | $<0.001$ |
| 3 | Later AD | PRS | 0.195 (0.130 to 0.260) | $<0.001$ | $<0.001$ |
| 3 | Other CN | Total prediction | 0.348 (0.111 to 0.585) | 0.0041 | 0.0171 |
| 3 | Later AD | Total prediction | -0.377 (-0.721 to -0.033) | 0.0319 | 0.1048 |
| 5 | Other CN | Education | 0.002 (-0.042 to 0.046) | 0.9291 | 1.0000 |
| 5 | Later AD | Education | 0.027 (-0.033 to 0.087) | 0.3795 | 0.6672 |
| 5 | Other CN | Cognition | -0.006 (-0.196 to 0.183) | 0.9469 | 1.0000 |
| 5 | Later AD | Cognition | -0.444 (-0.728 to -0.159) | 0.0023 | 0.0122 |
| 5 | Other CN | PRS | 0.533 (0.494 to 0.572) | $<0.001$ | $<0.001$ |
| 5 | Later AD | PRS | 0.272 (0.185 to 0.359) | $<0.001$ | $<0.001$ |
| 5 | Other CN | Total prediction | 0.490 (0.299 to 0.681) | $<0.001$ | $<0.001$ |
| 5 | Later AD | Total prediction | -0.234 (-0.532 to 0.065) | 0.1246 | 0.3624 |

#### Supplementary Table 9 | Education at high genetic risk and older age

This separate retrospective analysis compares education among participants with high genetic risk who were aged 75 or older at their last documented cognitive diagnosis. Eligibility does not require model-validation participation, and the CN group excludes anyone with preceding recorded AD. The final-record-confirmed analysis requires an explicit diagnosis at the final recorded visit. Education is measured at the last diagnostic assessment, or at the paired CN visit in PET subsets.

| Comparison | Group | Eligible | Education observed | Mean years | SD |
|---|---|---|---|---|---|
| Primary | CN | 411 | 410 | 16.31 | 2.76 |
| Primary | AD | 751 | 747 | 15.37 | 3.25 |
| Final record confirmed | CN | 340 | 339 | 16.32 | 2.76 |
| Final record confirmed | AD | 734 | 730 | 15.37 | 3.25 |
| CN-stage PET | CN | 97 | 96 | 16.81 | 2.71 |
| CN-stage PET | AD | 6 | 6 | 15.67 | 3.14 |
| Amyloid-positive CN-stage PET | CN | 56 | 56 | 17.11 | 2.47 |
| Amyloid-positive CN-stage PET | AD | 5 | 5 | 16.00 | 3.39 |

Adjusted CN-minus-AD differences control for age, age squared, sex, study, continuous PRS and PC1–5. PET comparisons also account for tracer and the interval to the last diagnostic assessment. HC3 robust t-based intervals are nominal. BH correction retains four comparison slots, with unestimable tests assigned P=1. Studies require both groups to be represented.

| Comparison | CN n | AD n | Years (95% CI) | P | BH q | Status |
|---|---|---|---|---|---|---|
| Primary | 293 | 649 | 0.97 (0.58 to 1.35) | $<0.001$ | $<0.001$ | Estimable |
| Final record confirmed | 270 | 636 | 1.01 (0.62 to 1.40) | $<0.001$ | $<0.001$ | Estimable |
| CN-stage PET | 20 | 5 | — (— to —) | — | — | Insufficient group support |
| Amyloid-positive CN-stage PET | 0 | 0 | — (— to —) | — | — | Insufficient group support |

#### Supplementary Table 10 | PRS and amyloid PET burden during normal cognition

Exploratory post hoc associations use one quality-controlled CN-stage scan per participant, with a paired cognitive assessment within 90 days and no preceding recorded MCI or AD. The older population retained normal cognition at age 75 or older at the last diagnostic assessment. The same frozen raw high-PRS interval was applied to the corrected scores. Continuous PRS was standardized using the complete post-QC genotype roster. Models adjust for scan age, sex, study, tracer and PC1–5. Lag-adjusted results condition additionally on post-PET follow-up and are sensitivity analyses only. The small subgroup with at least three years of follow-up omits tracer to avoid a singleton high-leverage category. Intervals and P values use HC3 robust t inference and are not multiplicity adjusted. These selected subsets provide biological context, not external prognostic validation.

| Analysis scope | n | Centiloids per PRS SD (95% CI) | Nominal P | Spearman rho |
|---|---|---|---|---|
| Older CN, all PRS | 802 | 14.1 (10.4 to 17.8) | $<0.001$ | 0.249 |
| Older CN, high PRS | 97 | 21.6 (-15.3 to 58.5) | 0.2478 | 0.107 |
| Older CN, other PRS | 705 | 13.1 (8.3 to 18.0) | $<0.001$ | 0.191 |
| Older CN, final-record confirmed | 577 | 5.6 (2.4 to 8.8) | $<0.001$ | 0.143 |
| Older CN, high PRS, final-record confirmed | 54 | -1.2 (-34.1 to 31.7) | 0.9417 | -0.029 |
| Older CN, follow-up adjusted | 783 | 14.3 (10.5 to 18.0) | $<0.001$ | 0.254 |
| Older CN, high PRS, follow-up adjusted | 96 | 20.6 (-17.7 to 59.0) | 0.2877 | 0.086 |
| All-age CN, all PRS | 3030 | 12.2 (10.8 to 13.7) | $<0.001$ | 0.279 |
| All-age CN, high PRS | 462 | 20.0 (6.9 to 33.1) | 0.0028 | 0.135 |
| All-age CN, other PRS | 2568 | 11.2 (9.4 to 13.0) | $<0.001$ | 0.213 |
| All-age CN, high PRS, 3-year follow-up | 48 | 23.6 (-13.1 to 60.2) | 0.2009 | 0.163 |
| All-age CN, high PRS, A4 | 386 | 18.2 (3.5 to 32.9) | 0.0155 | 0.127 |
| All-age CN, high PRS, ADNI | 53 | 38.3 (9.1 to 67.5) | 0.0114 | 0.376 |

#### Supplementary Table 11 | Participant characteristics and predictor missingness

Each participant contributes the first eligible visit for the indicated clinical task. Development includes folds 1–9. The evaluated subset comprises folds 1–3, held out once for internal validation. The evaluated subset is part of the development population, and participants may contribute to both clinical tasks. The columns therefore describe overlapping populations.

##### A. Characteristics at the first eligible visit

Continuous variables are median [first quartile, third quartile] among observed values. Female percentages and study percentages use all participants in the column. PHC sex code 2 denotes female. Cognitive scores and their descriptive available-domain mean retain PHC units. PRS values are the corrected, unstandardized scores calculated before stage-specific analyses.

| Characteristic | CN development | CN evaluated | MCI development | MCI evaluated |
|---|---|---|---|---|
| Participants | 2759 | 908 | 2082 | 714 |
| Female, n (%) | 1722 (62.4) | 565 (62.2) | 1006 (48.3) | 336 (47.1) |
| Age, years | 74.2 [70.3, 78.7] | 74.2 [70.3, 78.9] | 73.9 [68.5, 79.7] | 74.4 [68.5, 79.7] |
| Education, years | 16.0 [15.0, 18.0] | 16.0 [15.0, 18.0] | 16.0 [14.0, 18.0] | 16.0 [14.0, 18.0] |
| Memory, PHC units | 0.74 [0.45, 1.07] | 0.74 [0.40, 1.06] | 0.08 [-0.27, 0.46] | 0.10 [-0.25, 0.54] |
| Executive function, PHC units | 0.80 [0.44, 1.17] | 0.81 [0.45, 1.16] | 0.36 [-0.04, 0.76] | 0.37 [-0.07, 0.75] |
| Language, PHC units | 0.77 [0.47, 1.16] | 0.77 [0.46, 1.16] | 0.37 [-0.02, 0.72] | 0.39 [-0.01, 0.75] |
| Visuospatial performance, PHC units | 0.62 [0.15, 0.91] | 0.42 [0.15, 0.91] | 0.15 [-0.20, 0.91] | 0.15 [-0.20, 0.91] |
| Available-domain mean, PHC units | 0.79 [0.52, 1.05] | 0.79 [0.52, 1.03] | 0.28 [-0.01, 0.57] | 0.27 [-0.03, 0.59] |
| Measured cognitive domains | 3 [3, 4] | 3 [3, 4] | 3 [3, 4] | 3 [3, 4] |
| Unstandardized PRS | -0.135 [-0.442, 0.284] | -0.135 [-0.450, 0.282] | 0.135 [-0.288, 0.607] | 0.149 [-0.296, 0.627] |
| ADNI, n (%) | 507 (18.4) | 159 (17.5) | 736 (35.4) | 254 (35.6) |
| NACC, n (%) | 1954 (70.8) | 652 (71.8) | 1113 (53.5) | 376 (52.7) |
| ROS, n (%) | 174 (6.3) | 60 (6.6) | 148 (7.1) | 57 (8.0) |
| MAP, n (%) | 124 (4.5) | 37 (4.1) | 85 (4.1) | 27 (3.8) |

#### B. Predictor missingness before imputation

| Missing predictor, n (%) | CN development | CN evaluated | MCI development | MCI evaluated |
|---|---|---|---|---|
| Age | 0 (0.0) | 0 (0.0) | 0 (0.0) | 0 (0.0) |
| Sex | 0 (0.0) | 0 (0.0) | 0 (0.0) | 0 (0.0) |
| Education | 9 (0.3) | 7 (0.8) | 4 (0.2) | 2 (0.3) |
| Memory | 0 (0.0) | 0 (0.0) | 2 (0.1) | 1 (0.1) |
| Executive function | 2 (0.1) | 0 (0.0) | 1 (0.0) | 0 (0.0) |
| Language | 0 (0.0) | 0 (0.0) | 0 (0.0) | 0 (0.0) |
| Visuospatial performance | 1956 (70.9) | 652 (71.8) | 1116 (53.6) | 377 (52.8) |
| Available-domain mean | 0 (0.0) | 0 (0.0) | 0 (0.0) | 0 (0.0) |
| Measured cognitive-domain count | 0 (0.0) | 0 (0.0) | 0 (0.0) | 0 (0.0) |
| PRS | 0 (0.0) | 0 (0.0) | 0 (0.0) | 0 (0.0) |

Counts precede training-only imputation and scaling. Eligibility required at least two measured cognitive domains, so the available-domain mean and domain count are available by construction. Missing domain measurements were represented by indicators during model fitting. Non-PRS clinical inputs, participant allocations and outcome labels were unchanged during PRS reconstruction.

**Supplementary Table 12 | Age and outcome availability at prediction.** Age is measured at the first eligible prediction visit, not at the final diagnosis. All values are descriptive, and counts are unweighted. The same participants occur in both horizons within a clinical task.

**CN-to-MCI**

| Horizon | Outcome group | $n$ | Age, median (IQR) | Age ≥75, $n$ (%) |
|---|---|---|---|---|
| 3 years | All participants | 908 | 74.2 (70.3–78.9) | 421 (46.4) |
| 3 years | Recorded MCI | 78 | 76.2 (72.6–80.1) | 49 (62.8) |
| 3 years | No MCI or AD | 717 | 73.8 (70.0–78.5) | 309 (43.1) |
| 3 years | AD before MCI | 6 | 78.3 (73.4–83.1) | 4 (66.7) |
| 3 years | Follow-up incomplete | 107 | 76.0 (71.6–80.8) | 59 (55.1) |
| 5 years | All participants | 908 | 74.2 (70.3–78.9) | 421 (46.4) |
| 5 years | Recorded MCI | 124 | 76.5 (72.6–80.1) | 77 (62.1) |
| 5 years | No MCI or AD | 525 | 73.2 (69.7–77.8) | 203 (38.7) |
| 5 years | AD before MCI | 9 | 78.7 (72.0–83.5) | 6 (66.7) |
| 5 years | Follow-up incomplete | 250 | 75.8 (70.9–81.1) | 135 (54.0) |

**MCI-to-AD**

| Horizon | Outcome group | $n$ | Age, median (IQR) | Age ≥75, $n$ (%) |
|---|---|---|---|---|
| 3 years | All participants | 714 | 74.4 (68.5–79.7) | 337 (47.2) |
| 3 years | Recorded AD | 230 | 74.9 (68.2–80.1) | 114 (49.6) |
| 3 years | No recorded AD | 345 | 74.5 (68.5–79.3) | 159 (46.1) |
| 3 years | Follow-up incomplete | 139 | 73.7 (68.9–79.9) | 64 (46.0) |
| 5 years | All participants | 714 | 74.4 (68.5–79.7) | 337 (47.2) |
| 5 years | Recorded AD | 294 | 75.1 (68.1–80.3) | 147 (50.0) |
| 5 years | No recorded AD | 192 | 74.2 (68.9–78.0) | 79 (41.1) |
| 5 years | Follow-up incomplete | 228 | 74.1 (68.5–79.8) | 111 (48.7) |

**Interpretation of age in the current analysis.** All compared prognostic models included age at prediction. Censoring weights were estimated from training data within study and age bands, with fallback to study-level or pooled curves when support was insufficient. These procedures do not standardize performance to age 75 or evaluate performance exclusively among participants aged 75 and above. Prediction visits at age 85 or above were excluded in the existing analysis. The separate education comparison required age 75 or above at the last diagnostic assessment. That criterion was not applied to these prediction samples.

No-event status applies only to the indicated horizon. Participants without recorded AD in the MCI task may still have cognitive impairment, while direct AD before MCI is a competing event in the CN task and cannot be treated as a healthy control. The distributions do not establish age-independent discrimination, calibrated risk at a fixed age, or freedom from future disease. This descriptive age summary uses the same clinical population as the corrected-PRS model evaluation and adds no age-specific model fits.

#### Supplementary optimization diagnostics

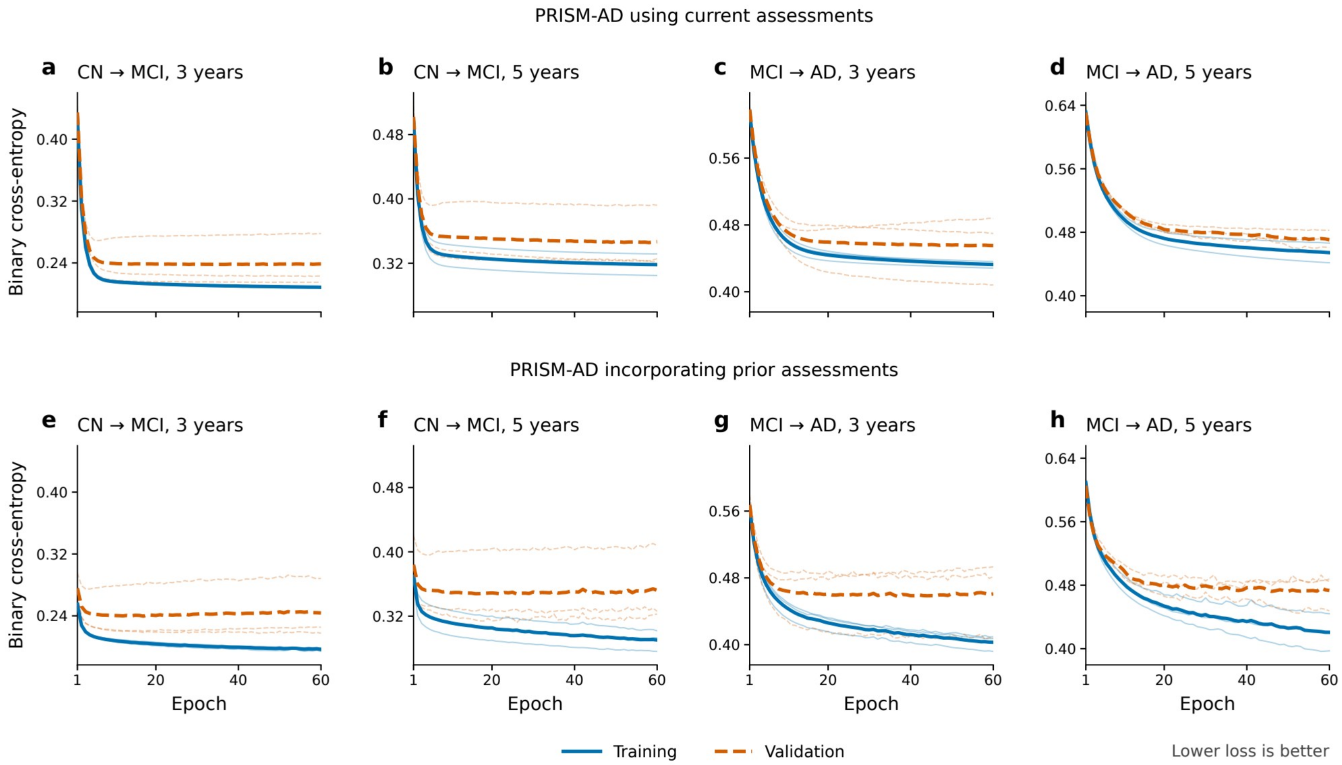


**Supplementary Figure 1 | Training and validation loss trajectories for PRISM-AD. a–d,** Models using current assessments. **e–h,** Models incorporating prior assessments. Columns show CN-to-MCI at three and five years, followed by MCI-to-AD at three and five years. Thin curves show the three prespecified internal validation folds, and thick curves show their unweighted means. Blue solid curves show end-of-epoch binary cross-entropy on the original fixed inverse-probability-of-censoring-weighted (IPCW) training resample. Orange dashed curves show IPCW binary cross-entropy across all eligible held-out visits with known outcomes, normalized by the sum of weights. These visit-level diagnostics differ from the primary evaluation, which uses the first eligible visit per participant. Both losses were evaluated with dropout disabled and no regularization penalty. Their difference reflects fitting behavior and differences between the training resample and weighted validation observations. The 24 fits with PRS underlying the primary model comparisons used 60 fixed epochs. Retrospective replay of the corrected-PRS fits reproduced every recorded training loss, final model parameter and held-out prediction exactly. The final epoch was retained under the original schedule, without using these retrospectively reconstructed losses to select a checkpoint. Validation loss generally decreased early and then leveled off, with modest late increases in models incorporating prior assessments. CN, cognitively normal. MCI, mild cognitive impairment. AD, Alzheimer’s disease dementia.

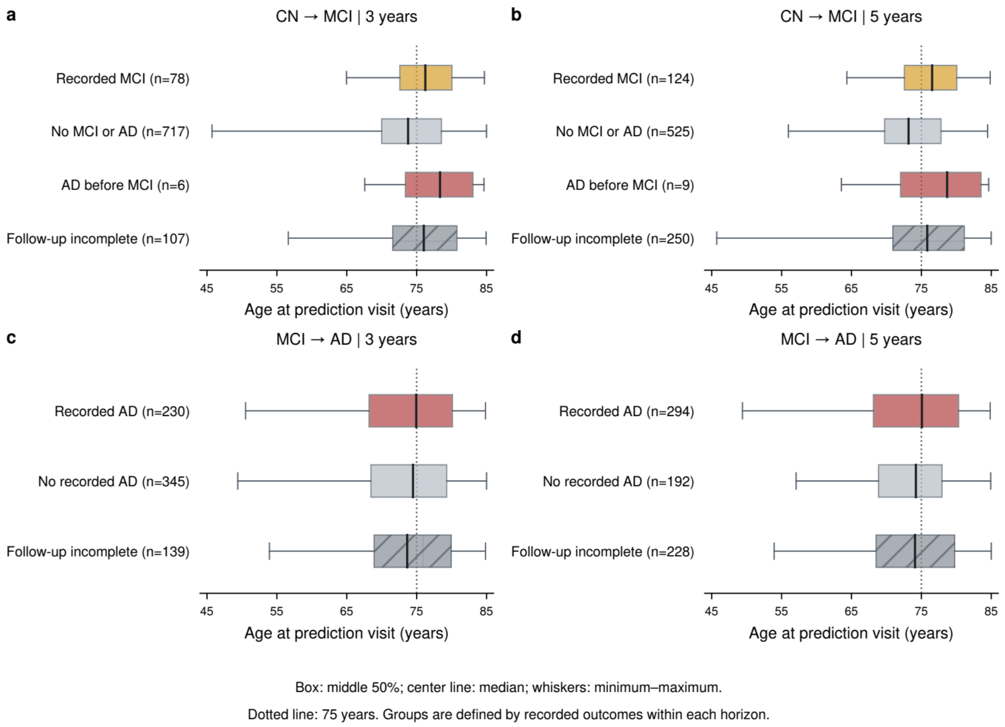


Supplementary Figure 2 | Age at prediction in the stage-specific validation samples. Age at the first eligible prediction visit is shown for participants with normal cognition (a,b) or MCI (c,d), for three-year (a,c) and five-year (b,d) outcomes. Each participant contributes once per task and horizon and may enter both clinical tasks. Samples match Table 1 and Fig. 2 and pool the three completed validation folds. Target events occurred by the stated horizon. No-event status required observed follow-up without the specified event through that horizon. Direct recorded AD before MCI is shown separately in the CN task. For participants whose follow-up ended early without a target or competing event, the horizon outcome remained unknown. Boxes span the first to third quartile. The central line is the median and whiskers span the full observed range. Counts and distributions are unweighted. The dotted line marks age 75 for interpretation, not an eligibility threshold for the prediction analyses. CN, cognitively normal. MCI, mild cognitive impairment. AD, Alzheimer’s disease dementia.

#### Supplementary genetic-increment analysis

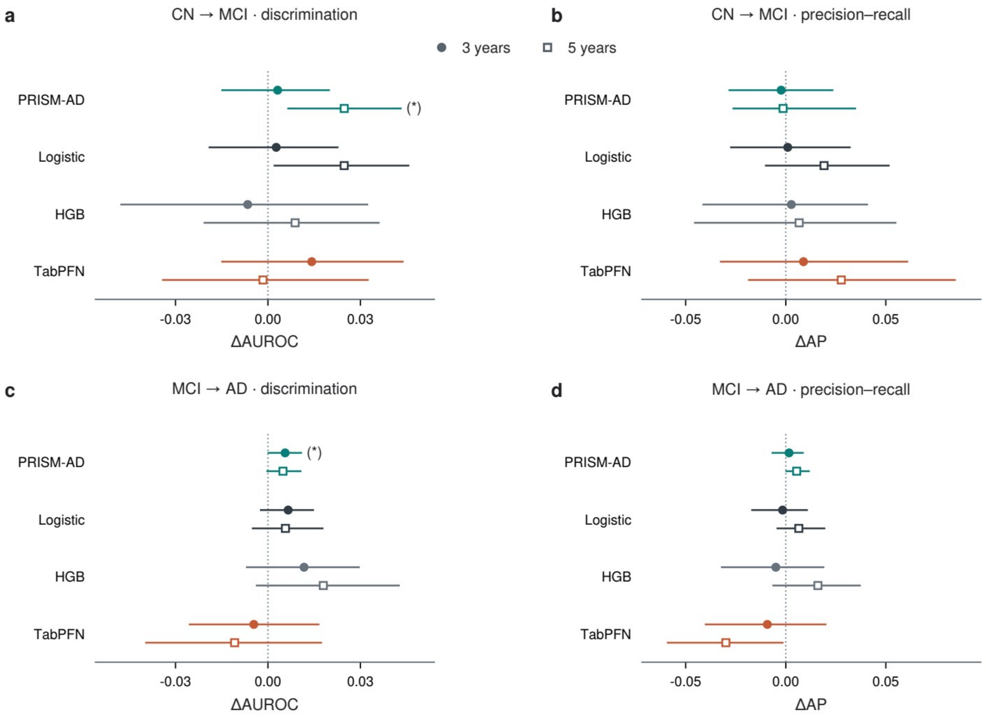


**Supplementary Figure 3 | Incremental prediction from the polygenic risk score. a,b,** CN-to-MCI. **c,d,** MCI-to-AD. Models were fitted separately with and without PRS using the same clinical inputs. Points show the difference in censoring-weighted AUROC (a,c) or average precision (AP, b,d) between these paired fits. Filled circles indicate three years and open squares five years. Colors identify the four models. Lines are nominal 95% percentile intervals from 500 paired participant-bootstrap resamples within validation fold, conditional on the fitted prediction and censoring models. Participants, clinical inputs, training resamples and settings were held fixed between input sets. Evaluation used the first eligible visit per participant in development-validation folds 1–3: 908 CN and 714 MCI participants, including those with unknown horizon outcomes. Only known outcomes contribute to the weighted discrimination metrics. All contrasts are shown. Intervals are nominal and are not multiplicity adjusted. Ordinary *, ** and *** indicate PRISM-AD BH $q < 0.05$, 0.01 and 0.001 across eight task-by-horizon-by-metric tests. The symbol (*) indicates nominal $P < 0.05$ with $q \geq 0.05$. All eight tests are reported in Supplementary Table 4. Other model increments are not part of that test family and have no inference markers. Positive differences indicate higher discrimination with PRS, while clinical benefit remains untested. These analyses do not evaluate an untouched test set or external cohort.

#### Supplementary Acknowledgements

##### Data resources

We thank the participants, families, investigators and staff who contributed data to the Alzheimer's Disease Sequencing Project (ADSP), the ADSP Phenotype Harmonization Consortium (ADSP-PHC) and the source studies. The statements below acknowledge the generation, harmonization and sharing of these data, rather than direct funding of the present secondary analysis. The ADSP umbrella statement describes the wider resource; not every cohort named in that statement contributed participants to each analysis reported here.

Data for this study were prepared, archived, and distributed by the National Institute on Aging Alzheimer's Disease Data Storage Site (NIAGADS) at the University of Pennsylvania (U24-AG041689), funded by the National Institute on Aging.

##### Alzheimer's Disease Sequencing Project (sa000001)

*The Alzheimer's Disease Sequencing Project (ADSP) is comprised of two Alzheimer's Disease (AD) genetics consortia and three National Human Genome Research Institute (NHGRI) funded Large Scale Sequencing and Analysis Centers (LSAC). The two AD genetics consortia are the Alzheimer's Disease Genetics Consortium (ADGC) funded by NIA (U01 AG032984), and the Cohorts for Heart and Aging Research in Genomic Epidemiology (CHARGE) funded by NIA (R01 AG033193), the National Heart, Lung, and Blood Institute (NHLBI), other National Institute of Health (NIH) institutes and other foreign governmental and non-governmental organizations. The Discovery Phase analysis of sequence data is supported through UF1AG047133 (to Drs. Schellenberg, Farrer, Pericak-Vance, Mayeux, and Haines); U01AG049505 to Dr. Seshadri; U01AG049506 to Dr. Boerwinkle; U01AG049507 to Dr. Wijsman; and U01AG049508 to Dr. Goate and the Discovery Extension Phase analysis is supported through U01AG052411 to Dr. Goate, U01AG052410 to Dr. Pericak-Vance and U01 AG052409 to Drs. Seshadri and Fornage.*

*Sequencing for the Follow Up Study (FUS) is supported through U01AG057659 (to Drs. PericakVance, Mayeux, and Vardarajan) and U01AG062943 (to Drs. Pericak-Vance and Mayeux). Data generation and harmonization in the Follow-up Phase is supported by U54AG052427 (to Drs. Schellenberg and Wang). The FUS Phase analysis of sequence data is supported through U01AG058589 (to Drs. Destefano, Boerwinkle, De Jager, Fornage, Seshadri, and Wijsman), U01AG058654 (to Drs. Haines, Bush, Farrer, Martin, and Pericak-Vance), U01AG058635 (to Dr. Goate), RF1AG058066 (to Drs. Haines, Pericak-Vance, and Scott), RF1AG057519 (to Drs. Farrer and Jun), R01AG048927 (to Dr. Farrer), and RF1AG054074 (to Drs. Pericak-Vance and Beecham).*

*The ADGC cohorts include: Adult Changes in Thought (ACT) (U01 AG006781, U19 AG066567), the Alzheimer's Disease Research Centers (ADRC) (P30 AG062429, P30 AG066468, P30 AG062421, P30 AG066509, P30 AG066514, P30 AG066530, P30 AG066507, P30 AG066444, P30 AG066518, P30 AG066512, P30 AG066462, P30 AG072979, P30 AG072972, P30 AG072976, P30 AG072975, P30 AG072978, P30 AG072977, P30 AG066519, P30 AG062677, P30 AG079280, P30 AG062422, P30 AG066511, P30 AG072946, P30 AG062715, P30 AG072973, P30 AG066506, P30 AG066508, P30 AG066515, P30 AG072947, P30 AG072931, P30 AG066546, P20 AG068024, P20 AG068053, P20 AG068077, P20 AG068082, P30 AG072958, P30 AG072959), the Chicago Health and Aging Project (CHAP) (R01 AG11101, RC4 AG039085,*

*K23 AG030944), Indiana Memory and Aging Study (IMAS) (R01 AG019771), Indianapolis Ibadan (R01 AG009956, P30 AG010133), the Memory and Aging Project (MAP) ( R01 AG17917), Mayo Clinic (MAYO) (R01 AG032990, U01 AG046139, R01 NS080820, RF1 AG051504, P50 AG016574), Mayo Parkinson's Disease controls (NS039764, NS071674, 5RC2HG005605), University of Miami (R01 AG027944, R01 AG028786, R01 AG019085, IIRG09133827, A2011048), the Multi-Institutional Research in Alzheimer's Genetic Epidemiology Study (MIRAGE) (R01 AG09029, R01 AG025259), the National Centralized Repository for Alzheimer's Disease and Related Dementias (NCRAD) (U24 AG021886), the National Institute on Aging Late Onset Alzheimer's Disease Family Study (NIA- LOAD) (U24 AG056270), the Religious Orders Study (ROS) (P30 AG10161, R01 AG15819), the Texas Alzheimer's Research and Care Consortium (TARCC) (funded by the Darrell K Royal Texas Alzheimer's Initiative), Vanderbilt University/Case Western Reserve University (VAN/CWRU) (R01 AG019757, R01 AG021547, R01 AG027944, R01 AG028786, P01 NS026630, and Alzheimer's Association), the Washington Heights-Inwood Columbia Aging Project (WHICAP) (RF1 AG054023), the University of Washington Families (VA Research Merit Grant, NIA: P50AG005136, R01AG041797, NINDS: R01NS069719), the Columbia University Hispanic Estudio Familiar de Influencia Genetica de Alzheimer (EFIGA) (RF1 AG015473), the University of Toronto (UT) (funded by Wellcome Trust, Medical Research Council, Canadian Institutes of Health Research), and Genetic Differences (GD) (R01 AG007584). The CHARGE cohorts are supported in part by National Heart, Lung, and Blood Institute (NHLBI) infrastructure grant HL105756 (Psaty), RC2HL102419 (Boerwinkle) and the neurology working group is supported by the National Institute on Aging (NIA) R01 grant AG033193.*

*The CHARGE cohorts participating in the ADSP include the following: Austrian Stroke Prevention Study (ASPS), ASPS-Family study, and the Prospective Dementia Registry-Austria (ASPS/PRODEM-Aus), the Atherosclerosis Risk in Communities (ARIC) Study, the Cardiovascular Health Study (CHS), the Erasmus Rucphen Family Study (ERF), the Framingham Heart Study (FHS), and the Rotterdam Study (RS). ASPS is funded by the Austrian Science Fond (FWF) grant number P20545-P05 and P13180 and the Medical University of Graz. The ASPS-Fam is funded by the Austrian Science Fund (FWF) project I904), the EU Joint Programme – Neurodegenerative Disease Research (JPND) in frame of the BRIDGET project (Austria, Ministry of Science) and the Medical University of Graz and the Steiermärkische Krankenanstalten Gesellschaft. PRODEM-Austria is supported by the Austrian Research Promotion agency (FFG) (Project No. 827462) and by the Austrian National Bank (Anniversary Fund, project 15435. ARIC research is carried out as a collaborative study supported by NHLBI contracts (HHSN268201100005C, HHSN268201100006C, HHSN268201100007C, HHSN268201100008C, HHSN268201100009C, HHSN268201100010C, HHSN268201100011C, and HHSN268201100012C). Neurocognitive data in ARIC is collected by U01 2U01HL096812, 2U01HL096814, 2U01HL096899, 2U01HL096902, 2U01HL096917 from the NIH (NHLBI, NINDS, NIA and NIDCD), and with previous brain MRI examinations funded by R01-HL70825 from the NHLBI. CHS research was supported by contracts HHSN268201200036C, HHSN268200800007C, N01HC55222, N01HC85079, N01HC85080, N01HC85081, N01HC85082, N01HC85083, N01HC85086, and grants U01HL080295 and U01HL130114 from the NHLBI with additional contribution from the National Institute of Neurological Disorders and Stroke (NINDS). Additional support was provided by R01AG023629, R01AG15928, and R01AG20098 from the NIA. FHS research is supported by NHLBI contracts N01-HC-25195 and HHSN268201500001I. This study was also supported by additional grants from the NIA (R01s AG054076, AG049607 and*

*AG033040 and NINDS (R01 NS017950). The ERF study as a part of EUROSPAN (European Special Populations Research Network) was supported by European Commission FP6 STRP grant number 018947 (LSHG-CT-2006-01947) and also received funding from the European Community's Seventh Framework Programme (FP7/2007-2013)/grant agreement HEALTH-F4- 2007-201413 by the European Commission under the programme "Quality of Life and Management of the Living Resources" of 5th Framework Programme (no. QLG2-CT-2002- 01254). High-throughput analysis of the ERF data was supported by a joint grant from the Netherlands Organization for Scientific Research and the Russian Foundation for Basic Research (NWO-RFBR 047.017.043). The Rotterdam Study is funded by Erasmus Medical Center and Erasmus University, Rotterdam, the Netherlands Organization for Health Research and Development (ZonMw), the Research Institute for Diseases in the Elderly (RIDE), the Ministry of Education, Culture and Science, the Ministry for Health, Welfare and Sports, the European Commission (DG XII), and the municipality of Rotterdam. Genetic data sets are also supported by the Netherlands Organization of Scientific Research NWO Investments (175.010.2005.011, 911-03-012), the Genetic Laboratory of the Department of Internal Medicine, Erasmus MC, the Research Institute for Diseases in the Elderly (014-93-015; RIDE2), and the Netherlands Genomics Initiative (NGI)/Netherlands Organization for Scientific Research (NWO) Netherlands Consortium for Healthy Aging (NCHA), project 050-060-810. All studies are grateful to their participants, faculty and staff. The content of these manuscripts is solely the responsibility of the authors and does not necessarily represent the official views of the National Institutes of Health or the U.S. Department of Health and Human Services.*

*The FUS cohorts include: the Alzheimer's Disease Research Centers (ADRC) (P30 AG062429, P30 AG066468, P30 AG062421, P30 AG066509, P30 AG066514, P30 AG066530, P30 AG066507, P30 AG066444, P30 AG066518, P30 AG066512, P30 AG066462, P30 AG072979, P30 AG072972, P30 AG072976, P30 AG072975, P30 AG072978, P30 AG072977, P30 AG066519, P30 AG062677, P30 AG079280, P30 AG062422, P30 AG066511, P30 AG072946, P30 AG062715, P30 AG072973, P30 AG066506, P30 AG066508, P30 AG066515, P30 AG072947, P30 AG072931, P30 AG066546, P20 AG068024, P20 AG068053, P20 AG068077, P20 AG068082, P30 AG072958, P30 AG072959), Alzheimer's Disease Neuroimaging Initiative (ADNI) (U19AG024904), Amish Protective Variant Study (RF1AG058066), Cache County Study (R01AG11380, R01AG031272, R01AG21136, RF1AG054052), Case Western Reserve University Brain Bank (CWRUBB) (P50AG008012), Case Western Reserve University Rapid Decline (CWRURD) (RF1AG058267, NU38CK000480), CubanAmerican Alzheimer's Disease Initiative (CuAADI) (3U01AG052410), Estudio Familiar de Influencia Genetica en Alzheimer (EFIGA) (5R37AG015473, RF1AG015473, R56AG051876), Genetic and Environmental Risk Factors for Alzheimer Disease Among African Americans Study (GenerAAtions) (2R01AG09029, R01AG025259, 2R01AG048927), Gwangju Alzheimer and Related Dementias Study (GARD) (U01AG062602), Hillblom Aging Network (2014-A-004-NET, R01AG032289, R01AG048234), Hussman Institute for Human Genomics Brain Bank (HIHGBB) (R01AG027944, Alzheimer's Association "Identification of Rare Variants in Alzheimer Disease"), Ibadan Study of Aging (IBADAN) (5R01AG009956), Longevity Genes Project (LGP) and LonGenity (R01AG042188, R01AG044829, R01AG046949, R01AG057909, R01AG061155, P30AG038072), Mexican Health and Aging Study (MHAS) (R01AG018016), Multi-Institutional Research in Alzheimer's Genetic Epidemiology (MIRAGE) (2R01AG09029, R01AG025259, 2R01AG048927), Northern Manhattan Study (NOMAS) (R01NS29993), Peru Alzheimer's Disease Initiative (PeADI) (RF1AG054074), Puerto Rican 1066 (PR1066) (Wellcome Trust (GR066133/GR080002), European Research*

*Council (340755)), Puerto Rican Alzheimer Disease Initiative (PRADI) (RF1AG054074), Reasons for Geographic and Racial Differences in Stroke (REGARDS) (U01NS041588), Research in African American Alzheimer Disease Initiative (REAAADI) (U01AG052410), the Religious Orders Study (ROS) (P30 AG10161, P30 AG72975, R01 AG15819, R01 AG42210), the RUSH Memory and Aging Project (MAP) (R01 AG017917, R01 AG42210Stanford Extreme Phenotypes in AD (R01AG060747), University of Miami Brain Endowment Bank (MBB), University of Miami/Case Western/North Carolina A&T African American (UM/CASE/NCAT) (U01AG052410, R01AG028786), Wisconsin Registry for Alzheimer's Prevention (WRAP) (R01AG027161 and R01AG054047), Mexico-Southern California Autosomal Dominant Alzheimer's Disease Consortium (R01AG069013), Center for Cognitive Neuroscience and Aging (R01AG047649), and the A4 Study (R01AG063689, U19AG010483 and U24AG057437).*

*The four LSACs are: the Human Genome Sequencing Center at the Baylor College of Medicine (U54 HG003273), the Broad Institute Genome Center (U54HG003067), The American Genome Center at the Uniformed Services University of the Health Sciences (U01AG057659), and the Washington University Genome Institute (U54HG003079*). *Genotyping and sequencing for the ADSP FUS is also conducted at John P. Hussman Institute for Human Genomics (HIHG) Center for Genome Technology (CGT).*

*Biological samples and associated phenotypic data used in primary data analyses were stored at Study Investigators institutions, and at the National Centralized Repository for Alzheimer's Disease and Related Dementias (NCRAD, U24AG021886) at Indiana University funded by NIA. Associated Phenotypic Data used in primary and secondary data analyses were provided by Study Investigators, the NIA funded Alzheimer's Disease Centers (ADCs), and the National Alzheimer's Coordinating Center (NACC, U24AG072122) and the National Institute on Aging Genetics of Alzheimer's Disease Data Storage Site (NIAGADS, U24AG041689) at the University of Pennsylvania, funded by NIA. Harmonized phenotypes were provided by the ADSP Phenotype Harmonization Consortium (ADSP-PHC), funded by NIA (U24 AG074855, U01 AG068057 and R01 AG059716) and Ultrascale Machine Learning to Empower Discovery in Alzheimer's Disease Biobanks (AI4AD, U01 AG068057). This research was supported in part by the Intramural Research Program of the National Institutes of health, National Library of Medicine. Contributors to the Genetic Analysis Data included Study Investigators on projects that were individually funded by NIA, and other NIH institutes, and by private U.S. organizations, or foreign governmental or nongovernmental organizations.*

The ADSP Phenotype Harmonization Consortium (ADSP-PHC) is funded by NIA (U24 AG074855, U01 AG068057 and R01 AG059716). The harmonized cohorts within the ADSP-PHC include: the Anti-Amyloid Treatment in Asymptomatic Alzheimer's study (A4 Study), a secondary prevention trial in preclinical Alzheimer's disease, aiming to slow cognitive decline associated with brain amyloid accumulation in clinically normal older individuals. The A4 Study is funded by a public-private-philanthropic partnership, including funding from the National Institutes of Health-National Institute on Aging, Eli Lilly and Company, Alzheimer's Association, Accelerating Medicines Partnership, GHR Foundation, an anonymous foundation and additional private donors, with in-kind support from Avid and Cogstate. The companion observational Longitudinal Evaluation of Amyloid Risk and Neurodegeneration (LEARN) Study is funded by the Alzheimer's Association and GHR Foundation. The A4 and A4-LEARN Studies are led by Dr. Reisa Sperling at Brigham and Women's Hospital, Harvard Medical School and Dr. Paul Aisen at the Alzheimer's Therapeutic Research Institute (ATRI), University of Southern California. The A4 and LEARN Studies are coordinated by ATRI at the University of Southern California, and the data are made available through the Laboratory for Neuro Imaging at the University of Southern California. The participants screening for the A4 Study provided permission to share their de-identified data in order to advance the quest to find a successful treatment for Alzheimer's disease. We would like to acknowledge the dedication of all the participants, the site personnel, and all of the partnership team members who continue to make the A4 and LEARN Studies possible. The complete A4 Study Team list is available on: a4study.org/a4-study-team.; the Adult Changes in Thought study (ACT), U01 AG006781, U19 AG066567; Alzheimer's Disease Neuroimaging Initiative (ADNI): Data collection and sharing for this project was funded by the Alzheimer's Disease Neuroimaging Initiative (ADNI) (National Institutes of Health Grant U01 AG024904) and DOD ADNI (Department of Defense award number W81XWH-12-2-0012). ADNI is funded by the National Institute on Aging, the National Institute of Biomedical Imaging and Bioengineering, and through generous contributions from the following: AbbVie, Alzheimer's Association; Alzheimer's Drug Discovery Foundation; Araclon Biotech; BioClinica, Inc.; Biogen; Bristol-Myers Squibb Company; CereSpir, Inc.; Cogstate; Eisai Inc.; Elan Pharmaceuticals, Inc.; Eli Lilly and Company; EuroImmun; F. Hoffmann-La Roche Ltd and its affiliated company Genentech, Inc.; Fujirebio; GE Healthcare; IXICO Ltd.;Janssen Alzheimer Immunotherapy Research & Development, LLC.; Johnson & Johnson Pharmaceutical Research & Development LLC.; Lumosity; Lundbeck; Merck & Co., Inc.;Meso Scale Diagnostics, LLC.; NeuroRx Research; Neurotrack Technologies; Novartis Pharmaceuticals Corporation; Pfizer Inc.; Piramal Imaging; Servier; Takeda Pharmaceutical Company; and Transition Therapeutics. The Canadian Institutes of Health Research is providing funds to support ADNI clinical sites in Canada. Private sector contributions are facilitated by the Foundation for the National Institutes of Health (www.fnih.org). The grantee organization is the Northern California Institute for Research and Education, and the study is coordinated by the Alzheimer's Therapeutic Research Institute at the University of Southern California. ADNI data are disseminated by the Laboratory for Neuro Imaging at the University of Southern California; Brain tissue and additional biological samples were collected from individuals who were assessed and followed in the Case Western ADC which was funded by NIA P50AG008012; Estudio Familiar de Influencia Genetica en Alzheimer (EFIGA): 5R37AG015473, RF1AG015473, R56AG051876; the Health & Aging Brain Study – Health Disparities (HABS-HD), supported by the National Institute on Aging of the National Institutes of Health under Award Numbers R01AG054073, R01AG058533, R01AG070862, P41EB015922, and U19AG078109; brain tissues were obtained from the Brain Bank at the John P. Hussman Institute for Human Genomics at the University of Miami Miller School of Medicine, with funding provided by Louis D. Scientific Prize from the Institut de France; the Indiana Alzheimer's Disease Research Center (IADRC) supported by National Institutes of Health (NIH) grant P30AG072976; the Korean Brain Aging Study for the Early Diagnosis and Prediction of Alzheimer's disease (KBASE), which was supported by a grant from Ministry of Science, ICT and Future Planning (Grant No: NRF-2014M3C7A1046042); Memory & Aging Project at Knight Alzheimer's Disease Research Center (MAP at Knight ADRC): The Memory and Aging Project at the Knight-ADRC (Knight-ADRC). This work was supported by the National Institutes of Health (NIH) grants R01AG064614, R01AG044546, RF1AG053303, RF1AG058501, U01AG058922 and R01AG064877 to Carlos Cruchaga. The recruitment and clinical characterization of research participants at Washington University was supported by NIH grants P30AG066444, P01AG03991, and P01AG026276. Data collection and sharing for this project was supported by NIH grants RF1AG054080, P30AG066462, R01AG064614 and U01AG052410. We thank the contributors who collected samples used in this study, as well as patients and their families, whose help and participation made this work possible. This work was supported by access to equipment made possible by the Hope Center for Neurological Disorders, the Neurogenomics and Informatics Center (NGI: https://neurogenomics.wustl.edu/) and the Departments of Neurology and Psychiatry at Washington University School of Medicine; the University of Miami Brain Endowment Bank (Miami Brain Bank); National Alzheimer's Coordinating Center (NACC): The NACC database is funded by NIA/NIH Grant U24 AG072122. SCAN is a multi-institutional project that was funded as a U24 grant (AG067418) by the National Institute on Aging in May 2020. Data collected by SCAN and shared by NACC are contributed by the NIA-funded ADRCs as follows: P30 AG062429 (PI James Brewer, MD, PhD), P30 AG066468 (PI Oscar Lopez, MD), P30 AG062421 (PI Bradley Hyman, MD, PhD), P30 AG066509 (PI Thomas Grabowski, MD), P30 AG066514 (PI Mary Sano, PhD), P30 AG066530 (PI Helena Chui, MD), P30 AG066507 (PI Marilyn Albert, PhD), P30 AG066444 (PI John Morris, MD), P30 AG066518 (PI Jeffrey Kaye, MD), P30 AG066512 (PI Thomas Wisniewski, MD), P30 AG066462 (PI Scott Small, MD), P30 AG072979 (PI David Wolk, MD), P30 AG072972 (PI Charles DeCarli, MD), P30 AG072976 (PI Andrew Saykin, PsyD), P30 AG072975 (PI David Bennett, MD), P30 AG072978 (PI Neil Kowall, MD), P30 AG072977 (PI Robert Vassar, PhD), P30 AG066519 (PI Frank LaFerla, PhD), P30 AG062677 (PI Ronald Petersen, MD, PhD), P30 AG079280 (PI Eric Reiman, MD), P30 AG062422 (PI Gil Rabinovici, MD), P30 AG066511 (PI Allan Levey, MD, PhD), P30 AG072946 (PI Linda Van Eldik, PhD), P30 AG062715 (PI Sanjay Asthana, MD, FRCP), P30 AG072973 (PI Russell Swerdlow, MD), P30 AG066506 (PI Todd Golde, MD, PhD), P30 AG066508 (PI Stephen Strittmatter, MD, PhD), P30 AG066515 (PI Victor Henderson, MD, MS), P30 AG072947 (PI Suzanne Craft, PhD), P30 AG072931 (PI Henry Paulson, MD, PhD), P30 AG066546 (PI Sudha Seshadri, MD), P20 AG068024 (PI Erik Roberson, MD, PhD), P20 AG068053 (PI Justin Miller, PhD), P20 AG068077 (PI Gary Rosenberg, MD), P20 AG068082 (PI Angela Jefferson, PhD), P30 AG072958 (PI Heather Whitson, MD), P30 AG072959 (PI James Leverenz, MD); National Institute on Aging Alzheimer's Disease Family Based Study (NIA-AD FBS): U24 AG056270; Religious Orders Study (ROS): P30AG10161,R01AG15819, R01AG42210; Memory and Aging Project (MAP - Rush): R01AG017917, R01AG42210; Minority Aging Research Study (MARS): R01AG22018, R01AG42210; the Texas Alzheimer's Research and Care Consortium (TARCC), funded by the Darrell K Royal Texas Alzheimer's Initiative, directed by the Texas Council on Alzheimer's Disease and Related Disorders; Washington Heights/Inwood Columbia Aging Project (WHICAP): RF1 AG054023;and Wisconsin Registry for Alzheimer's Prevention (WRAP): R01AG027161 and R01AG054047. Additional acknowledgments include the National Institute on Aging Genetics of Alzheimer's Disease Data Storage Site (NIAGADS, U24AG041689) at the University of Pennsylvania, funded by NIA.

**Alzheimer's Disease Neuroimaging Initiative (sa000002)**

ADNI investigators designed and conducted the source study and collected the data used here. ADNI-derived data were accessed through ADSP and the corresponding ADSP-PHC phenotypes. ADNI was established as a public-private partnership to investigate progression from MCI to AD using longitudinal imaging, biomarkers and clinical and neuropsychological assessments. The investigators are listed in the official ADNI acknowledgement list.

Current ADNI data collection and sharing are supported by the National Institute on Aging (U19 AG024904). The historical resource-specific funding statement is retained below.

Data collection and sharing for this project was funded by the Alzheimer's Disease Neuroimaging Initiative (ADNI) (National Institutes of Health Grant U01 AG024904) and DOD

ADNI (Department of Defense award number W81XWH-12-2-0012). ADNI is funded by the National Institute on Aging, the National Institute of Biomedical Imaging and Bioengineering, and through generous contributions from the following: AbbVie, Alzheimer's Association; Alzheimer's Drug Discovery Foundation; Araclon Biotech; BioClinica, Inc.; Biogen; Bristol-Myers Squibb Company; CereSpir, Inc.; Cogstate; Eisai Inc.; Elan Pharmaceuticals, Inc.; Eli Lilly and Company; EuroImmun; F. Hoffmann-La Roche Ltd and its affiliated company Genentech, Inc.; Fujirebio; GE Healthcare; IXICO Ltd.; Janssen Alzheimer Immunotherapy Research & Development, LLC.; Johnson & Johnson Pharmaceutical Research & Development LLC.; Lumosity; Lundbeck; Merck & Co., Inc.; Meso Scale Diagnostics, LLC.; NeuroRx Research; Neurotrack Technologies; Novartis Pharmaceuticals Corporation; Pfizer Inc.; Piramal Imaging; Servier; Takeda Pharmaceutical Company; and Transition Therapeutics. The Canadian Institutes of Health Research is providing funds to support ADNI clinical sites in Canada. Private sector contributions are facilitated by the Foundation for the National Institutes of Health (www.fnih.org). The grantee organization is the Northern California Institute for Research and Education, and the study is coordinated by the Alzheimer's Therapeutic Research Institute at the University of Southern California. ADNI data are disseminated by the Laboratory for Neuro Imaging at the University of Southern California.

The ADNI Data Use Agreement provides the source-attribution and publication requirements.

**The Familial Alzheimer Sequencing Project (sa000004)**

This work was supported by grants from the National Institutes of Health (R01AG044546, P01AG003991, RF1AG053303, R01AG058501, U01AG058922, RF1AG058501 and R01AG057777). The recruitment and clinical characterization of research participants at Washington University were supported by NIH P50 AG05681, P01 AG03991, and P01 AG026276. This work was supported by access to equipment made possible by the Hope Center for Neurological Disorders, and the Departments of Neurology and Psychiatry at Washington University School of Medicine.

We thank the contributors who collected samples used in this study, as well as patients and their families, whose help and participation made this work possible. This work was supported by access to equipment made possible by the Hope Center for Neurological Disorders, and the Departments of Neurology and Psychiatry at Washington University School of Medicine

**Washington Heights and Inwood Community Aging project (WHICAP) (sa000007)**

Data collection and sharing for this project was supported by the Washington Heights-Inwood Columbia Aging Project (WHICAP R01AG072474, PO1AG07232, R01AG037212, RF1AG054023) funded by the National Institute on Aging (NIA). We acknowledge the WHICAP study participants and the WHICAP research and support staff for their contributions to this study.

**Charles F. and Joanne Knight Alzheimer's Disease Research Center (sa000008)**

The Memory and Aging Project at the Knight-ADRC (Knight-ADRC), supported by NIH grants R01AG064614, R01AG044546, RF1AG053303, RF1AG058501, U01AG058922 and R01AG064877 to Carlos Cruchaga. The recruitment and clinical characterization of research participants at Washington University was supported by NIH grants P30AG066444, P01AG03991, and P01AG026276. Data collection and sharing for this project was supported by NIH grants RF1AG054080, P30AG066462, R01AG064614 and U01AG052410. This work was supported by access to equipment made possible by the Hope Center for Neurological Disorders, the Neurogenomics and Informatics Center (NGI: https://neurogenomics.wustl.edu/) and the Departments of Neurology and Psychiatry at Washington University School of Medicine.

**Accelerating Medicines Partnership-Alzheimer's Disease (AMP-AD) (sa000011)**

We are grateful to the participants in the Religious Order Study, the Memory and Aging Project. This work is supported by the US National Institutes of Health [U01 AG046152, R01 AG043617, R01 AG042210, R01 AG036042, R01 AG036836, R01 AG032990, R01 AG18023, RC2 AG036547, P50 AG016574, U01 ES017155, KL2 RR024151, K25 AG041906-01, R01 AG30146, P30 AG10161, R01 AG17917, R01 AG15819, K08 AG034290, P30 AG10161 and R01 AG11101.

**APOE Extremes WGS Study (sa000013)**

We would like to thank study participants, their families, and the sample collectors for their invaluable contributions. This research was supported in part by the National Institute on Aging grant U01AG049508 (PI Alison M. Goate) and U01AG052411 (MPIs Alison Goate, Carlos Cruchaga, Bin Zhang). This research was supported in part by the National Institute on Aging Intramural Program (MPIs Sonja Scholz, Bryan Traynor, Andrew Singleton). This research was supported in part by Genentech, Inc. (PI Alison Goate, Robert Graham).

The NACC database is funded by NIA/NIH Grant U01 AG016976. NACC data are contributed by these NIA-funded ADCs: P30 AG013846 (PI Neil Kowall, MD), P50 AG008702 (PI Scott Small, MD), P50 AG025688 (PI Allan Levey, MD, PhD), P30 AG010133 (PI Andrew Saykin, PsyD), P50 AG005146 (PI Marilyn Albert, PhD), P50 AG005134 (PI Bradley Hyman, MD, PhD), P50 AG016574 (PI Ronald Petersen, MD, PhD), P30 AG013854 (PI M. Marsel Mesulam, MD), P30 AG008017 (PI Jeffrey Kaye, MD), P30 AG010161 (PI David Bennett, MD), P30 AG010129 (PI Charles DeCarli, MD), P50 AG016573 (PI Frank LaFerla, PhD), P50 AG005131 (PI Douglas Galasko, MD), P30 AG028383 (PI Linda Van Eldik, PhD), P30 AG010124 (PI John Trojanowski, MD, PhD), P50 AG005142 (PI Helena Chui, MD), P30 AG012300 (PI Roger Rosenberg, MD), P50 AG005136 (PI Thomas Grabowski, MD), P50 AG005681 (PI John Morris, MD), P30 AG028377 (Kathleen Welsh-Bohmer, PhD), and P50 AG008671 (PI Henry Paulson, MD, PhD).

Samples from the National Cell Repository for Alzheimer's Disease (NCRAD), which receives government support under a cooperative agreement grant (U24 AG21886) awarded by the National Institute on Aging (NIA), were used in this study. We thank contributors who collected samples used in this study, as well as patients and their families, whose help and participation made this work possible.

The Alzheimer's Disease Genetics Consortium supported the collection of samples used in this study through National Institute on Aging (NIA) grants U01AG032984 and RC2AG036528.

**NIH, CurePSP and Tau Consortium PSP WGS (sa000015)**

This project was funded by the NIH grant UG3NS104095 and supported by grants U54NS100693 and U54AG052427. Queen Square Brain Bank is supported by the Reta Lila Weston Institute for Neurological Studies and the Medical Research Council UK. The Mayo Clinic Florida had support from a Morris K. Udall Parkinson's Disease Research Center of

Excellence (NINDS P50 #NS072187), CurePSP and the Tau Consortium. The samples from the University of Pennsylvania are supported by NIA grant P01AG017586.

**Dissecting the Genomic Etiology of non-Mendelian Early-Onset Alzheimer Disease (EOAD) and Related Phenotypes (sa000023)**

This work was supported by the National Institutes of Health (NIH) grant R01AG064614. The ADSP-FUS is supported by U01AG057659.

The National Institutes of Health, National Institute on Aging (NIH-NIA) supported this work through the following grants: ADGC, U01 AG032984, RC2 AG036528; samples from the National Centralized Repository for Alzheimer's Disease and Related Dementias (NCRAD), which receives government support under a cooperative agreement grant (U24 AG21886) awarded by the National Institute on Aging (NIA), were used in this study. Sequencing data generation and harmonization is supported by the Genome Center for Alzheimer's Disease, U54AG052427, and data sharing is supported by NIAGADS, U24AG041689. We thank contributors who collected samples used in this study, as well as patients and their families, whose help and participation made this work possible.

NIH grants supported enrollment and data collection for the individual studies including the Alzheimer's Disease Centers (ADC, P30 AG062429 (PI James Brewer, MD, PhD), P30 AG066468 (PI Oscar Lopez, MD), P30 AG062421 (PI Bradley Hyman, MD, PhD), P30 AG066509 (PI Thomas Grabowski, MD), P30 AG066514 (PI Mary Sano, PhD), P30 AG066530 (PI Helena Chui, MD), P30 AG066507 (PI Marilyn Albert, PhD), P30 AG066444 (PI John Morris, MD), P30 AG066518 (PI Jeffrey Kaye, MD), P30 AG066512 (PI Thomas Wisniewski, MD), P30 AG066462 (PI Scott Small, MD), P30 AG072979 (PI David Wolk, MD), P30 AG072972 (PI Charles DeCarli, MD), P30 AG072976 (PI Andrew Saykin, PsyD), P30 AG072975 (PI David Bennett, MD), P30 AG072978 (PI Neil Kowall, MD), P30 AG072977 (PI Robert Vassar, PhD), P30 AG066519 (PI Frank LaFerla, PhD), P30 AG062677 (PI Ronald Petersen, MD, PhD), P30 AG079280 (PI Eric Reiman, MD), P30 AG062422 (PI Gil Rabinovici, MD), P30 AG066511 (PI Allan Levey, MD, PhD), P30 AG072946 (PI Linda Van Eldik, PhD), P30 AG062715 (PI Sanjay Asthana, MD, FRCP), P30 AG072973 (PI Russell Swerdlow, MD), P30 AG066506 (PI Todd Golde, MD, PhD), P30 AG066508 (PI Stephen Strittmatter, MD, PhD), P30 AG066515 (PI Victor Henderson, MD, MS), P30 AG072947 (PI Suzanne Craft, PhD), P30 AG072931 (PI Henry Paulson, MD, PhD), P30 AG066546 (PI Sudha Seshadri, MD), P20 AG068024 (PI Erik Roberson, MD, PhD), P20 AG068053 (PI Justin Miller, PhD), P20 AG068077 (PI Gary Rosenberg, MD), P20 AG068082 (PI Angela Jefferson, PhD), P30 AG072958 (PI Heather Whitson, MD), P30 AG072959 (PI James Leverenz, MD). The Miami ascertainment and research were supported in part through: RF1AG054080, R01AG027944, R01AG019085, R01AG028786-02, RC2AG036528. The Columbia ascertainment and research were supported in part through: R37AG015473 and U24AG056270. The University of Washington ascertainment and research were supported in part through R01AG044546, RF1AG053303, RF1AG058501, U01AG058922 and R01AG064877.

**Genetic Architecture of Alzheimer's disease and Related Proteinopathies (sa000059)**

This work was made possible by the support from the National Institutes of Health grants:

PITT: AG064877, AG030653, AG041718, P30-AG066468, U01 AT000162, AG023651, AG052521, AG025516, UF1 AG051197, P01 AG025204, RF1 AG052525, AG052446.

WASHU: R01AG044546, P01AG003991, RF1AG053303, R01AG058501, U01AG058922, RF1AG058501 and R01AG057777). The recruitment and clinical characterization of research participants at Washington University were supported by NIH P50 AG05681, P01 AG03991, and P01 AG026276. This work was supported by access to equipment made possible by the Hope Center for Neurological Disorders, and the Departments of Neurology and Psychiatry at Washington University School of Medicine.

We thank the contributors who collected samples used in this study, as well as patients and their families, whose help and participation made this work possible. This work was supported by access to equipment made possible by the Hope Center for Neurological Disorders, and the Departments of Neurology and Psychiatry at Washington University School of Medicine.

**National Institute of Aging Alzheimer's Disease Family Based Study (NIA AD-FBS) (sa000063)**

The NIA-AD FBS study supported the collection of samples used in this study through National Institute on Aging (NIA) grants U24AG056270,U24AG026395, R01AG041797 and U24AG056270. We thank contributors, including the Alzheimer's disease Centers who collected samples used in this study, as well as patients and their families, whose help and participation made this work possible.

**National Alzheimer's Coordinating Center**

The NACC database is funded by NIA/NIH Grant U24 AG072122. NACC data are contributed by the NIA-funded ADRCs: P30 AG062429 (PI James Brewer, MD, PhD), P30 AG066468 (PI Oscar Lopez, MD), P30 AG062421 (PI Teresa Gomez-Isla, MD), P30 AG066509 (PI Thomas Grabowski, MD), P30 AG066514 (PI Mary Sano, PhD), P30 AG066530 (PI Helena Chui, MD, Arthur Toga, PhD), P30 AG066507 (PI Marilyn Albert, PhD), P30 AG066444 (PI David Holtzman, MD), P30 AG066518 (PIs Lisa Silbert, MD, Kevin Duff, PhD), P30 AG066512 (PI Thomas Wisniewski, MD), P30 AG066462 (PI Scott Small, MD), P30 AG072979 (PI David Wolk, MD), P30 AG072972 (PIs Charles DeCarli, MD, Rachel Whitmer, PhD), P30 AG072976 (PI Andrew Saykin, PsyD), P30 AG072975 (PI Julie Schneider, MD, MS), P30 AG072978 (PI Ann McKee, MD), P30 AG072977 (PI Robert Vassar, PhD), P30 AG066519 (PI Joshua Grill, PhD), P30 AG062677 (PIs Brad Boeve, MD, Ronald Petersen, MD, PhD), P30 AG079280 (PI Jessica Langbaum, PhD), P30 AG062422 (PI Gil Rabinovici, MD), P30 AG066511 (PI Allan Levey, MD, PhD), P30 AG072946 (PI Linda Van Eldik, PhD), P30 AG062715 (PI Sanjay Asthana, MD, FRCP), P30 AG072973 (PI Russell Swerdlow, MD), P30 AG066506 (PIs Glenn Smith, PhD, ABPP, David Lowenstein, PhD, Ranjan Duara, MD), P30 AG066508 (PIs Stephen Strittmatter, MD, PhD, Christopher Van Dyck, MD), P30 AG066515 (PI Victor Henderson, MD, MS), P30 AG072947 (PI Suzanne Craft, PhD), P30 AG072931 (PI Henry Paulson, MD, PhD), P30 AG066546 (PIs Sudha Seshadri, MD, Gladys Maestre, MD, PhD), P30 AG086401 (PI Erik Roberson, MD, PhD), P30 AG086404 (PI Gary Rosenberg, MD), P30 AG086403 (PI Angela Jefferson, PhD), P30 AG072958 (PIs Heather Whitson, MD, Gwenn Garden, MD, PhD), P30 AG072959 (PI Jagan Pillai, MD, PhD), P30 AG092752 (Ihab Hajjar, MD, MS).
